# Initial Description and Demonstration of a Family of Methods with the Potential to Approximate Confounding

**DOI:** 10.1101/2025.10.30.25339088

**Authors:** Eric G. Smith, Dane M. Netherton

## Abstract

Confounding is one of the most important concerns for randomized or nonrandomized intervention or exposure studies. This manuscript describes several metrics intended to provide quantitative approximations of confounding under certain conditions. Each metric quantifies differences in risk between intervention arms during time periods when the intervention (or exposure of interest) *is not occurring*. Because exposure is absent, these metrics have the potential to summarize the effects of other measured and unmeasured factors on outcome risk. A null association (e.g., a risk, rate, or hazard ratio of approximately 1.0 or a risk difference of 0.0) between intervention arms during nonexposure can suggest equal impacts from baseline factors affecting each study arm (i.e., an absence of confounding). However, other factors such as attrition bias, postexposure effects, or incomplete representation of the full cohort can also affect risks during nonexposure, causing the nonexposure risk to represent confounding less accurately. We propose four nonexposure metrics designed to limit these other influences on nonexposure risk, thus providing nonexposure risks that more exclusively reflect confounding. The metrics, however, vary in their potential to limit these other influences and also vary in their sensitivity to random error. We then demonstrate what we expect to be the most widely useful metric currently, the “briefly-exposed postexposure (bePE) risk metric.” We show how the bePE risk metric can inform multiple aspects of a real-world study, such as cohort derivation and interpretation of findings. Definitive validation of nonexposure risk metrics awaits further research. Nevertheless, these metrics have the potential to substantially improve intervention and exposure studies by approximating confounding under certain conditions. Their testing and validation should be a research priority.

## Introduction

Confounding is one of the most important concerns for randomized or nonrandomized intervention or exposure studies.^1–3^ Few analytic methods can address both measured and unmeasured confounding,^4,5^ and the extent to which these methods actually minimize confounding is often unclear.^6,7^

This manuscript describes and demonstrates a new set of metrics intended to approximate how much confounding, including unmeasured confounding, remains in a particular randomized or nonrandomized intervention study. (These metrics can also inform some other studies that compare groups of individuals, such as studies of environmental or occupational exposures. However, for simplicity, we will use the term “intervention studies” exclusively in this manuscript.) We briefly describe each metric and then demonstrate the metric that we expect to be the most useful currently. We also propose an initial set of quality assessments for the metric that we demonstrate and discuss additional research needs and potential future uses of these metrics.

Our proposed metrics quite significantly refine and extend a more general approach first employed in 2001.^8^ Our metrics seek to assess, rather than directly resolve, confounding. However, achieving even an imperfect quantitative approximation of confounding may significantly improve intervention studies by informing decisions about how to best resolve such confounding.

### The Concept of Nonexposure Risk as a Measure of Confounding

The basic premise of this manuscript is simple: if a study involving two or more exposed groups has successfully removed confounding, then, *if all other considerations are equal*, identical risks should be observed between the intervention arms when the exposure to the intervention is not present. More specifically, as has been described for related “negative control” approaches,^9^ a null association (e.g., a relative risk or rate-based ratio of 1.0 or a risk difference of 0.0) between the intervention arms during certain nonexposure periods may suggest an absence of confounding remaining in the analysis. (Such confounding is also termed “residual confounding.” However, for simplicity, we will exclusively use the briefer term “confounding” in this manuscript). In a sense, examining risks during nonexposure periods can provide a “window” into the sum effect of all confounding factors when the intervention is not present. This “window” can suggest whether confounding between intervention arms has been effectively resolved.

However, as will be discussed, it is crucial to evaluate the plausibility of other influences upon the risks observed during periods of nonexposure. These influences complicate the intended use of “nonexposure risks” as an indicator of confounding, since when these influences are present the nonexposure risks do not exclusively reflect confounding. “Nonexposure risk” (Supplement 1 [S1]) is the term we generally give to the risk observed during *specific time periods* of nonexposure either *immediately before or after* individuals are exposed to the study intervention. (For one of our four proposed metrics, the term “nonexposure risks” does not meet the precise definition given here but rather refers to the risk observed during the nonexposure period of individuals who were *intended to receive* the intervention but did not actually receive it.)

Because of these potential other influences on nonexposure risk, the nonexposure risk metrics proposed here are not perfect indicators of confounding. However, given the challenges presented by confounding, even if these metrics provide approximations of confounding that are sometimes imprecise, we expect that informed use of these metrics should significantly improve many intervention studies. To help facilitate the informed use of these metrics, we also propose an initial set of quality assessments for one of these metrics.

### Disadvantages of an Earlier Nonexposure Risk Metric

The concept of using nonexposure risk to assess confounding is not new (S2). To our knowledge, the first use of any nonexposure risk metric analogous to what we are proposing (i.e., assessing confounding by examining nonexposure risk in some of the same individuals used to assess risks during exposure) occurred in 2001 by Nielsen and colleagues,^8^ followed very shortly afterwards by Ray and colleagues.^10–12^ These two groups of investigators examined the risks that were observed after exposure to an intervention was completed for all the individuals who discontinued an intervention at some point during the time period under study. While conceptually important for the focus it put on nonexposure risk as a potential indicator of confounding, this ““all discontinuations” postexposure (adPE) risk metric,” as we call this indicator, has some distinct limitations. In particular, the adPE risk includes the effects of both confounding and attrition bias.

Attrition bias arises from differences between a study’s intervention arms in the individuals who stop the intervention (or have the intervention stopped) and those who continue receiving the intervention. This pronounced sensitivity to attrition bias actually poses a major drawback for the adPE risk metric. We provide a detailed discussion of this issue in S3. In brief, the issue relates to the concern that when confounding and attrition bias act to alter the effect estimate *in the same direction,* they would be expected to affect the postexposure nonexposure risk *in opposite directions* (and vice versa). Thus, if confounding and attrition bias are both present, then the adPE nonexposure risk metric would be expected to either underrepresent or overrepresent confounding.

Since the adPE risk metric generally provides substantial time for attrition bias to occur during the actual exposure to the intervention, this metric could easily misestimate confounding (especially for studies with high rates of attrition during the follow-up period).^13,14^ Similarly, the longer period of exposure allowed by this metric also often provides substantial time for one intervention to produce greater benefits or risks than the other by the time an individual discontinues the intervention. Such intervention-related benefits or risks often can extend into the period after exposure has ended. These “postexposure intervention-related effects” (S3) provide another reason besides confounding and attrition bias that nonexposure risks metrics may diverge from the null.

### Our Proposed Nonexposure Risk Metrics

Table 1 (and Figure 1A and 1B) presents the four new types of nonexposure risk metrics that we are proposing as improvements on the adPE risk metric. All these metrics deliberately either minimize or entirely avoid prior exposure to the intervention so as to minimize or eliminate attrition bias and/or postexposure intervention-related effects, as well as minimizing or eliminating effects from another phenomenon called “depletion-of-susceptible-individuals”^15–20^ (discussed further in S4). Our proposed metrics include two new “postexposure” risk metrics (Figure 1A): 1) the “briefly-exposed postexposure (bePE) risk metric,” which examines individuals who are only briefly exposed to the intervention, and a close variant, 2) the “ultra-briefly-exposed postexposure (ubePE) risk metric,” which further restricts the sample to individuals with extremely brief exposures to the intervention. Sometimes, it may even be possible to examine the generally very small sample of individuals who are assigned to the intervention but do not actually commence it, termed the “assigned-but-not-exposed (abNE) risk metric” (Figure 1B). All three of these metrics evaluate nonexposure risks occurring over the same follow-up period that partially or completely corresponds with the follow-up period for the main analysis. Finally, the “baseline preexposure (basePreE) risk metric” examines nonexposed risk up until the point of intervention initiation (Figure 1B).

**Figure 1A:**
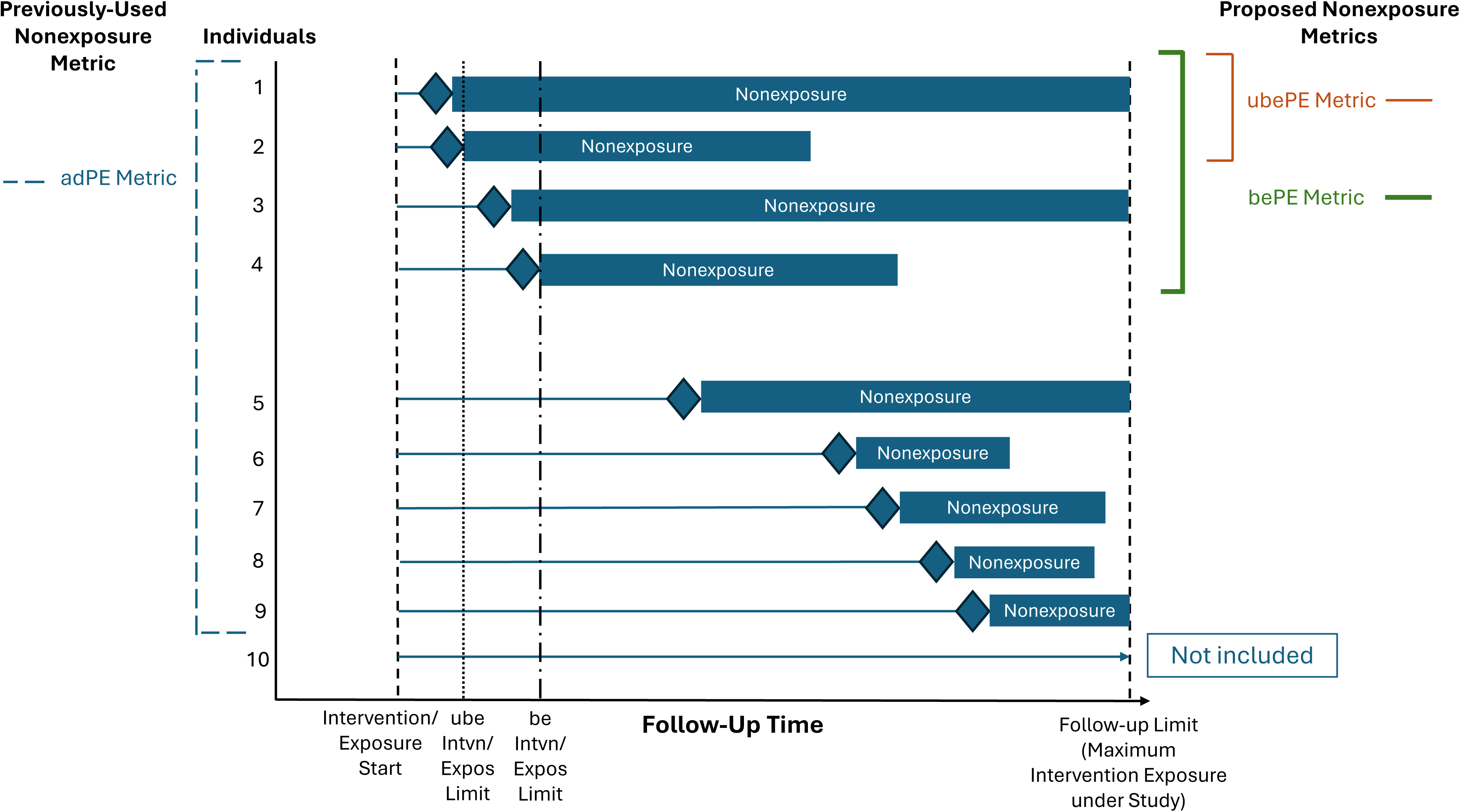
This figure depicts the nonexposure time that is included in the “all discontinuations” postexposure (adPE) risk metric, the briefly-exposed postexposure (bePE) risk metric, and the ultra-briefly-exposed postexposure (ubePE) risk metric. The key concept is that it is the period of nonexposure (the thick rectangular bar), rather than the period of exposure (the thin line), that is of interest for quantifying nonexposure risk metrics. For each individual in the figure, exposure is represented by the thin black line originating at the Intervention Start, with accruing follow-up time while exposed to the intervention being represented by the thin line progressing to the right until the exposure period (plus an allowable “grace period”) is terminated at the point represented by the diamond. The much thicker bar labeled “Nonexposure” represents the follow-up time when patients are not exposed to the Study Intervention. Not all of the bars extend to end of the study’s follow-up period (e.g., 365 days of follow-up), because some patients resume or switch their exposure or experience an outcome. The other key concept is that the adPE risk metric includes nonexposure time from all individuals who are discontinued from the intervention during the follow-up period under study. (In this example, this includes Individuals 1-9, but not Individual 10, who continues the Intervention to the end of the follow-up period [or beyond]). In contrast, the bePE or ubePE risk metrics do not include follow-up time for all the individuals being discontinued from the intervention. Rather, the bePE risk metric only includes the nonexposure time observed for those individuals who are only briefly exposed to the intervention (individuals 1-4), i.e., those for whom the intervention or exposure (with any “grace period”) only lasts until the “be Intvn/Expos Limit” (or before). (“be Intvn/Expos Limit” = briefly-exposed Intervention/Exposure Limit). The ubePE risk metric only includes nonexposure time from those individuals with the briefest exposures (Individuals 1-2), i.e., those for whom the intervention or exposure (plus “grace period”) only lasts until the “ube Intvn/Expos Limit” (or before). For these particular nonexposure risk metrics (also known as the “postexposure” risk metrics), all individuals contributing to the nonexposure metric already have been exposed to the intervention under study and have contributed to the main analysis findings prior to their nonexposure time commencing (in contrast to the nonexposure risk metrics shown in Figure 1B).

**Figure 1B:**
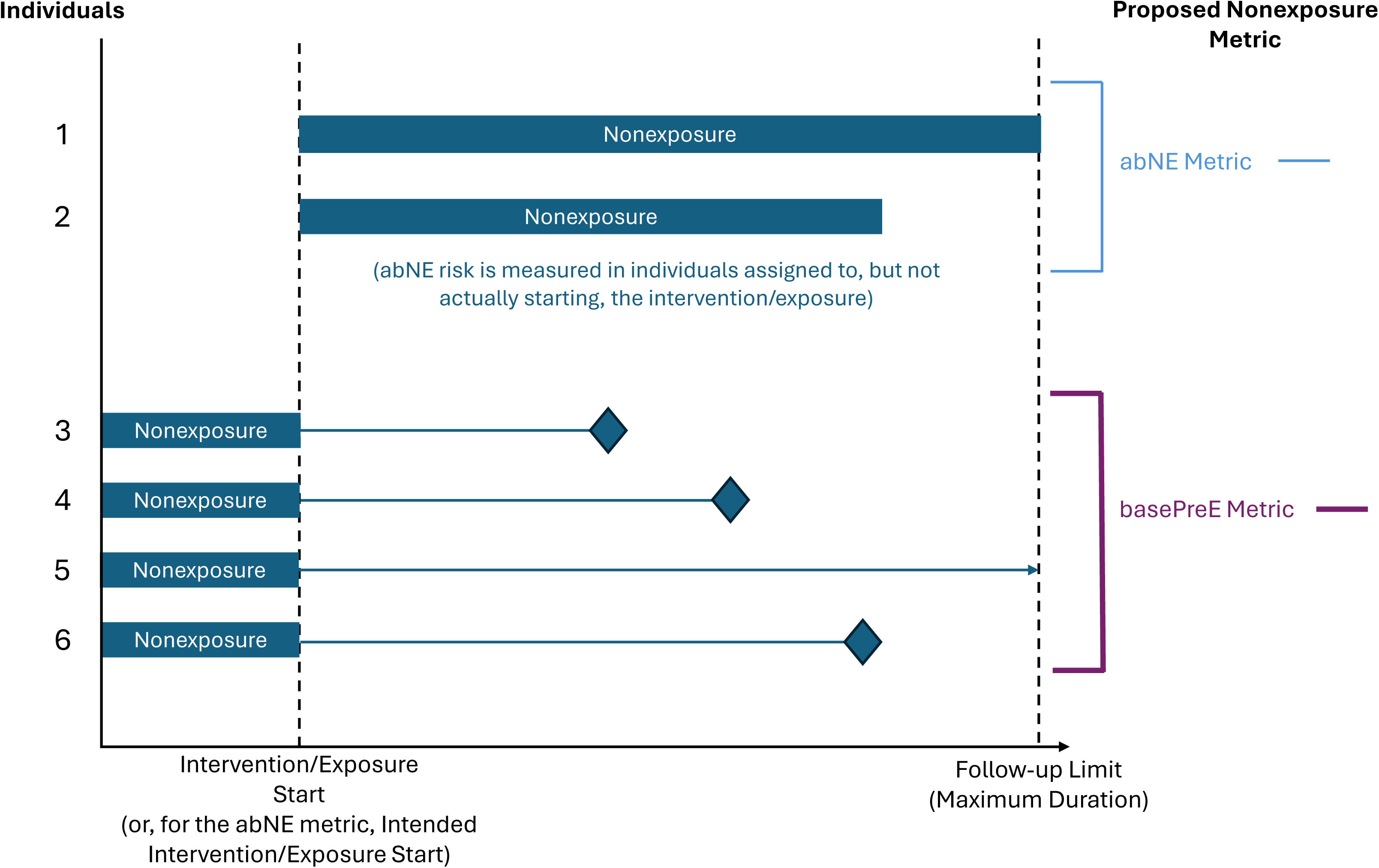
This figure depicts the two other proposed nonexposure risk metrics, the assigned-but-not-exposed (abNE) risk metric and the baseline preexposure (basePreE) risk metric. The follow-up time included in the nonexposure risk metric is represented for each individual by the thick bar labelled “Nonexposure,” as in Figure 1A. The abNE risk metric is measured in those individuals assigned to start an intervention or exposure who do not actually start that intervention or exposure. The assignment date is used as the starting date for nonexposure, and follow-up time is continued for the same time period that individuals starting the intervention are exposed (Individual 1), unless the individual subsequently starts an intervention later in this period, ending their nonexposure period, or experiences an event (Individual 2). As such, it is the only non-exposure metric to index risk across precisely the same time period that individuals starting the intervention are followed. The basePreE risk metric is restricted to, at maximum, a defined duration of time of nonexposure prior to initiating the intervention (Individuals 3-6). In this figure, for instance, the baseline preexposure period is shown as always having a uniform duration. However, if individuals are on other interventions that affect the risk for the outcome of interest prior to starting the intervention or exposure, and these are discontinued prior to the study intervention initiation date (and thus generally would not constitute “baseline confounding”), it may be advisable to not start preexposure follow-up time until individuals are no longer receiving this alternative intervention. Because risk is only measured prior to the exposure, the basePreE risk metric is the only nonexposure risk metric that can include individuals who never discontinue the intervention during the follow-up time (Individual 5). In addition, if valid baseline measurements exist for all cohort members, it can potentially include nonexposure risk from all cohort members contributing to the main analysis.

**Table 1.**
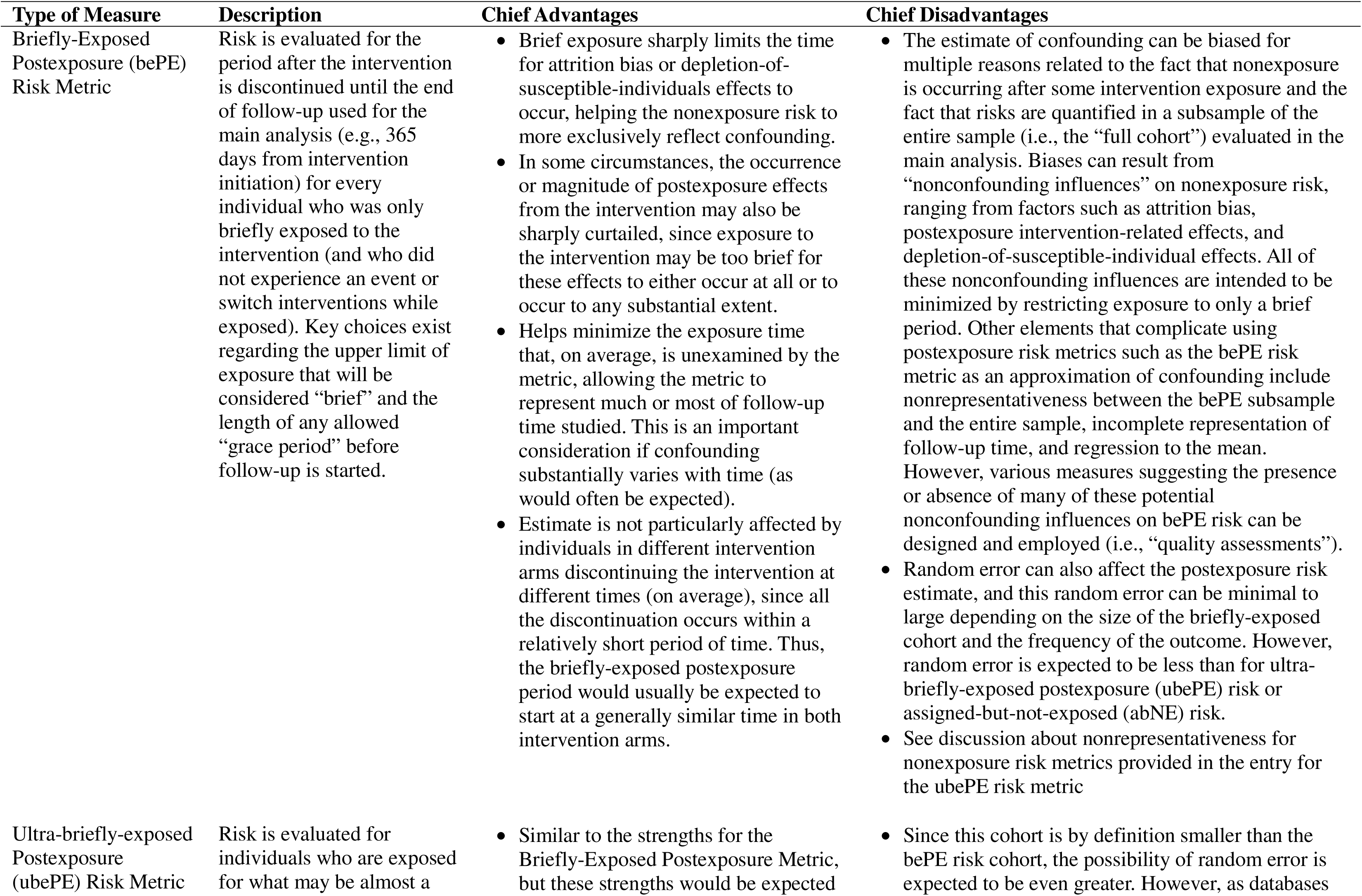
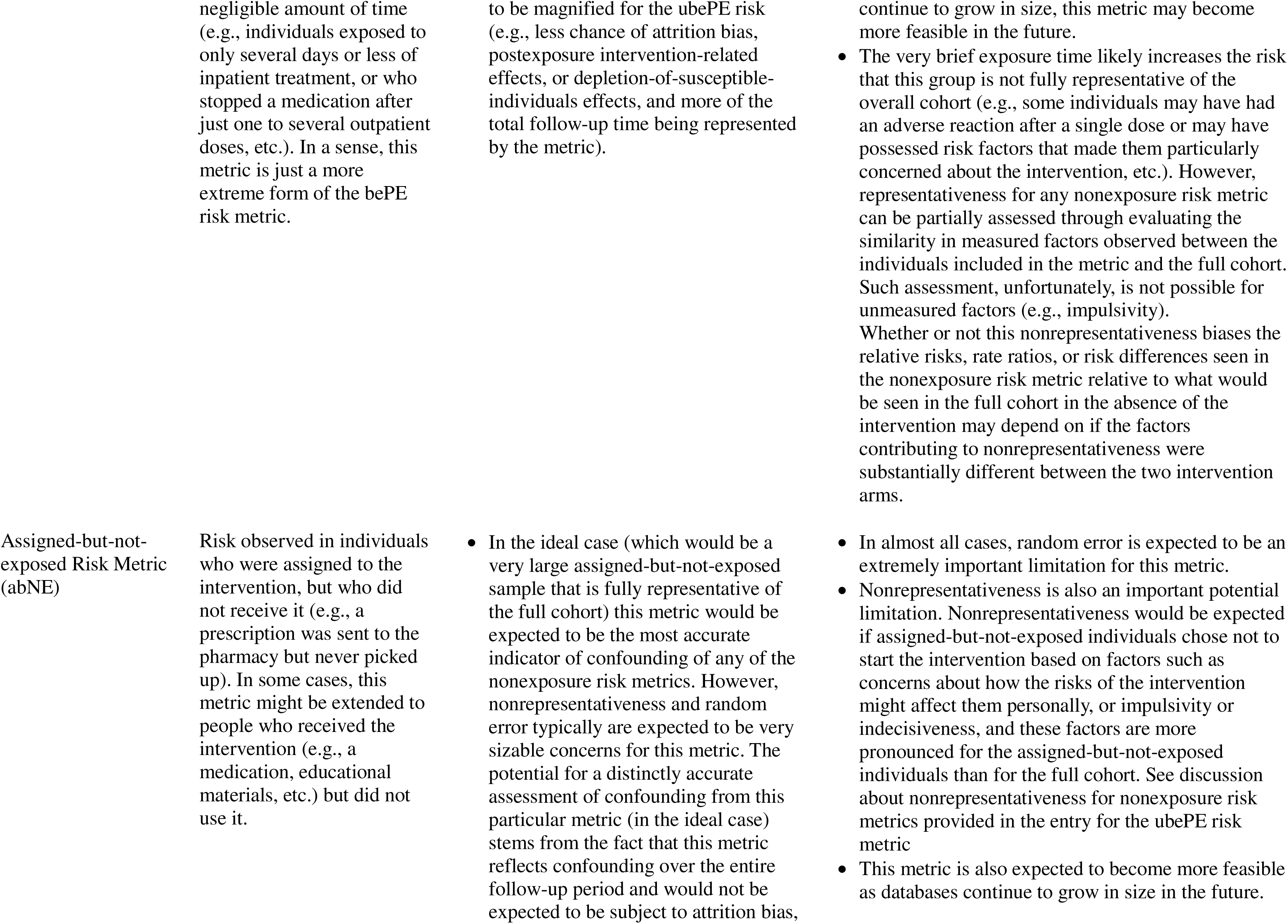
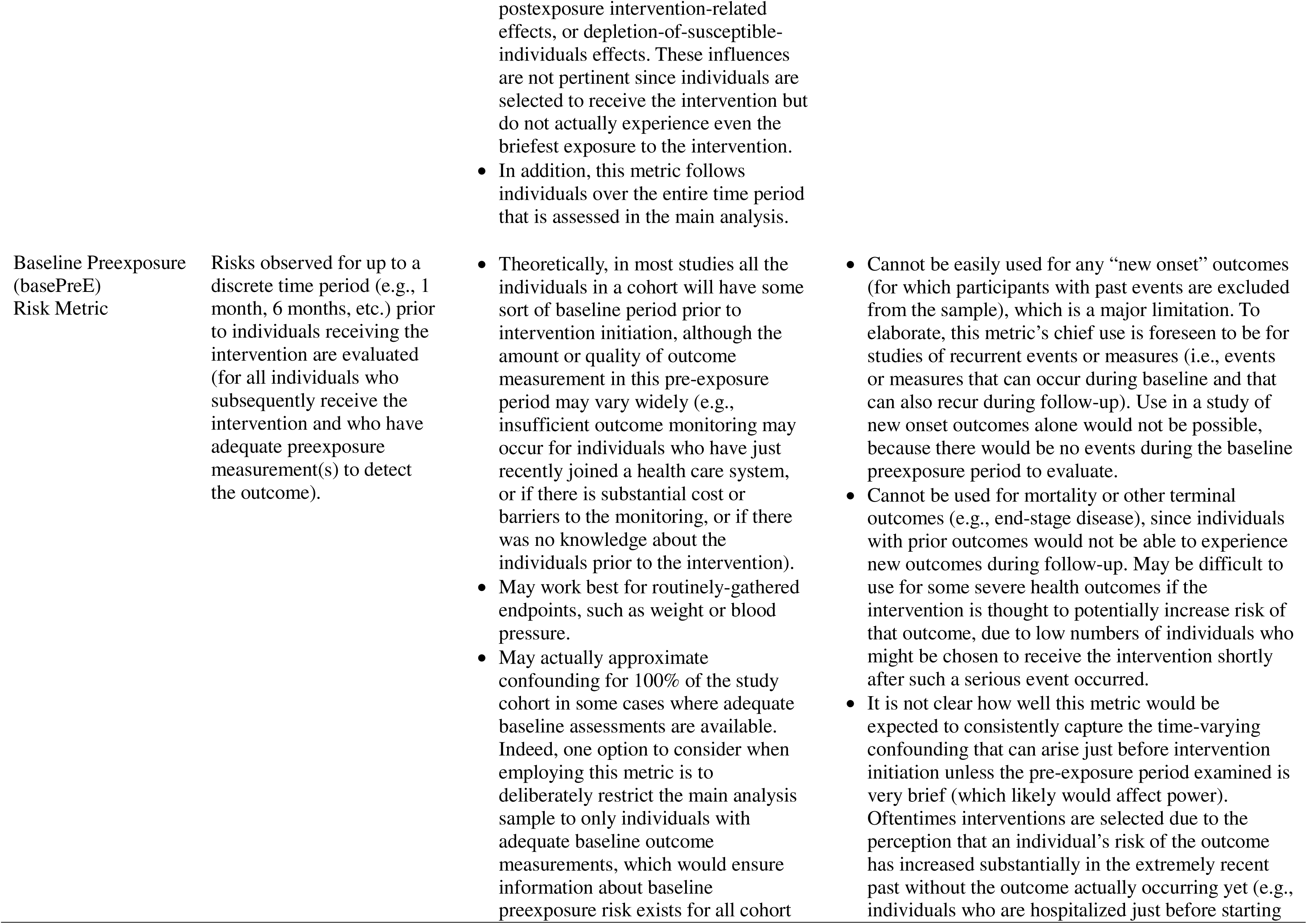
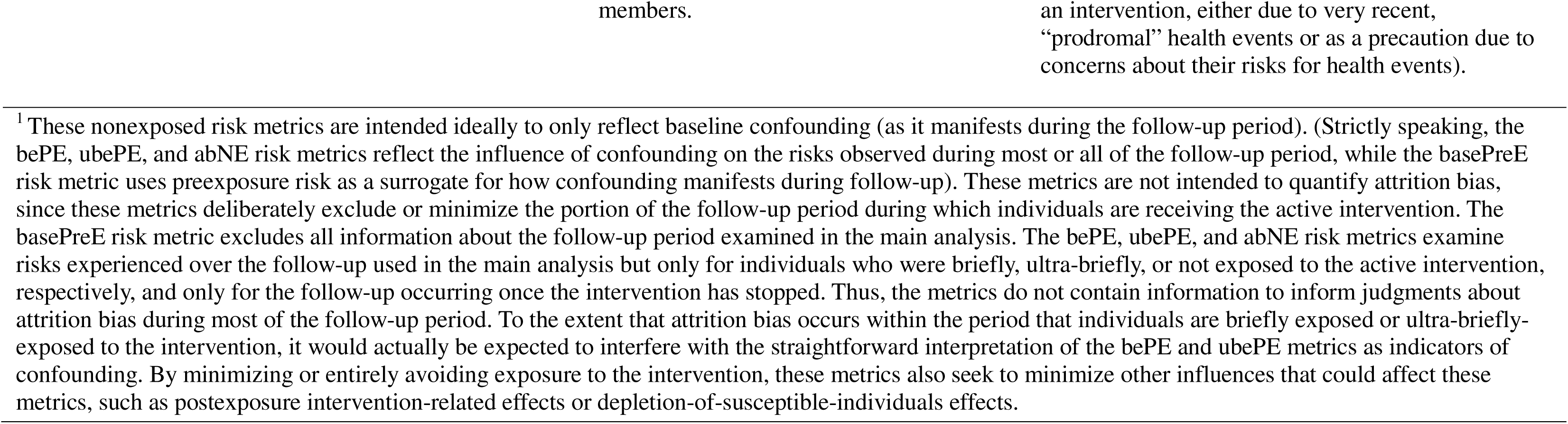
Proposed Nonexposure Risk Metrics to Approximate Confounding^1^ while Addressing Limitations in the “All Discontinuations” Postexposure (adPE) Risk Metric.

### General Considerations for Nonexposure Risk Metrics

These four proposed metrics vary according to when or for whom this period of nonexposure occurs. As a result, these four proposed metrics vary in their susceptibility to the influences that may distort the relationship between the metric and confounding. In addition to attrition bias, postexposure intervention-related effects, and depletion-of-susceptible-individuals effects, these influences include random error, nonrepresentativeness of the sample, incomplete representation of follow-up time (S5), and regression to the mean^21–24^ (S6). Regarding random error, some of these metrics are expected to examine such a small proportion of individuals that they may only become practical as databases grow larger in the future (S7).

One concern with examining nonexposure risk in individuals with little or no intervention exposure relates to “representativeness” or “comparability.” Generally, the individuals who experience little or no intervention exposure constitute only a minority of the individuals in each intervention arm. Therefore, these individuals may not be fully representative of the whole intervention arm. For instance, in a medication study this subsample might include individuals who are: a) particularly intolerant of side effects, or worried about certain side effects, b) prone to be impulsive, c) not enthusiastic or motivated about the intervention, or d) the least ill. Thus, nonexposure risk metrics may be comparing absolute nonexposure risks that are different than those that would be observed for the whole cohort. However, if the difference in absolute risks between the nonexposed individuals and the full cohort are similar for both intervention arms, then similar *relative* risks may still be observed. This would potentially still allow nonexposure risks to approximate confounding. Nevertheless, this possible noncomparability between the subsample experiencing a period of nonexposure and the full sample under study may be one of the biggest challenges for nonexposure risk metrics (except for, in some cases, the basePreE risk metric [see Table 1]). It also may be possible to conduct assessments that highlight when other influences on nonexposure risk outside of confounding, including random error and unrepresentativeness, are particularly likely (see below).

### Rationale for the bePE Risk metric

The remainder of this paper focuses on the bePE risk metric, which we expect to currently be the most widely-applicable of our proposed nonexposure risk metrics. Like the other proposed metrics, the bePE risk metric at least partially addresses some of the limitations of the adPE risk metric. However, the bePE risk metric is expected to possess more power (and likely be more representative) than some other nonexposure risk metrics (e.g., the ubePE and abNE risk metrics). The bePE risk metric is also expected to be applicable to more outcomes than the basePreE risk metric.

The bePE risk metric addresses four major concerns about the previously-used adPE metric by markedly limiting intervention exposure. Limiting exposure to the intervention to such a drastic extent serves to limit the time for attrition bias, postexposure intervention-related effects, and depletion-of-susceptible-individuals effects to develop. Additionally, the bePE risk metric also has the benefit that, since exposure is so brief, the nonexposure period starts very early in the follow-up period for all individuals contributing to the metric. Thus, the metric can reflect confounding occurring across much of the follow-up period (S5).

### A Real-World Demonstration of the bePE Risk Metric

We illustrate the value of the bePE risk metric by showing how it can inform multiple aspects of a healthcare database study (in this case, a medication-focused high-dimensional propensity score [hdPS] historical cohort study).

Animal and human^25^ research has reported effects of the mood stabilizer lithium that may be associated with longer life, and previous nonrandomized studies have suggested that lithium may be associated with decreased all-cause mortality risk.^26–29^ However, these earlier studies could have been confounded in ways that exaggerated lithium’s mortality benefit. For instance, since lithium can adversely affect kidney function, there may be a tendency for individuals with impaired kidney function to be initiated on other medications than lithium. Since many mortality pathways involve impaired kidney function,^30–32^ this tendency could plausibly create a confounded estimate decreasing lithium’s association with all-cause mortality.

Our high-dimensional propensity score cohort study (S8) examined whether lithium initiation was associated with a survival benefit relative to an alternative intervention, valproate. This study illustrates the value of bePE risk to informing both a study’s Design and Analysis phases. The bePE risk metric was defined for this study as the risks observed in the postexposure period for all individuals receiving a single prescription of ≤ 30 days duration. The postexposure time for these briefly-exposed individuals started once exposure to the intervention (and an allowed “grace period”) was over and ended based on the same criteria (a modified form of intent-to-treat criteria) as our primary mortality analysis. Our modified intent-to-treat criteria continued follow-up time until the end of the follow-up period under study or the end of available data, even if individuals stopped the intervention of interest, but also ended follow-up time if individuals received a new prescription of either medication (S8, S9). More restrictive nonexposure period definitions, based on ending follow-up time upon initiation of other interventions affecting outcome risk, could be considered (S10).

### Use of the bePE Risk Metric in the Study Design Phase

Table 2 shows our mortality study’s Cohort Derivation Flowchart. Besides providing standard information about cohort characteristics and size, this flowchart also reports the mortality risk observed among individuals who are briefly-exposed to lithium or valproate at each step in the cohort derivation process. We have highlighted in light gray the key elements of this flowchart: the traditional information conveyed in a Cohort Derivation Flowchart (number of individuals in each cohort) and the briefly-exposed postexposure (bePE) risk metric findings.

**Table 2.**
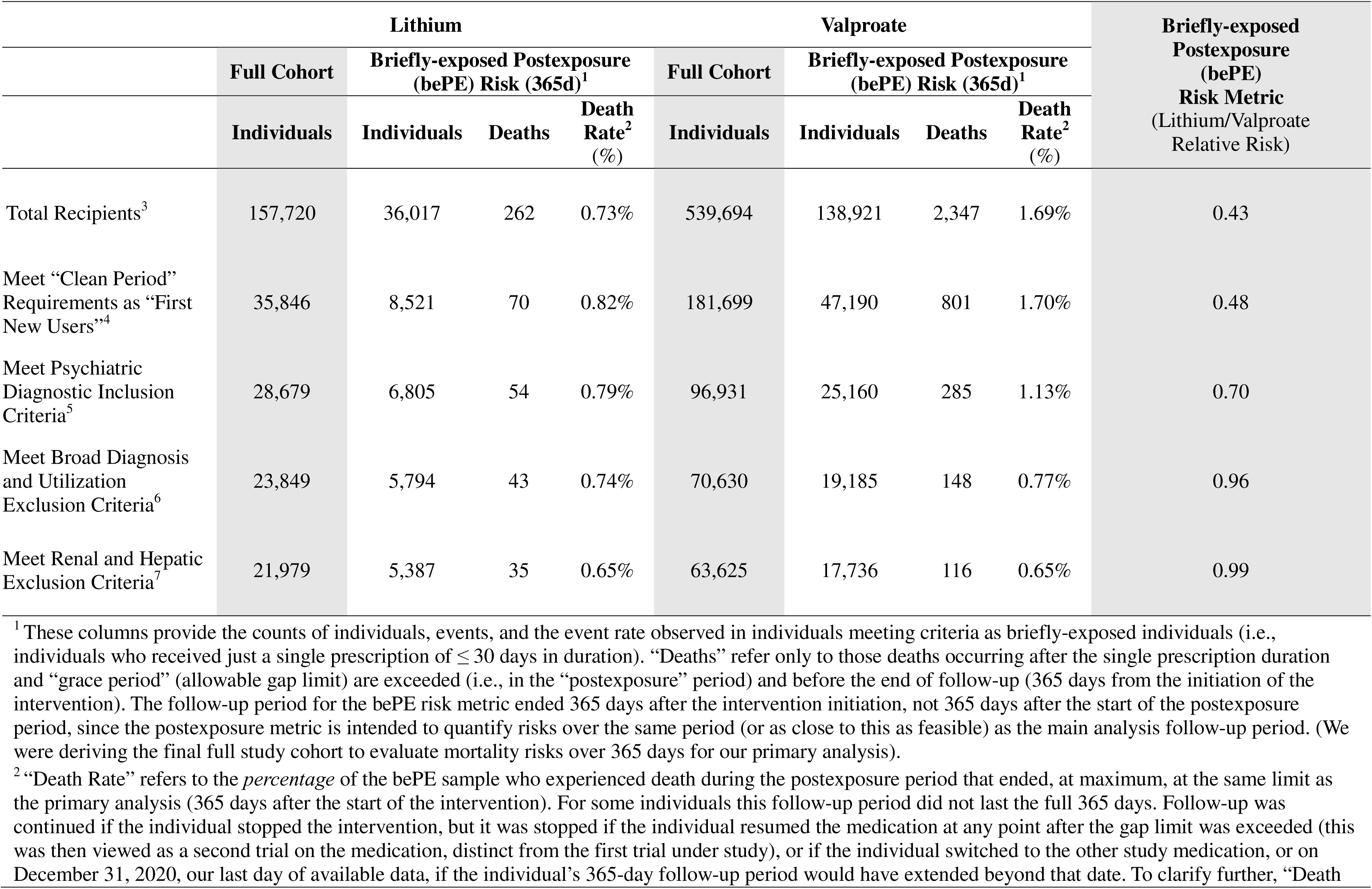
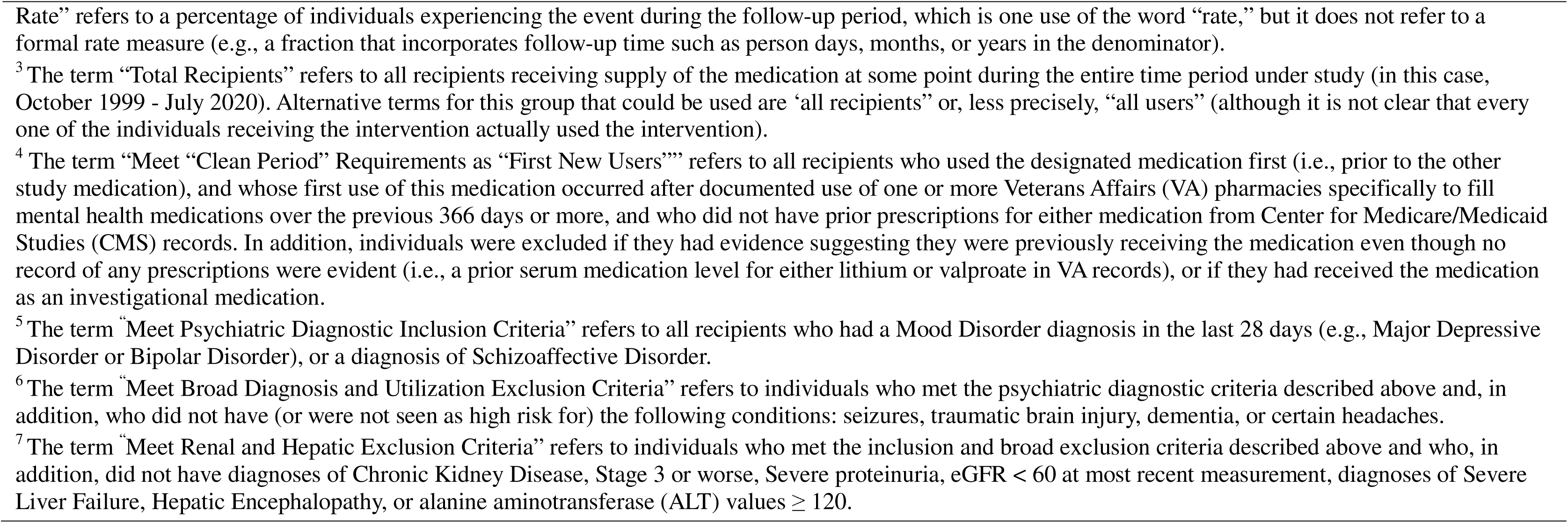
Illustration of the Use of Briefly-Exposed Postexposure (bePE) Risk Metric (Proposed Indicator of Approximate Confounding) During the Study Design Phase: Cohort Derivation Flow Chart, Lithium and Valproate Recipients, October 1999 - July 2020.

The initial cohort (“total medication recipients”) exhibited mortality risks among the briefly-exposed individuals once the intervention had been stopped that differ markedly between the lithium and valproate arms. The 365-day mortality risk observed during the nonexposure period for briefly-exposed lithium initiators was less than half of the nonexposure risk observed for briefly-exposed valproate initiators (Relative Risk [RR] central estimate = 0.43). This sharply differing risk between intervention arms among individuals no longer receiving the intervention raises concerns of substantial confounding biasing against valproate. Presumably, since these individuals are no longer exposed to the intervention, non-intervention factors are producing this difference in risk. Confounding is a prime candidate for a non-intervention factor that could plausibly be creating differences in risk observed between the two intervention arms during nonexposure. Other non-intervention factors, such as attrition bias or postexposure intervention-related effects, could also be creating differences in mortality risks between the lithium and valproate intervention arms that are observed during nonexposure. However, by drastically minimizing exposure time, it is hoped that the mortality risk among briefly-exposed individuals during nonexposure will primarily or nearly exclusively reflect confounding. The higher bePE risk observed among total recipients (or, if preferred, “all users”) of valproate may reflect valproate’s use in other higher-risk conditions than mood disorders (such as seizure disorders), or among individuals with renal function impairments.

Once we progressively applied eligibility criteria, bePE risks became much more similar between the intervention arms. Of note, after excluding individuals with relative kidney or liver contraindications, the final intervention arms exhibited highly similar central estimate bePE mortality risks (i.e., RR central estimate = 0.99). That is, our two intervention arms exhibited bePE relative risks that differed little from the target of RR = 1.0, even prior to high-dimensional propensity score matching. The bePE risk metric appears to allow for an ongoing assessment of changes in confounding that result from cohort restrictions based on logic or well-established practice,^33^ thus helping to inform judgments made during cohort derivation decision-making (S11).

Of note, because the bePE risk quantifies risks among individuals no longer exposed to the intervention, active intervention effects (or expectations about these effects) do not influence cohort derivation judgements. To the extent that bePE risk accurately approximates confounding, the impact of decisions made during the study’s Design phase can be assessed without biasing the main analysis (S12).

### Use of the bePE Risk Metric in the Study Analysis Phase

Table 3 compares our all-cause mortality survival analysis main effect and bePE risk estimates before and after hdPS matching. Results are provided for our primary (365 day) and secondary (730 day and 1825 day) all-cause mortality analyses. Prior to hdPS matching, our main analysis effect estimate for our primary mortality analysis (i.e., 365-day follow-up) does not statistically differ from the null (Hazard Ratio [HR] 0.91, 95% Confidence Interval [CI] [0.77, 1.06]). However, the 730-day (HR 0.86, 95% CI [0.76, 0.97]) and 1825-day (HR 0.88, 95% CI [0.81, 0.96]) estimates do show statistically significant lower risks of all-cause mortality associated with lithium treatment, similar to other studies.^26–28^ Importantly, the bePE risk metrics (with central estimate HRs ranging from 0.88 to 0.90) suggest that each time period’s modified intent-to-treat analysis may contain confounding that biases the effect estimate towards underestimating the mortality associated with lithium. In contrast, once hdPS matching is performed, the bePE risk metric for each time period has a value closer to the null (i.e., the central estimate HRs range from 0.93 to 0.99, although the wide confidence intervals suggest that substantial random error is possible). These findings suggest that the hdPS-matched main effect estimates may indeed be less confounded, although uncertainty still exists from random error (or possibly other influences that can affect postexposure risk, such as attrition bias or postexposure intervention-related effects, although these influences are expected to have been largely minimized). Of note, the central estimates for the main analysis effect estimates after hdPS matching are all substantially closer to the null (ranging from HR 1.00-1.02), and the 95% confidence intervals for each time period include the null.

**Table 3.** Comparison of Findings from the All-Cause Mortality Unmatched and High-dimensional Propensity Score-Matched Main Analyses and the Briefly-exposed Postexposure (bePE) Risk Metric (Proposed Indicator of Approximate Confounding)

| Unmatched Sample |  |  | High-dimensional Propensity Score Matched Sample |  |
| --- | --- | --- | --- | --- |
| Length of Follow-up (Days) | Main Analysis (mITT<sup>1</sup>) | bePE Risk Metric | Main Analysis (mITT<sup>1</sup>) | bePE Risk Metric |
|  | All-Cause Mortality Hazard Ratio (95% Confidence Interval) | All-Cause Mortality Hazard Ratio (95% Confidence Interval) | All-Cause Mortality Hazard Ratio (95% Confidence Interval) | All-Cause Mortality Hazard Ratio (95% Confidence Interval) |
| **Primary Outcome** |  |  |  |  |
| 365 | 0.91<sup>2</sup> (0.77, 1.06) | 0.90 (0.62, 1.31) | 1.01 (0.82, 1.24) | 0.93 (0.57, 1.50) |
| **Secondary Outcomes** |  |  |  |  |
| 730 | 0.86<sup>3</sup> (0.76, 0.97) | 0.88 (0.66, 1.18) | 1.00 (0.84, 1.17) | 0.99 (0.68, 1.45) |
| 1825 | 0.88<sup>4</sup> (0.81, 0.96) | 0.90 (0.76, 1.08) | 1.02 (0.91, 1.13) | 0.94 (0.75, 1.17) |

<sup>1</sup> mITT= modified ITT. The Intent-to-Treat sample is modified from a full intent-to-treat sample, but not in the manner typically done for “modified” Intent-to-Treat analyses. Rather, everyone receiving an initial prescription is included, but the modification occurs in allowing individuals to be censored before the end of follow-up if they switch to the other intervention or stop and then resume the original intervention. Typically, this is a small percentage of individuals. A more descriptive term that we sometimes apply to this analysis is to call it an “additionally-censored” ITT analysis.
<sup>2</sup> NOTE: This estimate does not correspond exactly to the estimate in the Cohort Derivation Flowchart (Table 2) because, unlike the Cohort Derivation Flowchart relative risk estimates, these hazard ratio estimates account for differences in follow-up time between the two intervention arms.
<sup>3</sup> $X^2 = 6.056$ , $p=0.014$
<sup>4</sup> $X^2 = 8.693$ , $p=0.003$

The hdPS-matched, modified intent-to-treat Kaplan-Maier survival curve shows a highly consistent similarity of mortality risk between the lithium and valproate intervention arms for up to five years of follow-up (Figure 2). Although our study’s main approaches to reducing confounding involved detailed cohort derivation and the implementation of high-dimensional propensity scores, bePE risk metric findings supported these approaches throughout the analysis.

**Figure 2:**
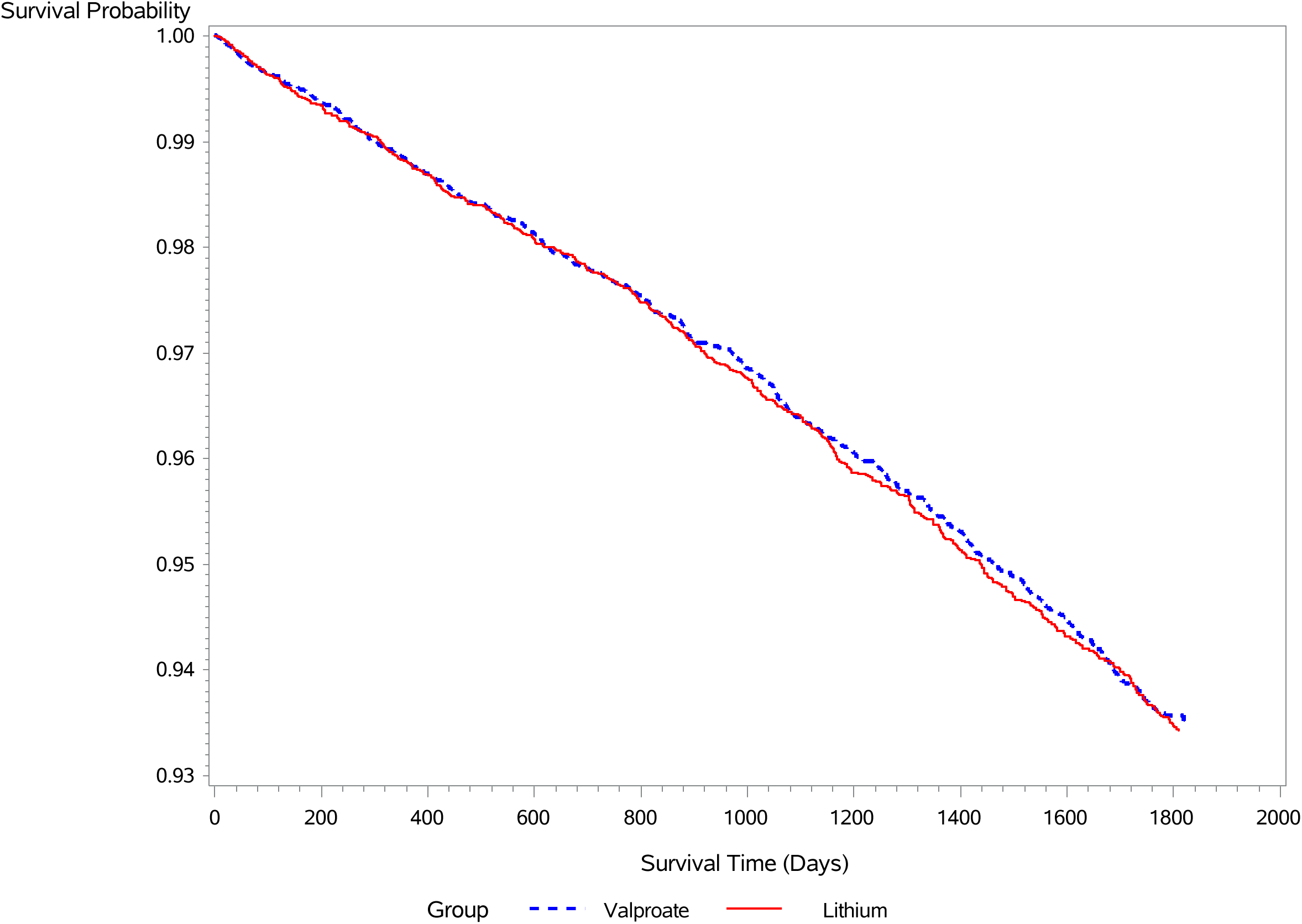
Survival plot (Kaplan-Meier plot) for the all-cause mortality outcome (modified intent-to-treat analysis) comparing lithium (solid line) and valproate (dashed line) over 1,825 days of follow-up.

Of course, we do not know with certainty the true effect of lithium on one- to five-year all-cause mortality. However, given that associations between stronger kidney function and lithium initiation are plausible and could be an overlooked source of confounding in otherwise well-performed lithium mortality studies, we strongly suspect that our study’s all-cause mortality estimates are more accurate than many previous studies. If so, this may be notable given the numerous biases potentially affecting mortality studies.^34^

### Potential “Quality Assessments” for bePE risk

By definition, exposure and nonexposure time periods are separate and distinct. Thus, since confounding can vary so much over time, no nonexposure risk metric can be expected to perfectly reflect confounding. Fortunately, a number of “quality assessments” can be envisioned that might identify instances in which a nonexposure risk metric may not correctly approximate confounding. Table 4 lists eight potential quality assessments for the bePE risk metric and includes their rationales and potential disadvantages. Two of the assessments actually can be expressed in two different forms, and other quality assessments can be easily envisioned (S13). Additional research may delineate which assessments, and threshold values for these assessments, are the most useful. Weighting results from multiple quality assessments may also prove useful.

**Table 4.**
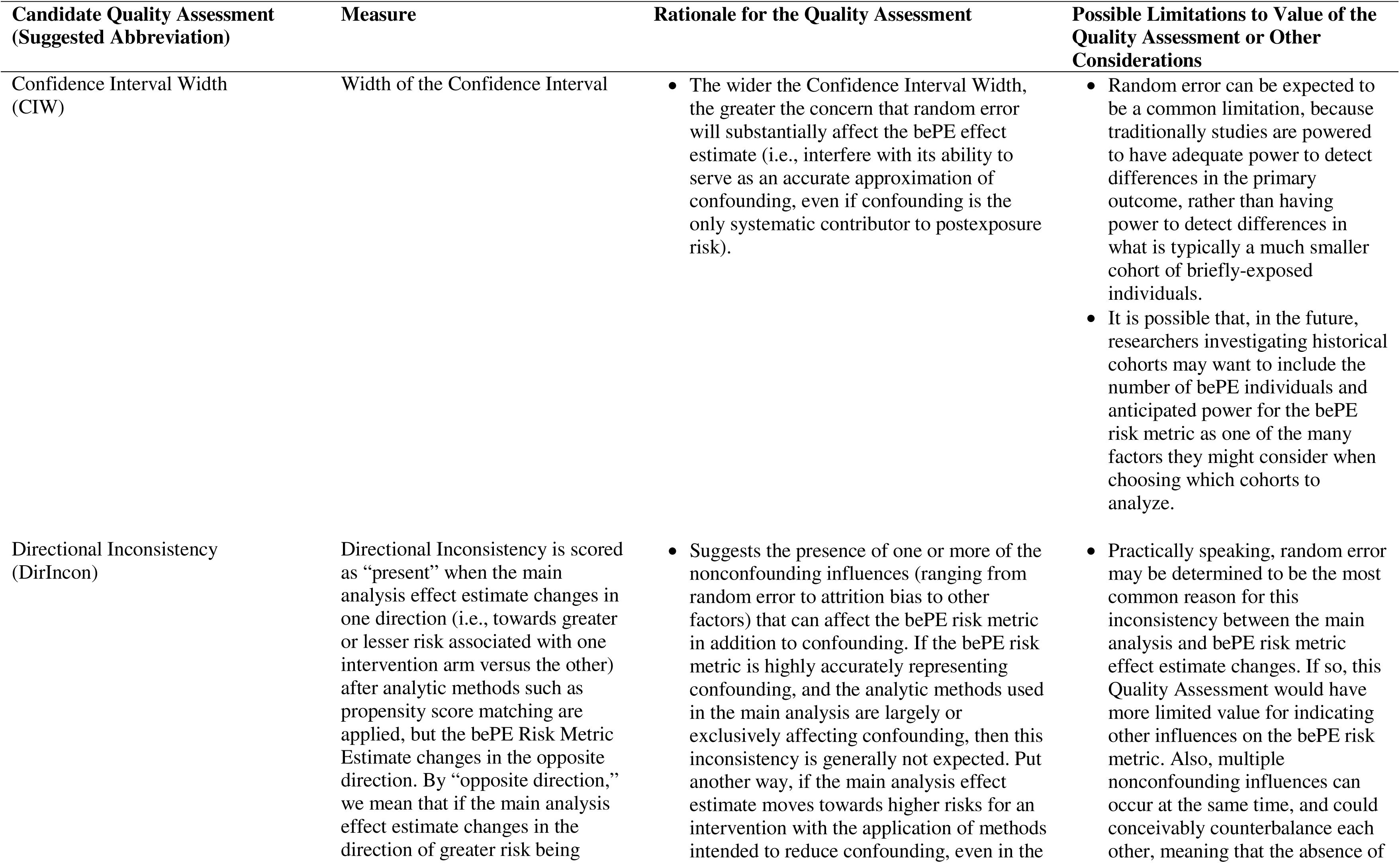
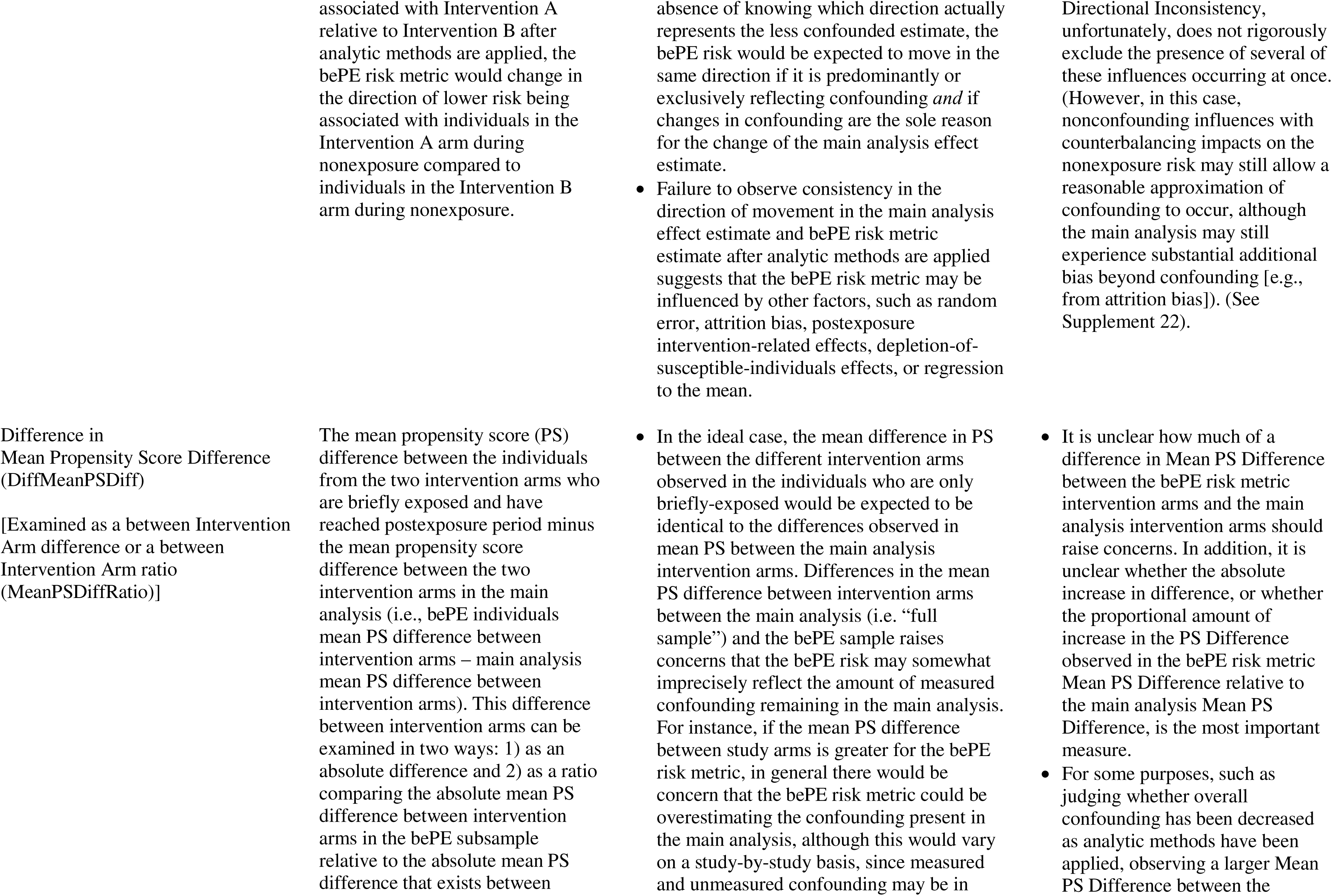
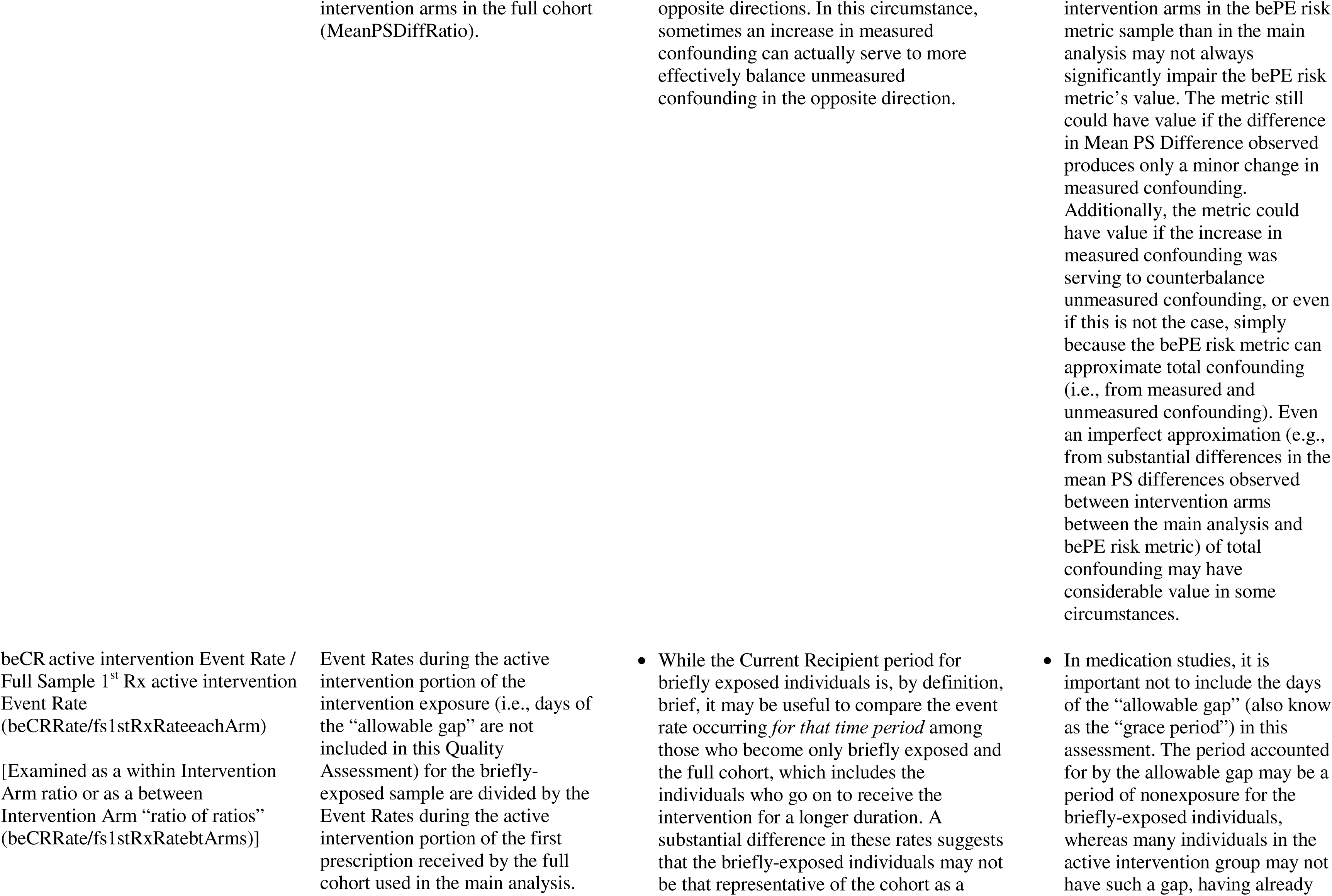
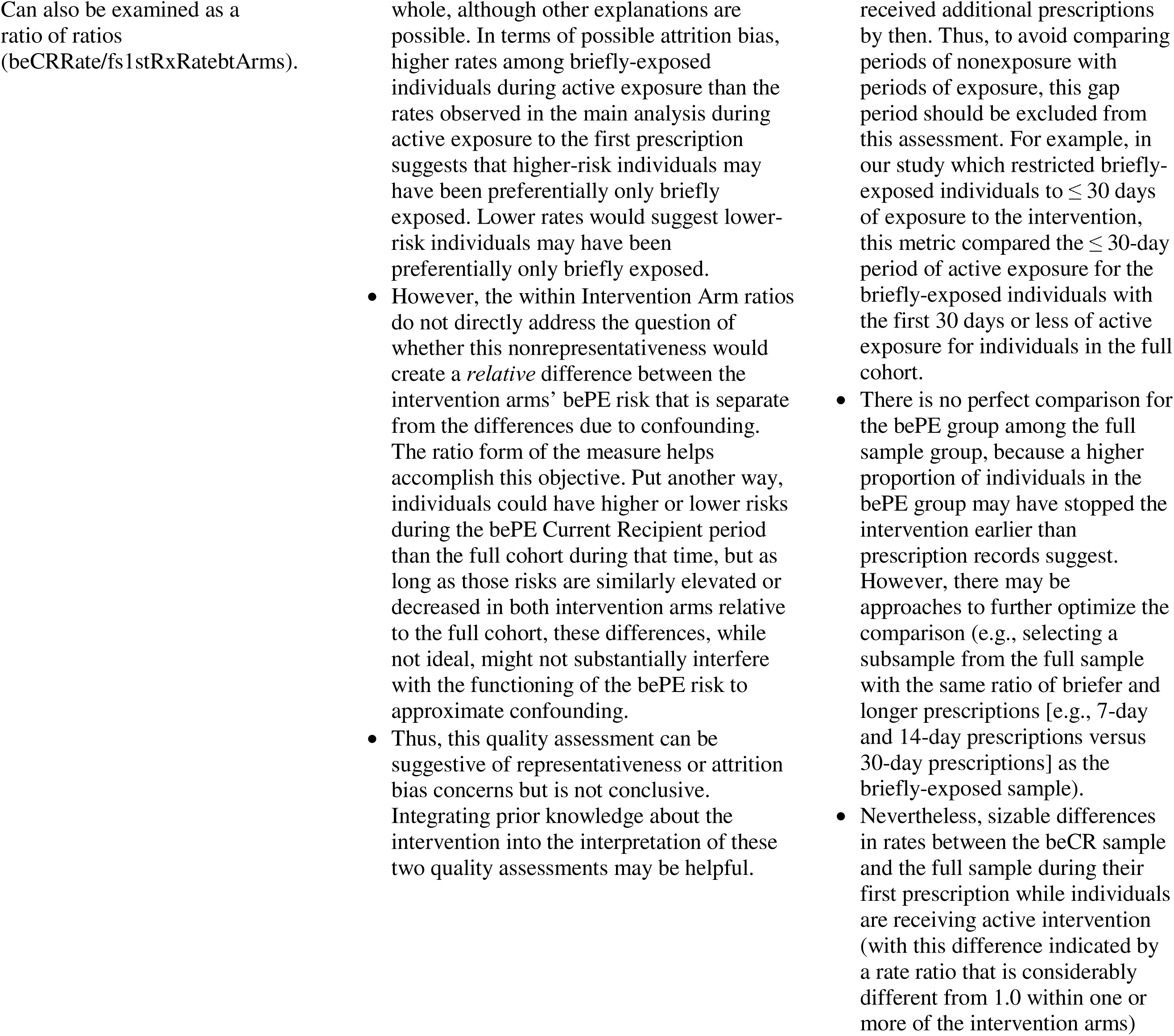
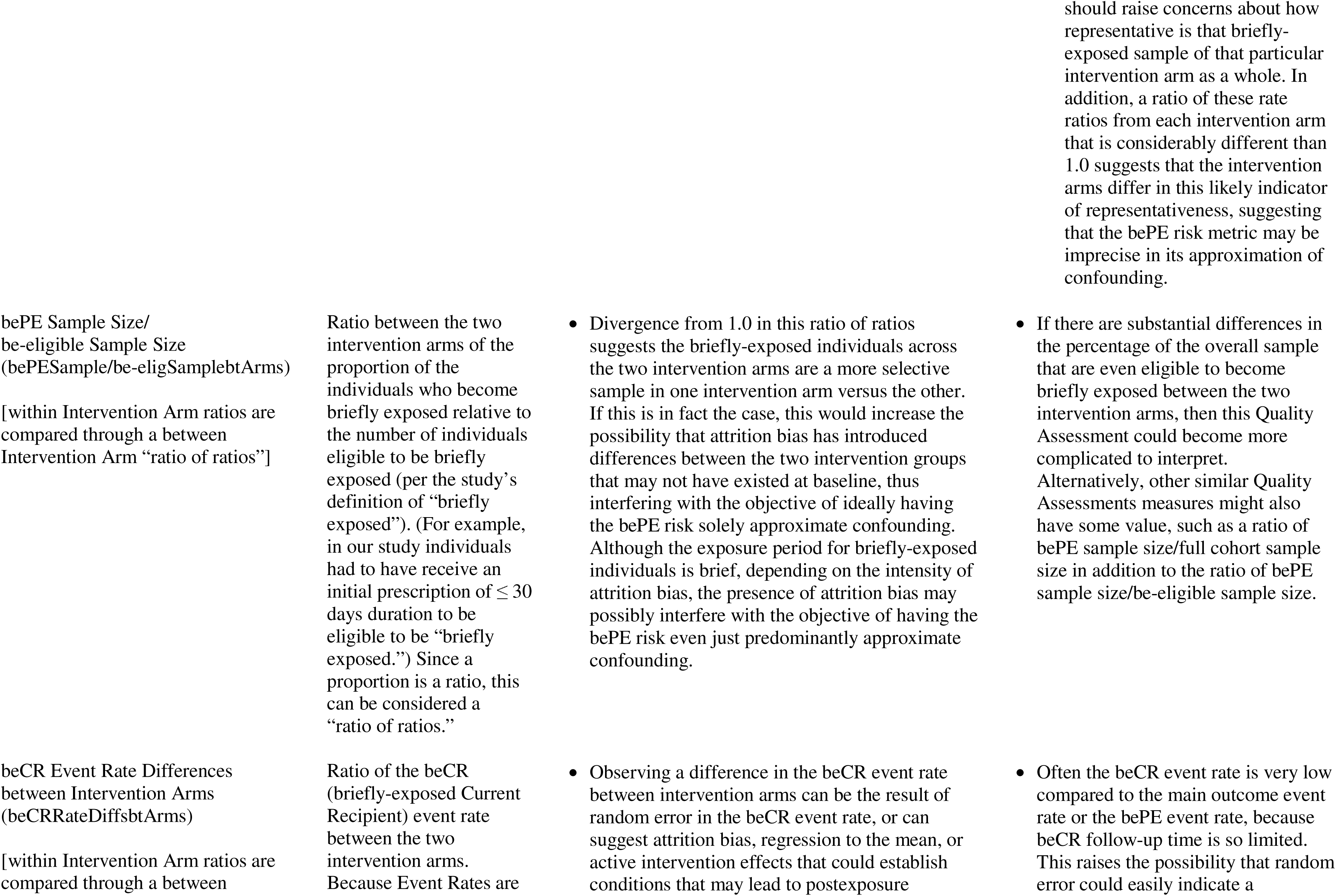
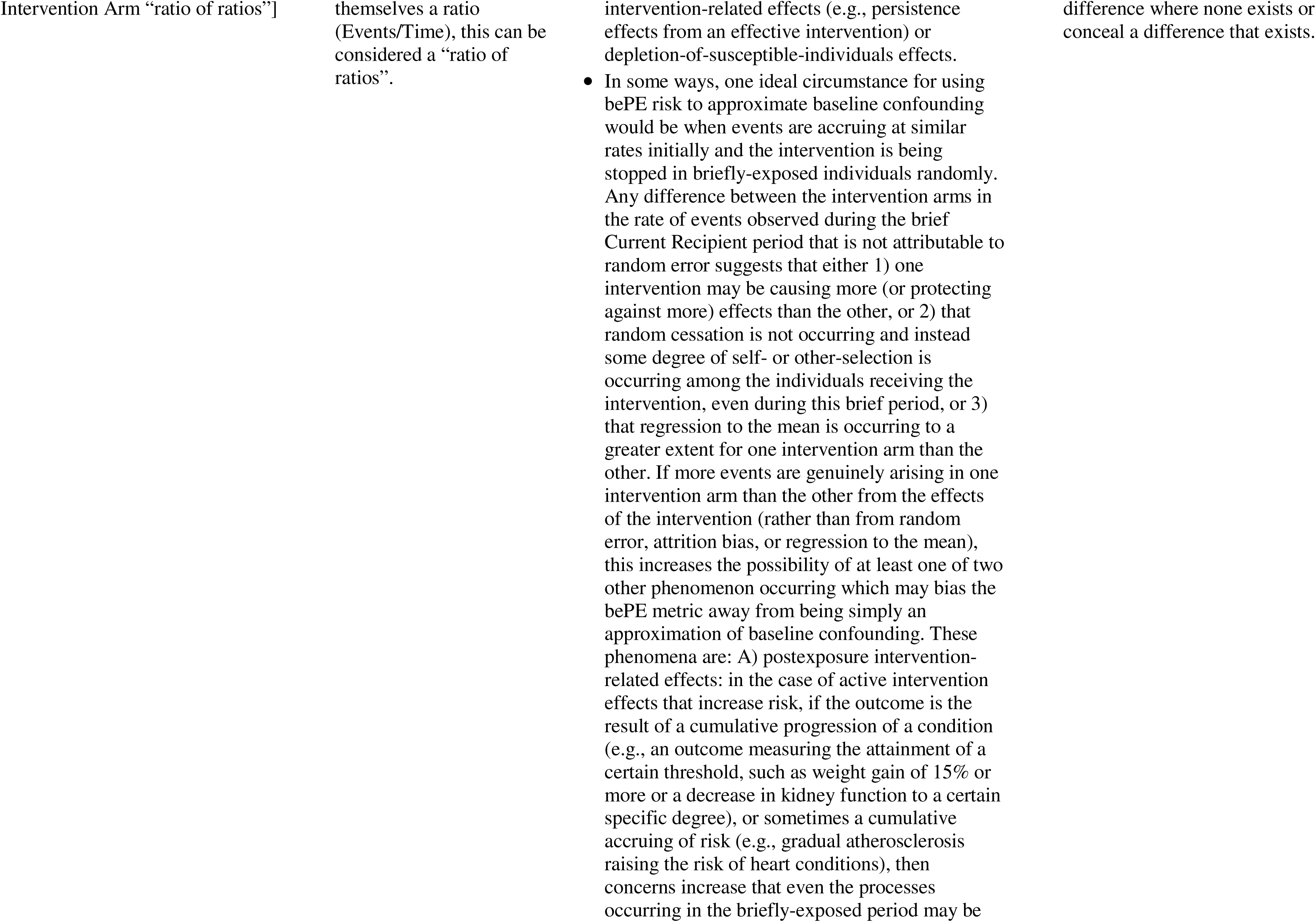
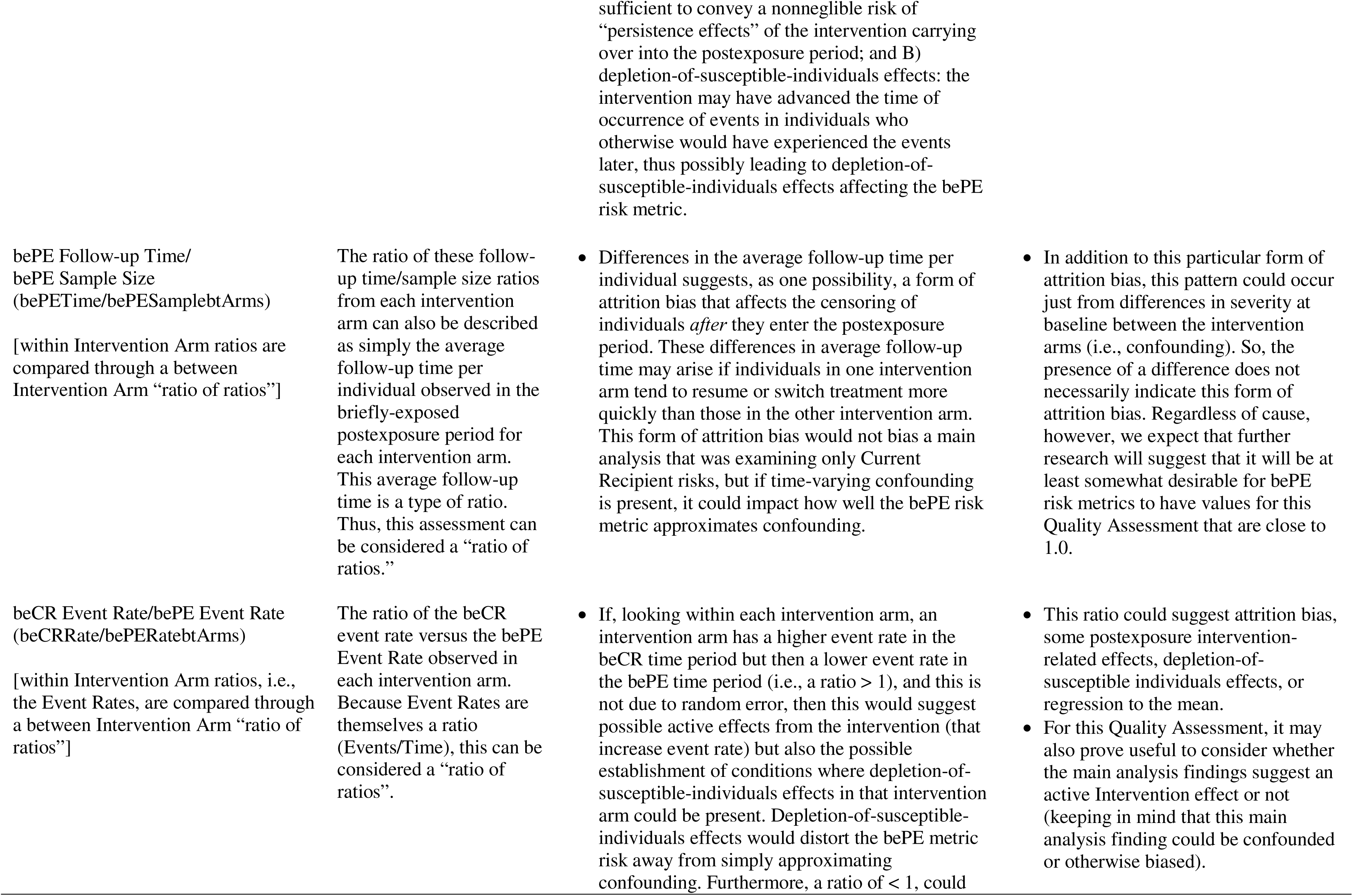
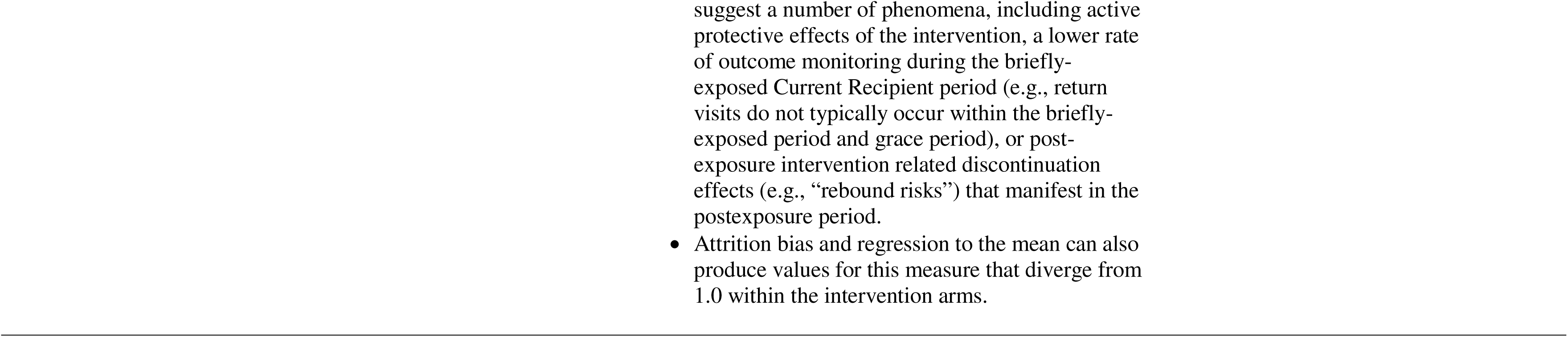
Candidate “Quality Assessments” suggesting whether Nonconfounding Influences on the bePE Risk may be Present that would Interfere with how well the bePE Risk Metric may Approximate Confounding.

### A Second Demonstration of the bePE Risk Metric, including Quality Assessments

Table 5 illustrates the application of the bePE risk metric to an analysis of medically-significant adverse effects associated with lithium and valproate, including ten different quality assessments. Cohort derivation was not optimized for each endpoint, but the selection of propensity score covariates was specific to each endpoint (S8). Over 365 days of follow-up, the central estimates for the bePE risk metrics for two of the three hdPS-matched analyses (Hypertension and ≥15% Weight Gain) are closer to the null (suggesting that these hdPS-matched analyses are less confounded than the unmatched analyses). However, this is not the case for the third hdPS-matched analysis (Hyperlipidemia). For this analysis, the bePE risk metric for the unmatched analysis is closer to the null (Hazard Ratio central estimate = 1.03) than the bePE risk metric for the hdPS-matched analysis (Hazard Ratio central estimate = 1.09).

**Table 5.**
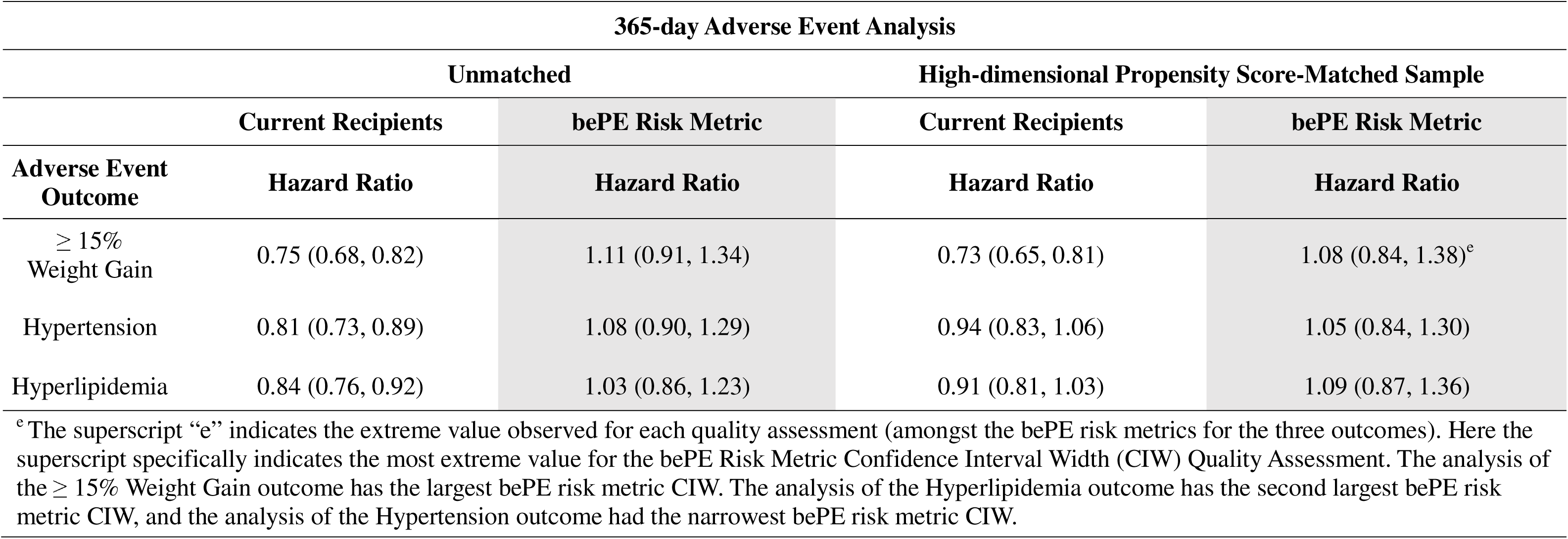
Application of the Briefly-exposed Postexposure (bePE) Risk Metric (Proposed Indicator of Approximate Confounding) to the Evaluation of Common Medically-Significant Adverse Events Associated with Lithium and Valproate, including Proposed Quality Assessments.

**Table 5B.**
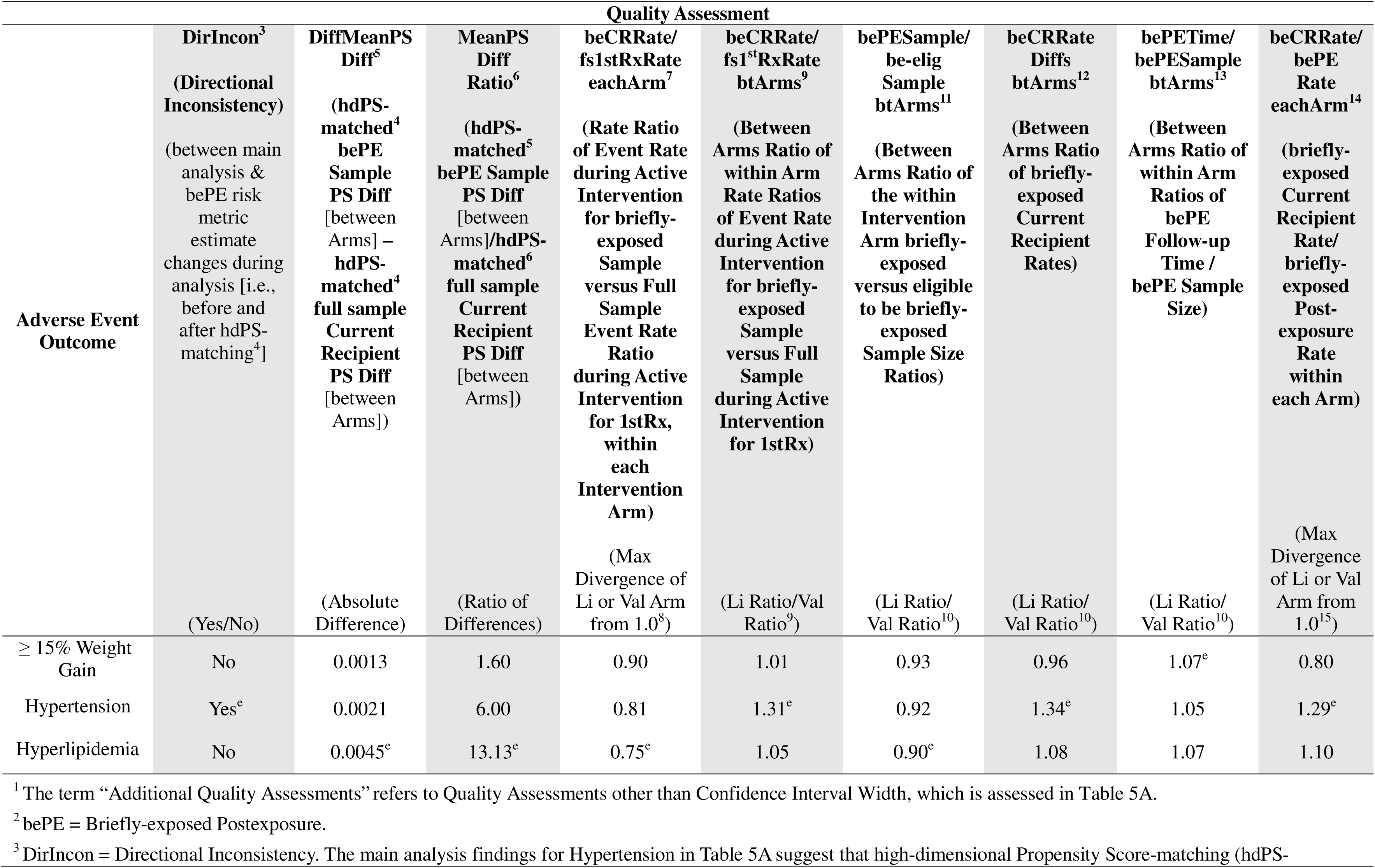
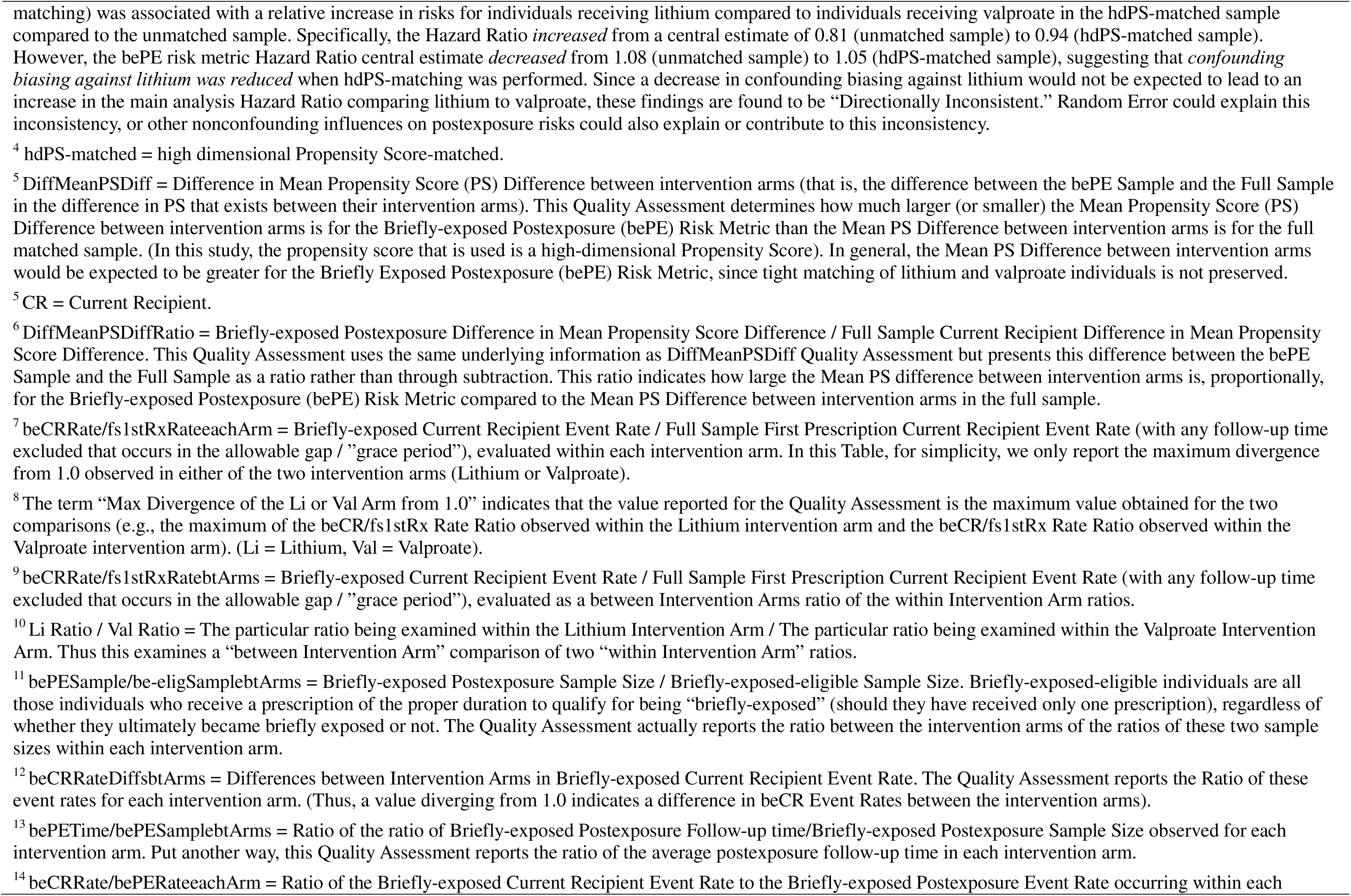
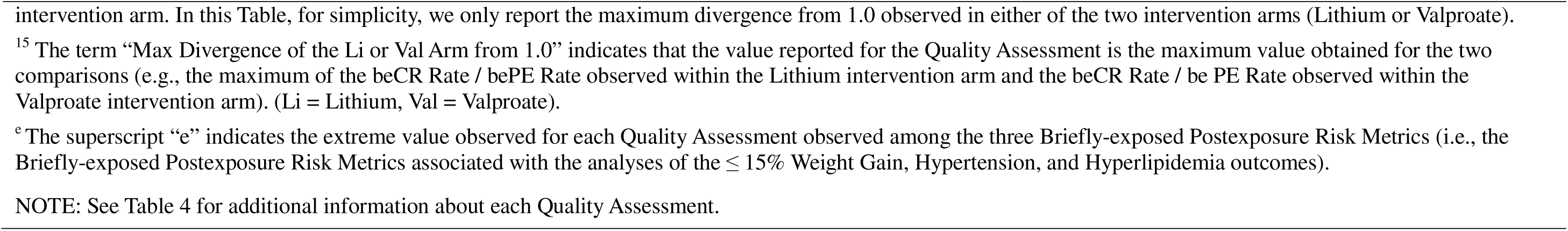
Findings from Additional Quality Assessments^1^ Relevant to Judgments Concerning How Well the Briefly-exposed Postexposure (bePE^2^) Risk Metric May Function as an Approximation of Confounding.

Random error, however, may substantially influence these bePE risk estimates, as suggested by the sizable width of the bePE risk metric’s confidence intervals for the hdPS-matched analyses (95% CI [0.87, 1.36]). In addition, the Hyperlipidemia analysis’ bePE risk metric was also associated with more extreme values for some, but not all, of the other quality assessments compared to the Hypertension and Weight Gain analyses. However, the significance of these differences is currently difficult to judge (S14).

For the Hyperlipidemia analysis, the larger divergence from the null observed for the hdPS-matched analysis bePE risks compared to the unmatched analysis might be explained in several ways. These include: 1) the unmatched analysis bePE risk metric estimates are simply not very different from the hdPS-matched analysis bePE risk metric estimates (increasing the potential impact of random error), 2) our hdPS contains more variables than some hdPS scores (thus it is possible that, despite our efforts, some problematic variables such as semi-instrumental variables^35^ or collider variables^35^ could have been inadvertently included), or 3) the selection of hdPS variables (based on a hazard ratio of >1.2^36^ with the forced inclusion of certain variables) may not perform as well as expected. Numerous other selection approaches exist,^37,38^ and researchers have observed that hdPS models using covariate selection strategies do not always outperform models that include all available covariates.^39^

Based on these results, one could consider reporting both the unmatched and hdPS-matched main effect estimates for the Hyperlipidemia analysis or highlighting the possible uncertainties for this particular analysis (such as the possible presence of confounding or the potential sizable random error in the bePE risk metric). Although the bePE risk metric needs careful interpretation, the mere existence of a metric with the potential to approximate confounding can help highlight which analytic findings may be more uncertain or warrant further inquiry.

### Limitations of Nonexposure Risk Metrics

As mentioned, nonexposure risk metrics typically assess risks only among subsets of the full cohort (except for some instances of the basePreE risk metric). In addition, since individuals are only examined while nonexposed, most nonexposure metrics do not index confounding over the entire time period under study in the main analysis. (However, by making the permitted intervention exposure as brief as feasible, this limitation can largely be minimized.) The sole exception is the abNE risk metric, which tracks risk across the entire study follow-up period but only involves those individuals who do not actually start the intervention. These forms of cohort nonrepresentativeness or incomplete representation of follow-up time mean that nonexposure risk metrics clearly are not perfect indicators of confounding for the entire cohort or period of exposure. For our proposed postexposure risk metrics (the bePE and ubePE risk metrics), another important challenge is that, for some outcomes, some degree of attrition bias, postexposure intervention-related effects, or depletion-of-susceptible-individuals effects could arise, even from only a very brief exposure to the intervention. For some outcomes, regression to the mean also could conceivably interfere with any nonexposure metrics’ ability to accurately approximate confounding. Some relatively convenient quality assessment measures, such as those we have proposed, may help indicate whether these other influences on nonexposure risk are present.

However, for the postexposure metrics, it is virtually impossible to completely rule out certain influences such as attrition bias (since it could be based on unmeasured factors) or some degree of postexposure intervention-related effects (S15).

The baseline preexposure risk metric can sometimes examine the entire sample (and thus achieve complete sample representativeness) and is not susceptible to attrition bias, postexposure intervention-related effects, or depletion-of-susceptible-individuals effects. However, this metric also has a number of limitations: 1) it does not measure risks over the actual time period of exposure for *any* of the study participants, 2) given the likely need to include weeks or months of time prior to intervention to examine enough events, this metric may inadequately capture highly time-varying confounding at the point of initiation (S16), and 3) it cannot be used in some important circumstances (e.g., for studies exclusively examining new-onset conditions, or mortality or other permanent conditions). For these reasons, we expect the choice of nonexposure risk metric(s) to vary on a study-by-study basis. Nevertheless, with appropriate use and interpretation, these nonexposure risk metrics should improve the quality of inferences from many intervention studies.

### Future Research Needs

This manuscript demonstrates the potential of the bePE risk metric to improve intervention studies. Furthermore, given the significance placed on deviations from the null, nonexposure risk metrics are, in general, consistent with the growing practice of using “negative controls” to assess confounding.^40–46^ This manuscript, however, only contains a few examples involving the bePE metric (and none involving the other proposed metrics). Even the examples provided do not evaluate the validity of bePE metric as rigorously as some other approaches might. There is a need to thoroughly test the bePE and other nonexposure risk metrics to determine how consistently they approximate confounding in intervention studies. This testing could be performed in simulated datasets (for which the unconfounded effect can be known) or large real-world studies examining frequent outcomes (to minimize random error). There is also a need to develop more quality assessments and to evaluate whether particular quality assessments, or a weighting of quality assessments, might reliably detect instances when nonexposure metrics inaccurately approximate confounding. Finally, the optimal technical specification of nonexposure risk metrics (S17) and their application in analyses featuring weighting or regression adjustment (S18) may need further investigation.

### Potential Future Applications

If supported by further research, bePE and other nonexposure risk metrics could help investigators decide upon methodology, both for specific studies and in general (e.g., assessing whether one approach consistently appears to more fully reduce confounding). Nonexposure risk metrics also could help inform what confidence should be given to specific findings within a study. These metrics may also help assess which studies should be given more weight when informally or formally (e.g., through meta-analysis) assessing a group of intervention studies.

In addition, one particularly valuable use of nonexposure risk metrics may be to directly assist other efforts to produce unconfounded estimates. For example, instrumental variables (IVs) are often difficult to validate. Nonexposure risk metrics could be used to at least partially assess whether employing a possible IV actually seems to produce a less confounded effect estimate (S19). Another potential innovation could be to leverage the basePreE risk metric to rapidly derive minimally confounded “surveillance-quality” effect estimates from relatively large, diverse samples (S20).

## Conclusions

This manuscript has described the overall rationale and key limitations of a new class of metrics intended to help approximate confounding, including confounding from unmeasured factors. In particular, it has served as a “demonstration of concept” for the bePE risk metric, which currently is almost certainly the most widely useful of our proposed nonexposure metrics. This new class of metrics expands and, in our view, markedly improves upon a basic approach of examining risks during nonexposure periods first used in 2001.^8^ This manuscript discusses the many potential validity threats that need to be considered when contemplating using nonexposure risks to approximate confounding and presents four newer, more specific metrics that minimize some of these threats. It also opens the investigation into approaches to assess the quality of these metrics.

We expect that our proposed nonexposure risk metrics will frequently serve as valuable quantitative approximations of confounding for intervention studies and certain other studies, given their ability to incorporate both measured and unmeasured confounding (S21, S22). Our demonstrations from a real-world study appear to support this expectation. Furthermore, three of our four proposed metrics approximate confounding using risks that are quantified from the same individuals (or a subset of the same individuals) who are also informing the main analysis effect estimates. Future research will need to determine whether this “internal” assessment of confounding has advantages that outweigh the fact that a number of nonconfounding influences can affect nonexposure risks. Ultimately, the value of nonexposure risk metrics may vary on a study-by-study, or even outcome-by-outcome, basis. Finally, in addition to informing individual studies, these tools may also help inform overall methodology development and meta-analyses.

While the benefits and limitations of these new metrics need more investigation, we expect that nonexposure risk metrics will help significantly improve the quality of many intervention studies and other epidemiological studies.

## Authorship Statement

<u>Eric G. Smith, MD, PhD, MPH</u>: Conceptualization, Methodology, Funding Acquisition, Supervision, Validation, Writing-Original Draft Preparation, Writing-Review and Editing Dr. Smith was the lead for the Conceptualization, Development, and Implementation of the briefly-exposed postexposure (bePE) risk metric as well as the other nonexposure risk metrics as presented in the manuscript, and the primary author of the manuscript. He also helped validate analytic findings and derived some of the Quality Assessment findings based on results provided by Dr. Netherton. Dr. Smith wrote all the Supplements other than Supplement 8 and co-wrote Supplement 8 with Dr. Netherton. <u>Dane M. Netherton, PhD</u>: Data Curation, Formal Analysis, Software, Validation, Writing-Review and Editing. Dr. Netherton was the lead (and only) data analyst and programmer throughout the project. He developed all the code to implement the bePE risk metric and performed all the cohort derivation and survival analyses presented in the manuscript (under the guidance of Dr. Smith). Dr. Netherton also performed all data curation including linking different datasets and evaluating the output of all models and debugging code, as well as designing and implementing the code to perform the internal data checks. Dr. Netherton derived some of the Quality Assessments reported in the manuscript. Dr. Netherton reviewed and edited all components of the manuscript. Dr. Netherton also helped validate analytic findings and co-wrote Supplement 8 with Dr. Smith.

## Data Availability

Data used for this manuscript are from Department of Veterans Affairs databases and associated databases (Center for Medicare/Medicaid Studies and VA/DoD Mortality Data Registry) that was gathered during the healthcare of veterans and is not publicly available.

## Acknowledgments

This study was directly supported by Independent Investigator-Initiated Research grants I01HX002794 and I01HX003453 from the United States (U.S.) Department of Veterans Affairs, Health Systems Research Service (HSR). (In addition, Dr. Smith’s previous work incorporating the all-discontinuations postexposure risk metric was supported by VA HSR&D Career Development Award 09-216.) This material is the result of work supported with resources and the use of facilities at the Center for Health Optimization and Implementation Research (CHOIR), a VA Health Systems Research (HSR)-funded Center of Excellence. The views expressed here do not reflect the views of the U.S. Department of Veteran Affairs or the United States Government.

We also acknowledge the support of the Veteran’s Administration VIReC’s VA/CMS Data for Research Project, SDR 02-237, for providing Veteran data obtained from the Center for Medicaid/Medicare Studies and/or the United States Renal Data System (USRDS), and to the Department of Veterans Affairs/Department of Defense Mortality Data Registry for providing mortality data.

Dr. Smith would like to particularly acknowledge and thank the following individuals: First and foremost, Brian C. Sauer, PhD, for a discussion more than 10 years ago about the limitations of the “all discontinuers” post-exposure risk that provided the genesis for the idea of focusing upon more briefly-exposed individuals. I believe that Brian was the first in our conversation to give explicit verbalization to the idea in both of our heads of assessing confounding using a measure restricted to individuals who had received only brief periods of exposure such as single prescriptions. My work on this approach in the intervening years owes a definite debt to this initial encouragement.

In addition, Katherine J. Hoggatt, PhD, MPH, has generously offered supportive statistical and epidemiological input (as well as other support) as needed for more than 10 years (and reviewed a draft of the manuscript). Any faults in this work, however, are those of the authors alone. I am also grateful to Marcia Valenstein, MD, who provided invaluable and essential support to my efforts to analyze lithium and valproate mortality risks using high-dimensional propensity scores, and whose critical judgment helped educate me on the general importance of considering and assessing, if possible, unmeasured confounding; Jeroan J. Allison, MD, MS, my PhD thesis advisor whose scientific curiosity and practical judgment helped deepen my thinking about my research; James F. Burgess, PhD (deceased) who helped reinforce the importance of evaluating unmeasured confounding and supported earlier efforts of mine to assess unmeasured confounding through the ACCE Method; Susan S. Jick, DSc, who unselfishly provided mentorship and support for my large database research interests at two different times in my research career; Steve M. Banks, PhD (deceased), an early mentor who strongly encouraged and supported my earliest efforts to understand and conduct propensity score analyses; Emelia J. Benjamin, MD, Scum, who helped spark my early interest in nonrandomized intervention studies and the challenges posed by confounding, and to C. Arden Pope III, PhD, a very early mentor who was extremely generous with his time as he educated me about epidemiology and cohort studies in the early 1990s and who has been an ongoing source of support and encouragement ever since then. In addition, I would like to thank the following individuals who reviewed multiple drafts of this manuscript, especially my wife Amy D. Borg, my brother David G. Smith, Martin P. Charns, DBA, and Angela M. Kyrish, MA. Ms. Kyrish also did invaluable work helping to prepare and proofread all documents, Tables, and Figures for submission (including helping to prepare Figure 1), along with the valuable help of Maura Slattery, MA and Shawn Dunlap, MA. I also wish to thank others who reviewed at least one draft of the manuscript: Sophie M. Bartels, PhD, MSPH, Ben Kragen, PhD, MBA, Mark S. Zocchi, PhD, MPH, Sarah L. Cutrona, MD, MPH, Allen L. Gifford, MD, Christopher J. Miller, PhD, Lawrence R. Karen Quigley, PhD, Talia R. B. Smith, and Nicholas J. Smith. Any errors, however, are those of the authors alone. I also want to thank all my directors, supervisors, and colleagues who supported my work at the Bedford VA Medical Center over the last 17 years, especially Dan Berlowitz, MD, MPH, Allen Gifford, MD, Barbara Bokhour, PhD, Sarah Cutrona, MD, MPH, Renda Weiner, MD, MPH, Donald Miller, DSc, Susan Eisen, PhD (deceased), Graeme Fincke, MD, and Lawrence R. Herz, MD, along with Janet E. Osterman, MD, my residency training director who supported my interest in research in multiple important ways. In addition, I want to thank the Max P. Oeschger, Charles Waechter, and Robert J. Maier research labs, where I first learned about scientific research, and the research program of K. Ranga Rama Krishnan, MBBS, where I first learned about medical research. Finally, I wish to especially thank my family for their love and support throughout this research that unfortunately too often occupied time that might have been spent with them. Their sacrifice is deeply appreciated.

