## Supplementary material for "Initial Description and Demonstration of a Family of Methods with the Potential to Approximate Confounding": Text Supplements (S1 - S22)

**for**

Provided by the Authors

*NOTE TO READERS: These “Supplements” are largely intended to provide clarifying information or potentially useful details that were not able to be included in the text. In a sense, they are generally intended to serve the same function as the “footnotes” or “endnotes” that are used in the manuscripts of some academic disciplines.*

*To facilitate ease of use, almost all of the Supplements contain a 2-4 sentence summary of the Supplement’s main point as the first paragraph. This summary is followed by more technical details or more detailed considerations in subsequent paragraphs that follow the statement “To elaborate:” An exception to this structure is Supplement 8, which provides details of our methodology to serve the same purpose as the “Methods” section typically included in a manuscript. We suggest that readers prioritize reading those Supplements that concern topics of particular interest to them. Furthermore, depending on their level of interest, we suggest readers then decide whether the first paragraph provides them with sufficient additional information or whether reviewing the entire Supplement would be helpful to them.*

*Some of the material in some of these Supplements describes supposition, speculation, or hypothesis, especially those parts of the Supplements that describe potential additional applications of these methods or additional research that could be done. We try to clearly delineate when we are engaging in speculation. We hope that these more speculative discussions of certain topics will allow researchers to benefit from our latest thinking (whether valuable or not) about these metrics. We include this material in the hope that it will speed the process of other researchers building on our manuscript to conduct research into nonexposure risk metrics.*

*ADDITIONAL NOTE (about the Terms used to refer to the Nonexposure Risk Metrics): For clarity, in these Supplements, we provide the full name of the nonexposure risk metrics each time we mention the metric. We expect that some individuals will prefer to read the full name for the metric rather than our abbreviations for the metric. However, other individuals may prefer reading the abbreviation, and in our manuscript we most often used the abbreviations rather than the metric names. Thus, we have decided in almost all cases, except the Supplement headers, to use both the full name and the abbreviation to refer to a nonexposure risk metric (e.g., the “briefly-exposed postexposure [bePE] risk metric). We hope that this usage is not distracting or appears unnecessarily redundant to readers. We also hope it does not imply that we believe that, in the future, the full name and abbreviation should both be used to refer to these metrics each time they are mentioned. Rather, we hope that, over time, using the abbreviations alone will become usual practice in most instances.*

**Table of Contents**

Supplement 1. Clarification of the term “nonexposure risk” and of some other terms used in the manuscript

Supplement 2. A brief summary of prior use of the nonexposure risk concept

Supplement 3. Substantial limitations exist to using “all discontinuations” postexposure (adPE) risk as an approximation of confounding

Supplement 4. Possible influence of depletion-of-susceptible-individuals effects on nonexposure risk metrics

Supplement 5. Incomplete representation of follow-up time

Supplement 6. Further thoughts about potential influences on nonexposure risk metrics, especially regression to the mean

Supplement 7. The assigned-but-not-exposed risk metric and ultra-briefly-exposed risk metric are likely to have particularly pronounced random error

Supplement 8 . Methods used in the real-world data demonstrations of the briefly-exposed postexposure (bePE) risk metric

Supplement 9. Rationale for the use of “modified” or “additionally-censored” intent-to-treat analysis censoring conditions

Supplement 10. Alternative definitions of nonexposure periods can be considered that vary based on the stringency with which follow-up time on other interventions are excluded from the metric

Supplement 11. Clarification of the purpose of using a nonexposure risk metric during cohort derivation

Supplement 12. Further consideration of why using the briefly-exposed postexposure (bePE) risk metrics to aid cohort derivation is not expected to allow additional biases to affect the main analyses

Supplement 13. Additional quality assessments and other measures that could be considered to assess nonexposure risk metrics

Supplement 14. Further discussion of the Hyperlipidemia analysis briefly-exposed postexposure (bePE) risk metric quality assessment findings

Supplement 15. A potential sensitivity analysis to assess for possible attribution bias, postexposure intervention-related effects, and/or depletion-of-susceptible-individuals effects

Supplement 16. Considerations that are relevant to concerns that the baseline preexposure risk measure may not precisely capture confounding at the point of intervention initiation

Supplement 17. The optimal form for the briefly-exposed postexposure (bePE) risk metric effect measure (and other nonexposure risk metrics effect measures) may need exploration

Supplement 18. Suggested approaches to using the briefly-exposed postexposure (bePE) risk metric in analyses including regression covariates or weighting approaches

Supplement 19. Use of nonexposure metrics to validate instrumental variables

Supplement 20. The baseline preexposure risk metric may be able to be leveraged to rapidly derive “surveillance-quality” effect estimates from large, diverse samples

Supplement 21. A rationale to examine multiple nonexposure risk metrics when feasible

Supplement 22. Additional thoughts about the value of our proposed nonexposure metrics

**Supplement 1.** **Clarification of the term “nonexposure risk” and of some other terms used in the manuscript**

We intend the term “nonexposure risk” to refer to time-limited periods of nonexposure experienced by some (or, for the baseline preexposure [basePreE] risk metric, sometimes all) of the individuals who also receive the exposure (except for the one metric that examines individuals who are not exposed at all to the intervention).

To elaborate: To clarify why we are using the term “nonexposure” rather than “nonexposed” or “unexposed,” to us “nonexposure” suggests that *periods* of nonexposure, in addition to periods of exposure, can occur. In contrast, to us at least, “unexposed” (and, unfortunately, the term “nonexposed,” which is highly similar to our preferred term of “nonexposure”) tends to suggest that exposure *never* occurred, at least within the time frame in question. These differences are quite subtle and also quite likely not to be universally shared. We also acknowledge that the term “nonexposure” may sound slightly less grammatical or more awkward than “nonexposed.” However, we reiterate that we favor this term because of the concern that “nonexposed” might be read as designating individuals who are either never exposed at any point or never exposed during the period of the study. In contrast, to us “nonexposure” seems to make the point at least somewhat clearer that these individuals could have just had an exposure or be about to have an exposure. We welcome suggestions for improvements on this terminology that may be even less ambiguous than this approach.

Technically speaking by this usage, the assigned-but-not-exposed metric is not a “nonexposure” metric, but an “unexposed” metric, since it examines people who are completely unexposed to the intervention or agent studied (at least for the time period studied). However, for convenience, we continue to use the term “nonexposure risk metric” to refer to this metric as well.

Also, at times we may use the more specific term “postexposure” risks to refer to the subset of nonexposure metrics that evaluate risks after exposure to the study (i.e., the briefly-exposed or ultra-briefly-exposed postexposure (bePE) risk metrics). These metrics have some common limitations (e.g., the possibility of attrition bias, postexposure intervention-related effects, or depletion-of-susceptible-individuals interfering with these metrics) that are not shared with the assigned-but-not-exposed (abNE) and baseline pre-exposure (basePreE) risk metrics, hence the value of using the “umbrella” term to refer to these two metrics (or three metrics, if the “all discontinuations” postexposure (adPE) risk metric is also being referenced).

To clarify our use of several other terms, here is a description of how we use some of the other terms referred to in our manuscript:

1) We use the broader term “confounding” to apply to the confounding remaining in the analysis, which can also be termed “residual confounding.” We favor the simpler term “confounding” and use it exclusively, even when we are discussing the confounding remaining in the analysis. We use this simpler term in order to use less “jargon” and improve the readability of our manuscript.
2) We use the term “approximate” as in “to approximate confounding” or “approximating confounding” to replace what we believe to be the more typical term “estimate confounding.” We make this distinction because of the large number of factors that can affect nonexposure risk metrics and the difficulties in conclusively eliminating all of these factors as potential contributors to observed nonexposure risk. These nonconfounding influences can especially affect postexposure nonexposure risks, ranging from attrition bias, postexposure intervention-related events, depletion-of-susceptible-individuals effects, regression to the mean, random error, and possibly others, such as change in risk factors or initiation of new interventions during the postexposure period (see Supplement 22). To us, given these additional uncertainties besides random error, it did not seem proper to use the term “estimate,” which can also be applied to values that are mostly or solely affected by random error. Of course, any approximation can, in the common sense of the word, be termed an “estimate.” On the other hand, many “estimates,” such as effect estimates from regression models, can be highly dependent on assumptions, and therefor may also have many sources of uncertainty. Thus, we may be “outliers” in our preference for using the term “approximate” or “approximation” in this work on quantitatively assessing confounding, rather than the term “estimate.” However, we will note that our usage of the term “approximate” appears to be highly congruent with language used by Lipsitch and colleagues when discussing “negative controls”^1^ for assessing confounding. For instance, these authors indicate that the comparability between an exposure and a negative control exposure will be “only approximate.”^1^

We leave open for discussion by the research community whether separate terms, such as how we are using the terms “estimate” and “approximate,” should be used for quantities that are thought to vary largely or entirely due to random error versus quantities that are potentially sensitive to substantial influences besides simply random error.

3) In addition, in case it is not clear, we use the term “central estimate” to refer to the mean effect from our analyses (i.e., the “central” value that is flanked by the upper and lower confidence limits).
4) We use the term “current recipient” to replace the term “current user” that we believe has been used most often in the past to refer to individuals (in medication studies) who are currently receiving or using the active intervention. We favor the term “current recipient” because it helps highlight, or at least note, that in database studies can be very difficult to determine conclusively whether individuals are “using” a particular medication or intervention, but it is often much less difficult to determine if they have at least received the intervention. Using the term “current recipient” helps us at least remember that there are some uncertainties about whether individuals are actually using the intervention that they are receiving.
5) We use the term “main analysis” to refer to standard analyses that involve the entire sample under study for that particular analysis. Additionally, during the main analysis this sample is actively receiving the intervention (except for any ‘grace” period) during all (i.e., a current recipient analysis) or part (i.e., an intent-to-treat or modified intent-to-treat analysis) of the follow-up period.
6) We use the term “nonconfounding influences” to refer to other processes or phenomena that can affect nonexposure risks (e.g., see Supplements 3 and 22). These influences can include random error as well as systematic influences such as attrition bias, postexposure intervention-related effects, depletion-of-susceptible-individuals effects, etc.
7) We use the term “postexposure intervention-related effects” to discuss effects from exposure to the intervention that manifest during the postexposure period. These can include persistence effects (i.e., persistent benefits or risks from the intervention), or risks or benefits associated with discontinuation (which can also be termed “rebound effects”). The reason we use such a lengthy term, rather than simply “postexposure effects,” is because other factors besides the intervention can have effects in the postexposure period. In addition, “postexposure effects” could even be read by some as indicating effects that the postexposure period has on other risks, or risks even subsequent to the postexposure period. So, for the sake of clarity, we use the term “postexposure intervention-related effects” to specifically refer to effects from the intervention that are observed in the postexposure period. The one exception is in the Abstract, where, to reduce jargon at the earliest part of the manuscript, we simply say “postexposure effects.”
8) We use the term “intervention studies” to refer to studies that make deliberate, intentional changes to one group compared to another, including nonrandomized and randomized studies. However, although the briefly-exposed postexposure (bePE) risk metric and other nonexposure risk metrics can be applied to randomized as well as nonrandomized intervention studies, it is unclear whether the need for them will be as great. Most randomized trials that are large enough to have enough participants informing the nonexposure risk metrics to give reliable estimates of nonexposure risk are probably going to be large enough to have done an adequate to very good job of addressing confounding already. However, there may be a role for the nonexposure risk metrics in helping to confirm that a large randomized trial does not have a confounded sample from some systematic study flaw (e.g., inconsistent procedures across sites in a multi-site study). There may also be a role for nonexposure metrics in evaluating small or medium-sized randomized trials for residual confounding, although statistical variability in the nonexposure risk estimates may be too large to make this approach practical. It should also be noted, as is noted in the manuscript, that the baseline preexposure (basePreE) metric is the one nonexposure metric for which the sample size may, in certain instances, be as large as the entire cohort under study. Thus, there may be instances when this nonexposure risk metric could be of value for assessing trials, especially small or medium trials, if outcome information has been systematically acquired for the period prior to trial enrollment. However, to be very useful the baseline preexposure (basePreE) metric would need to have enough power to detect what are likely to be modest differences in confounding. It is unclear how often this power would be expected to be present for small- to medium-sized trials.
9) We use the term ”full cohort” or “full sample” to refer to the final analytic sample being evaluated in the main analysis. Since the real-world demonstrations of the bePE risk metric in our manuscript primarily involve examples from high-dimensional propensity score-matched analyses, this term most often refers to the full sample of individuals who have successfully been matched using high-dimensional propensity scores. To be specific, this “full sample” or “full cohort” are the individuals remaining in the cohort after both propensity score trimming to the “Common Support Area” and matching has occurred. (To reinforce this point, individuals who are not matched or who fall outside of the Common Support Area are not included in this “full sample”). We may use this “full sample” or “full cohort” term occasionally to refer to the Unmatched cohort, if we are discussing the bePE risk metric that is associated with the Unmatched cohort. We should clarify that the “Unmatched” cohort has also not undergone any propensity score trimming, thus a more precise name would be the “Untrimmed and Unmatched” cohort.

To be clear, the term “full sample” or “full cohort” never refers to the “Total Recipients” sample noted at the top of our Cohort Derivation Flowchart (Manuscript Table 2), or any other sample from any other step in the Cohort Derivation. This term just refers to the sample from the final step of our cohort derivation (the “Unmatched” sample) or the hdPS-matched sample derived from this sample. In most cases, it refers to the hdPS-matched sample. We hope that which of the samples is being referred to by the term will be obvious, given the context provided by the sentence or overall discussion occurring when the term is used.

10) Finally, in the manuscript we use the term “some other studies,” as in the phrase “intervention studies and some other studies that compare groups of individuals” to refer to certain types of nonintervention studies which are potential candidates to employ nonexposure risk metrics. What we principally have in mind are studies of exposures to substances or things, such as in environmental or occupational epidemiology. If those studies involve two or more groups being exposed, and outcome data is compiled either before or after exposure (in addition to during exposure), then it seems to us that they are candidate studies for applying one of our proposed nonexposure risk metrics. In our experience, not all studies of exposures incorporate outcome assessment in the period before or after exposure is over. Therefore, these metrics would only be useful for studies that have included such measurement.

The need to have continued assessment of outcomes either before or after exposure is over is one reason why these methods seem particularly well suited to database studies. In many databases, outcome data is being continually compiled, regardless of whether an individual is receiving an intervention or experiencing exposure to an agent of interest.

**Supplement 2.** **A brief summary of prior use of the nonexposure risk concept**

When viewed in the strictest sense, we have found few very similar antecedents to the methods proposed here, which specifically involve examining periods of nonexposure in *some or all of the same individuals who are also exposed to the intervention.* In fact, we have only identified two clear antecedents: 1) an approach examining postexposure risk that was used at least a few times from 2001 - 2011 that is highly comparable in some ways to our “all discontinuations” postexposure (adPE) risk metric,^2-6^ and 2) the use of preexposure periods in self-controlled case series and some other studies as an antecedent for the baseline preexposure (basePreE) risk metric.^7-9^ (Sometimes self-controlled case series also examine postexposure periods in a manner seemingly closely similar to the “all discontinuations” postexposure (adPE) risk metric).^7^ Despite these few close antecedents, it should be noted that the approaches that we propose are generally consistent with the well-established approaches for evaluating confounding through the use of “negative outcome controls” and “negative exposure controls.”^1^

To elaborate: In general, our proposed approaches involve examining periods of nonexposure in some or all of the same individuals who are also exposed to the intervention (with the sole exception of the assigned-but-not-exposed metric). In fact, the exposed individuals that make up the briefly-exposed postexposure (bePE), ultra-briefly-exposed postexposure (ubePE), or baseline preexposure (basePreE) risk metrics actually make up part of the main analyses effect estimates (in addition to informing the nonexposure risk metric). Furthermore, in our proposed approaches, these nonexposure risks between intervention groups are compared in studies that involve two or more cohorts of individuals that are followed over time. (By “cohorts of individuals” we mean to include traditional nonrandomized “cohort studies” and randomized studies, but not alternative designs that do not involve full cohorts, such as case-control studies.)

The most obvious antecedent to the methods we are proposing, as mentioned in the manuscript, is the approach that we have termed the “all discontinuations” nonexposure risk metric. (This term was not used by the original authors. We have coined this term in order to differentiate this approach from the approaches that we are proposing, and to facilitate discussion about the strengths and limitations of each approach.) The approach that we are referring to with the term the “all discontinuations” postexposure (adPE) risk metric has been applied very occasionally to cohort studies, we believe, since 2001.^2-6^ (The year 2001 is the earliest use of a form of the “all discontinuations” postexposure (adPE) risk metric that we could identify in a PubMed search by using the term “former users” that was used by investigators at the time. A PubMed search that included the terms “former user” as well as “former users” or “past user” or “past users” revealed no earlier use of this approach in randomized or nonrandomized cohort studies that we could detect.)

In 2001, Nielsen, Sorensen and others used former users as a control for confounding, with former users in their study being defined in a manner that is highly similar to what we are terming the “all discontinuations” postexposure (adPE) risk metric.^2^ In their study of the association between current use of corticosteroids and risk of hospitalization from upper gastrointestinal bleeding, “current use” of these typically transiently-used medications was described as the date of prescription until 50 days later or until censoring.^2^ Former use of corticosteroids then extended from 51 days after a prescription until the date of the next prescription or censoring.

In their paper, the authors make it clear that they viewed former users, thus defined, as having value as a representation of confounding. They found that former users had a risk for adverse outcomes that was intermediate between current users and nonusers. The authors interpreted this finding as suggesting “that confounding, probably by indication for corticosteroids use or by severity of underlying diseases, accounts for at least part of the increased risk [that they observed as associated with the current use of corticosteroids in their study].”^2^ Since all individuals discontinuing the medication during the time period under study were included in this former users group, we would consider this one form of an “all discontinuations” postexposure (adPE) risk metric.^2^ There are some differences, however, from what we would view as a more typical “all discontinuations” postexposure (adPE) risk metric: 1) all individuals had a uniform initiation date for the start of their post-exposure risk, and 2) former users could reenter the current user cohort if they received a subsequent glucocorticoid prescription, and even apparently reenter the former use cohort 51 days after that next medication reinitiation date.

It should be noted that in this Nielsen et al. 2001 study^2^, their former user definitions, while more closely following the “all discontinuations” risk metric, also begins to approach our “briefly-exposed” postexposure risk metric definition. Nielsen and colleagues started their follow-up for former users relatively soon after the initial prescription date. As a result, their former users follow-up began on day 51 of follow-up (from the intervention initiation date) for all individuals, while follow-up for the briefly-exposed postexposure (bePE) risk metric as implemented in the lithium and valproate study in our manuscript began, on average, on day 45 or 46 of follow-up (from the intervention initiation date). (See Supplement 5 for more detail). Nevertheless, to fully match our recommendations for the briefly-exposed postexposure (bePE) risk metric, only users of single prescriptions would be considered, whereas it appears individuals receiving multiple prescriptions could be included in the Nielsen et al. study.^2^ In addition, our approach does not allow patients to become current users or former users a second time during follow-up (see manuscript and Supplement 8).

However, while the *definition* that Nielsen and colleagues used for former users is similar to our definition of the “all discontinuations” post exposure risk metric, the i*mplementation* of definition in the study differs in some important ways from what we refer to as the “all discontinuations” postexposure (adPE) risk metric. Most importantly, Nielsen and colleagues do not compare the risks associated with former users across two or more intervention groups. (That is, they do not observe the risk observed among former users across intervention arms during the period in which the individuals are no longer receiving the intervention). Rather, in their study, they apparently only compare current users of corticosteroids to former users of corticosteroids, even though they were also examining other types of medication in their study.

We suspect that there may be specific drawbacks to the practice of simply comparing current users to former users in a single intervention arm design, rather than in some sort of multiple intervention arm design in which former users in one intervention arm are compared to former users in the other intervention arm(s). In particular, we are concerned that the issue of how well the former user cohort represents the current user cohort may become even more important. This heightened concern arises from the fact that individuals discontinuing an intervention at any given time might differ more from individuals remaining on the intervention than from individuals discontinuing a similar intervention. The comparison between individuals who are postexposure from different intervention arms would be expected in some instances to potentially reduce concerns about factors such individuals’ impulsivity or reluctance to implement an intervention, if these factors were similarly present in individuals discontinuing each intervention. That is, this approach of comparing nonexposure risks across intervention arms may reduce concerns about unrepresentativeness, at least in some cases, because the individuals in the other intervention arm may also possess these characteristics to a greater or lesser extent. Interestingly, examining postexposure risks across intervention arms may be even more important for a metric that examines individuals who are only exposed briefly or extremely briefly to the intervention than for studies and approaches in which almost all the individuals who are exposed to the intervention contribute to the nonexposure risk estimate. We highlight this point because when nonexposure risks from only briefly or extremely briefly exposed individuals are examined, concerns about factors such as impulsivity and health-seeking behaviors are more prominent. Thus, for briefly-exposed postexposure (bePE), ultra-briefly-exposed (ubePE), and assigned-but-not-exposed risk metrics, comparing postexposure risks across intervention arms may be particularly important.

We suspect that, in general, it will often be preferable to compare postexposure risks across two or more intervention arms, rather than comparing postexposure risks to the risks observed during active exposure to the intervention in a single arm design. However, this supposition is speculation and the magnitude of difference in the estimates or conclusions that would result estimates between the two approaches is uncertain. The advantages and disadvantages of assessing postexposure risk across two or more arms of an intervention study, rather than in reference to current recipients in the same intervention arm, are certainly a potential topic for additional research.

In another interesting wrinkle, Nielsen and colleagues actually presented a subanalysis of their results for their current users as risks relative to the risks observed amongst former users, which reduced the risks from 2.9 (95% CI 2.2 to 3.7) to 1.9 (95% CI 1.4 to 2.5). In our manuscript we have used the briefly-exposed postexposure (bePE) risk metric as an *indicator* of the approximate amount of confounding, rather than *directly adjusting* the main effect analysis finding by the value of the nonexposure risk metric. (That is, we do not divide the effect estimate for the main analysis by the effect estimate for the nonexposure risk metric). Our impression is that using the nonexposure risk metrics directly as adjustment factors for the main analysis factor would risk amplifying the potential impacts of uncertainties in how accurate an approximation the nonexposure risk metric is of confounding. However, our perspective also involves some speculation, and we have not evaluated this question formally. The approach of using nonexposure risk metrics to directly adjust the main effect estimates could be a research topic to consider in the future.

Another research group that has used a form of “all discontinuations” postexposure (adPE) risk metric is the research group of Wayne Ray and colleagues.^3-6^ Their first use of former users in a fashion that yields an “all discontinuations” postexposure (adPE) risk metric was almost simultaneous with Nielsen and colleagues in 2002 (i.e., their paper was published just two months later).^3^ Their manuscript does not contain much detail about their “former users,” but it appears that their non-aspirin, non-steroidal anti-inflammatory medication (NANSAIDS)-receiving cohort was defined at the point of initiating NANSAIDS. Thus, the “former use” they examined (“no use on that day”) appear to always have followed a period of NANSAID exposure among individuals who were also considered current users earlier in the same study. Of note, their study did not involve two intervention groups being compared, but rather NANSAID recipients and a matched, unexposed cohort, and former users were only evaluated for the NANSAID recipient group and not the unexposed cohort. Interestingly, the duration of the former user period was also apparently capped at 365 days, while apparently NANSAID exposure could last as long as five years. (Judging from the amount of follow-up time, however, it appears most “current use” of NANSAID treatment were for only short durations, given that 181,441 individuals had only 65,502 person-years of “current use,” but provided 210,063 person years of “former use.”^3^ Therefore the fact that the study design established conditions under which a greater amount of follow-up time might be present for current users than former users appears to be more of theoretical than actual importance.) However, similar to the Nielsen et al. 2001 study,^2^ individuals were apparently allowed to become current users and then former users multiple times. (The investigators did perform an analysis restricted to the first current user follow-up period to assess the effect of cohort reentry, but this analysis appeared to literally only include the first current user follow-up period, thus an analysis of the first former user period was apparently either not conducted or not reported).^3^ Of note, the investigators specifically commented on the fact that their “former use” cohort exhibited risks that were close to those observed in the unexposed control group (former use had an adjusted rate ratio of 1.02 [95% CI 0.97-1.08], with the unexposed control group being used as the reference group). The authors suggested that this close similarity in risk between the former users and the unexposed control group indicated that “residual confounding by behavioral or lifestyle factors is possible, [but] the fact that the risk for former non-current users of NANSAIDs was virtually identical to that of non-users suggests that the size of such confounding is not large.”^3^

The Ray group subsequently used former users risks in a similar manner in 2004,^4^ 2009,^5^ and 2011^6^ as well. In these instances as well, the Ray group also discusses, to a greater or lesser extent, the fact that they view these former user analyses as helping to address concerns about confounding. For instance, in 2004 the Ray group specifically mentioned that former users of the study medication should be similar to current users “with regard to potential risk factors” that are “difficult to measure.”^4^ (We note that the Ray group also apparently reports former user risks defined in the same manner as the 2002,^3^ 2004,^4^ 2009,^5^ and 2011^6^ studies in another study in 2009,^10^ but do not comment about the use of those findings to address confounding or to aid interpretation of the main effect analyses.)

In one sense, the Ray groups’ definition of former user risks comes even closer to what we are terming the “all discontinuations” postexposure risk than the definition used by Nielsen and colleagues. Specifically, an individual’s contribution to the postexposure metric can begin at any point during follow-up and can continue until the end of the intended follow-up period under study, rather than having the universal starting point of 51 days used in the Nielsen and colleagues corticosteroids study.

However, there are at least two distinctions from the use of this approach taken by Nielsen and colleagues and the Ray group in these prior studies and the “all discontinuations” postexposure (adPE) risk metric as we as we have described it. As mentioned, these prior studies do not compare the “all discontinuations” postexposure risk between two intervention arms (i.e., they do not compare the risks that individuals in different intervention arms are experience when they are no longer receiving the intervention). Rather, they compare the “all discontinuations” postexposure (adPE) risk metric values to those the analyses examining the risks seen during current use (i.e., active receipt of the intervention). As also mentioned, a second difference is that their protocols allow patients to reenter the cohort later as “new users” again, whereas in our analyses we only allow individuals to be “first new users” once, and to only contribute to the nonexposure risk metric once. To us, the approach of allowing reentry into the cohort risks introducing some level of the very bias that new user designs are specifically intended to avoid (i.e., allowing individuals to have prior knowledge of how effective or safe and tolerable a medication is for them specifically). (Sometimes the Ray group did perform supplementary analyses in which individuals were only allowed one cohort entry, but they do not appear to have ever reported the former users risks that were observed during those supplementary analyses).

Interesting differences also exist in how soon the period of time begins that was evaluated to determine “former user risks.” For example, the Nielsen et al. 2001 study^2^ had a strict initiation date for postexposure risk (i.e., former user status) of 51 days after medication initiation. The Ray group’s 2002 study instead had former use beginning the day after current use ended^3^ (as defined by the start date and the number of days in the prescription, with apparently no “grace period”). In contrast, the Ray group’s 2009 study allowed for both a 7-day “grace period” after the end of prescribed supply of the medication and a 90-day “indeterminant use” period before individuals became “former users.”^5^ These variations might be expected to affect the extent to which several phenomenon might influence the former user risks, including intermittent use of the intervention, any highly-transient postexposure intervention-related effects, and attrition bias that manifest in events that were experienced very shortly after intervention discontinuation, although this is speculation. (By “extremely short-term attrition bias” we are referring to selection to discontinue the intervention that occurred within days or just a few weeks before the outcome occurred). Future research might examine whether estimates of nonexposure risk metrics become more stable if longer periods of discontinuation are required before starting follow-up time for the postexposure risk metric, and at what point such stability is largely achieved. Of course, as longer periods of discontinuation are required, the postexposure risk metric decreases in its ability to approximate confounding in the early portions of the follow-up period. (To put this topic in simpler language, future research might examine the optimal timing to start measuring a postexposure risk metric).

The Ray group also reported another study using former users in 2004, but here the former users were defined as individuals with use in the last 365 days but seemingly not individuals who had been current users in their study.^11^ Similar definitions have been used in cohort studies by Sorensen and colleagues focusing on risks resulting in hospitalization or occurring after hospitalization (Sorensen was the second author on the Nielsen and colleagues 2001 study^2^). For instance, depending on the study, former use of the intervention was defined as use of the intervention only prior to 60 days^12^ or 125 days^13^ before hospitalization. Since 2001, Sorensen and colleagues have also used definitions of former users in cohort studies in which the former users have used the medication in the past prior to an event rather than after a period of current use that is being studied. (Their study of venous thromboembolism from 2013^14^ is just one example).

Of note, however, it appears from our review of the literature to date that since the Ray group’s 2011 paper^6^, neither the Ray or Sorensen groups (or other researchers) have used the approach of evaluating confounding by examining risks among former users who also were analyzed as current users earlier in the analyses. However, such literature searches are challenging since important details about how former users are being defined or implemented are difficult to identify using computerized literature searches. It is possible that, despite our literature searches, we have overlooked some studies.

If a broader perspective is taken to this topic beyond studies comparing cohorts of individuals, then there are multiple case-control studies in which former users or past users have been examined. In these studies, former users or past users are typically used in addition to nonusers as comparison groups for current users (i.e., individuals who are actively exposed to the intervention at the time of achieving case status).

There is a clear distinction between this approach and our focus in the manuscript, given that these former users are being examined in a case-control study design, not a cohort study design. For instance, typically, the former users group is used as a separate control group to the current users of the medication. This is in contrast to a design in which the former users who have been analyzed are drawn from current users in the same study. Also, similar to the cohort studies mentioned above, these case-control studies do not appear to evaluate former user risks across two or more interventions.

However, sometimes use of a ‘former user” group in a case-control study will be specifically highlighted as helping to address confounding. As an example, in a 2014 study Bruderer and colleagues noted that in their view that their control group of formers users served to “further minimize bias by indication” (which is also termed “residual confounding by indication” in their manuscript).^15^

In another example, we were able to identify a case-control study that used nonexposure risks that presumably occurred postexposure to help interpret their findings (in addition to serving as a formal reference group).^16^ The 2015 study of Lai and colleagues^16^ compared both current use of zopiclone (use within 30 days of a first-time diagnosis of pancreatitis) and “non-active use” (use ≥ 31 days before individuals’ acute pancreatitis diagnosis to “never use” of zopiclone. However, as far as we can determine, despite the fact that “non-active use” could start as soon as day 31 after an exposure, this study should be viewed as implementing a form of “all discontinuations” postexposure (adPE) risk metric within a case-control study rather than a “briefly-exposed” postexposure risk metric. We say this because it appears that some individuals could have exposures that lasted longer than 30 days, and thus their postexposure period would start later as well. It also appears that, if an individual was using zopiclone episodically, there could be multiple periods of current use and non-active use within the year under study, similar to the cohort studies we have discussed above, although this is difficult to determine with certainty. These authors observe an adjusted odds ratio for “non-active use” and pancreatitis (compared to never use) of 2.07 (95% CI 1.60-2.66) a finding which was not extremely different from the adjusted odds ratio for current use of zopiclone and pancreatitis (2.36 [95% CI 1.70 – 3.28]). To their credit, the authors provide considerable discussion about how this association in non-active users could be explained by a confounding effect, while also considering poor compliance or “as needed” use as possible explanations for their former user findings.^16^

The focus of our manuscript is not case-control studies. However, we will note in passing that presumably the same potential nonconfounding influences that we discuss in the manuscript that could affect postexposure risks in studies of cohorts of individuals could also affect postexposure risks in case-control studies. These influences include attrition bias, postexposure intervention-related effects, depletion-of-susceptible-individuals effects, and potentially regression to the mean. Furthermore, concerns about limited representativeness of the postexposure groups may also apply, although these concerns may be reduced if almost everyone who started the medication became a non-active user.

Despite these limitations, we do expect, as other investigators apparently have, that including former users as a control group has improved these case-control studies’ ability to assess or address confounding over and above what would have been possible without examining risks among former users. However, this is also speculation and could benefit from further research.

There are also obvious similarities between the approaches we are proposing as nonexposure risk metrics and the approaches taken in self-controlled case series.^7,17^ In self-controlled case series, risks during periods before and/or after exposure are compared to risk during exposure. The design helps control for confounders that are time-invariant under the entire period under study (e.g., past personal history or family history of a condition). However, by examining risks in studies involving multiple intervention groups, our approaches endeavor to address the totality of both measured and unmeasured time-invariant and time-varying confounding. (As discussed in the manuscript and these supplements, however, there are limits to how well even our proposed approaches can address highly time-varying confounding, due, for instance, to incomplete representation of follow-up time. Those of our proposed metrics that are particularly strong for evaluating time-varying confounding, such as the ultra-briefly-exposed postexposure (ubePE) and assigned-but-not-exposed (abNE) risk metrics, may have other limits to how well they precisely represent confounding due to concerns that they are less likely to be completely representative of the full sample of individuals undergoing evaluation.)

Recently, however, there have been efforts to introduce active comparator designs into self-controlled case series analysis.^18,19^ To the extent that the two comparison groups share similar time-varying confounding, this may allow time-varying confounding to be addressed in self-controlled case series designs as well. We have not considered this question in detail, but one difference between the approaches used in the self-controlled case series and our proposed approaches is that the active comparator self-controlled case series first considers risks within an intervention arm, and then compares those within-intervention arm results to the within-intervention arm results from another intervention group.^19^ Our proposed approaches begin with between-group risk measures for the full cohort under study, and then uses the nonexposure risk metrics to assess how well confounding appears to have been resolved. Examining the value of these alternative approaches for approximating confounding could be a topic for further research, especially in simulation studies.

In addition, there are cohort studies that do not appear to adopt a strict self-controlled case series design that have examined preexposure risk as well. As just one example, the study of Gibbons and colleagues^20^ looked at suicide attempt rates among patients prior to their receiving antiepileptic medications as well as suicide attempt rates during their treatment with antiepileptic medications. However, in this study it is not clear that they examined suicide attempts within a defined preexposure period, although the small number of person years examined suggests that, on average, preexposure follow-up was likely to be within a time period that was fairly proximal to the start of the medication. In addition, the authors used the preexposure period to provide context for the risks observed during treatment within each medication arm, but not to compare preexposure risk across the treatment arms.

Of our four proposed nonexposure risk metrics, as the brief discussion of self-controlled case series and some other cohort studies examining preexposure periods suggests, approaches similar to the baseline preexposure risk metric appear to have the most extensive history of use. We did not include the baseline preexposure (basePreE) risk metric in our set of four proposed nonexposure risk metrics to imply that what we were proposing regarding using baseline risk between treatment groups to assess potential confounding was as novel as the briefly-exposed postexposure (bePE), ultra-briefly-exposed postexposure (ubePE), and assigned-but-not exposed (abNE) risk metrics that we are proposing. Rather we are including the baseline preexposure (basePreE) risk metric in our set of “proposed” metrics because it is one of several nonexposure risk metrics that investigators could consider using for a given research question, and it has some distinct limitations but also some distinct advantages compared to the other nonexposure risk metrics.

In addition, as discussed in Supplement 21, over time it is possible that investigators will start to use multiple nonexposure risk metrics (or multiple approaches to assess or address confounding that includes unmeasured confounding, as some investigators are already doing) to compensate for the limitations of any one metric. Among the nonexposure risk metrics, in addition to the briefly-exposed postexposure (bePE) risk metric, the baseline preexposure (basePreE) risk metric and the “all discontinuations” (adPE) postexposure risk metric would seem to be the candidate approaches that would be the most feasibly implemented in current databases. (See Supplement 21 for more discussion).

Finally, with respect to the fact that our proposed metrics examine nonexposure risks across intervention groups to determine how much these risks diverge from the null, our approach bears some similarities to approaches that use “negative controls.”^1^ Similarly, in our opinion, because three of our four proposed nonexposure risk metrics examine the same individuals who are exposed to the intervention, our study could be viewed as an “internal validation”^21^ approach. However, these are topics that potentially merit greater discussion.

Clearly, even this brief survey of the preceding literature indicates that intermittently for more than twenty years multiple researchers have appreciated the advantages of examining nonexposure risk periods and have employed less-specific variants of the basic approaches we are proposing. The concept of evaluating nonexposure risk has a definite history. We hope our manuscript helps propel this area of epidemiological research forward in three important ways. First, we introduce new nonexposure risk metrics that are specifically constructed to minimize some of the limitations to using nonexposure risk as an approximation of confounding. Second, the manuscript provides (along with these Supplements) a particularly detailed discussion of the many factors to be considered when choosing and interpreting a nonexposure risk metric. The manuscript also delineates the advantages and limitations of various nonexposure metrics that can be considered for use in intervention and exposure studies. Finally, the manuscript shows the potential advantages of using a more specifically-defined nonexposure risk metric such as the briefly-exposed postexposure risk in both the Study Design and Study Analysis phases of a given study. We hope that our work builds in some very helpful ways upon the valuable earlier work about nonexposure risks that has been already done by other researchers.

**Supplement 3.** **Substantial limitations exist to using “all discontinuations” postexposure (adPE) risk as an approximation of confounding**

We view the “all discontinuations” postexposure (adPE) risk metric as generally having more limitations than the briefly-exposed postexposure (bePE) and ultra-briefly-exposed postexposure (ubePE) postexposure risk metrics because this metric can include individuals experiencing longer durations of exposure to the intervention. This longer duration of intervention exposure would be expected to at least potentially allow for greater amounts of attrition bias, postexposure intervention-related effects, or other influences on the metric (e.g., depletion-of-susceptible-individuals effects) to occur. These influences are all threats to the validity of using nonexposure risk metric to approximate confounding since each represents an entirely independent process besides confounding by which nonexposure risks between intervention arms may diverge from the null.

To elaborate: As discussed in the manuscript and Supplement 2 above, we believe the first (and possibly only) postexposure metric to be previously suggested as a potential indicator of confounding was what we are terming “all discontinuations” postexposure (adPE) risk.^2,3^ However, as we describe briefly in the manuscript, multiple fundamental limitations to this metric can exist. The principal limitations relate to the possibility that attrition bias, postexposure intervention-related effects, and depletion-of-susceptible-individuals effects can all potentially interfere, perhaps greatly, with adPE risk serving as an effective approximation of confounding. In addition, the adPE risk typically would not be expected to as effectively approximate time-varying confounding, on average, over as much of the follow-up period as metrics that permit only shorter exposure times (Supplement 5).

To elaborate further on attrition bias, in our view attrition bias can be helpfully seen as “risk shifting”: that is, the shifting of events that would have occurred if exposure had continued from current recipients to individuals in the postexposure period, or potentially to time periods even farther in the future. These future time periods might not be tracked by the time periods under study, or even at all. (For convenience, we refer to risks that do not manifest in outcomes during the follow-up period of interest as “averted risks,” even though it may not be clear if these risks are completely averted or merely shifted to a period beyond the maximum follow-up time period under study).

Not only are risks simply “shifted”, however, but a *reciprocal relationship* can be expected between the changes in observed current recipient risk and the changes in observed postexposure risk that occur as a result of this “risk shifting.” That is, for those risks that are shifted from the current recipient period that do show up in the form of outcomes experienced in the postexposure risk period (i.e., are not “averted”), shifting that *decreases risk* in the current recipient period would be expected to *increase risk* in the postexposure period (all other considerations being equal), and vice versa.

Consider, for illustration, attrition bias that occurs just a day or two before the event occurs. This censoring would then remove this event from what would have been observed if the current recipient periods had lasted a day or two longer. The event would now appear in the postexposure risk period (assuming the individual did not switch treatments or resume their initial treatment in that brief 1- to 2-day span, or that the discontinuation did not stop the event from occurring). Thus, the event count occurring in the current recipient period would decrease by one event from what would have occurred without the cessation of the intervention and would increase by one event in the postexposure period beyond what would have been observed if the cessation of this individual’s intervention exposure had not occurred. This is precisely what we mean by “risk shifting”: in general, non-averted attrition bias would be expected to produce a change in current recipient risks and produce a change in risk in the *opposite* direction in the postexposure risk period.

It must be stressed that while this “risk shifting” does not manifest in the postexposure period if the shifted events *do not occur* in that particular follow-up period, such risk shifting would *nevertheless still affect* the risks observed during the current recipient phase. For example, consider the case in which someone would have experienced an adverse side effect such as hyperlipidemia or chronic kidney disease if a medication exposure had continued for the full 365 days under study. However, let us assume that if the medication is discontinued after only the first 90 days, then this individual will not experience this outcome for several years or more. In this case, the early discontinuation *does* create attrition bias in the current recipient risk, which is going to indicate less risk than if the exposure was continued for 365 days. However, in this particular case there *would not* be a reciprocal increase in risk in the postexposure period over the first 365 days after initiation. In this particular instance, the attrition bias would not interfere with the use of the postexposure risk to index confounding (from the influence of baseline factors over time) observed over the first 365 days. If the shifted event (e.g., hyperlipidemia or chronic kidney disease) *had* *occurred* in the postexposure period in the first 365 days of the study’s follow-up period, however, this shifted event would interfere with the use of the postexposure risk metric as a straightforward index of baseline confounding over the first 365 days from intervention initiation.

Because of this potential reciprocal relationship between current recipient and postexposure risk when attrition bias occurs, this “risk shifting” can reduce, cancel out, or even reverse the direction of the postexposure risk from the direction of actual confounding (as long as the shifted events are detected in the postexposure risk period because they are still occurring within the time period under study). The more current recipient time that is allowed for individuals who subsequently contribute to the postexposure risk, the more time is present for attrition bias to occur, and the more potential exists for attrition bias to substantially distort the postexposure risk away from simply reflecting confounding. In our view, this uncertainty about the degree to which attrition bias may be influencing postexposure risks greatly limits the use of the “all discontinuations” postexposure risk metric to approximate confounding.

In the manuscript, we primarily describe the impact of this reciprocal relationship upon an example in which biases from attrition and from confounding bias the main analysis effect estimate in the same direction during active receipt of the intervention. We expect this circumstance to be the most common scenario encountered in studies of cohorts of individuals, since this would reflect a consistent “selective pressure” to assign and maintain lower- or higher-risk individuals on a certain intervention. However, circumstances can certainly be envisioned in which attrition bias would bias the main analysis effect estimate in the opposite direction as confounding. In this case, to the extent that attrition bias occurs, the reciprocal nature of its impact on postexposure risk would produce a postexposure risk that overestimates confounding (if the postexposure risk was interpreted as reflecting confounding, and with all other considerations being equal). Consider a study where higher risk individuals receive Intervention A than Intervention B, but higher risk individuals are then preferentially discontinued from Intervention A compared to Invention B. The excess events that occurred in the postexposure period of Intervention A from the attrition bias removing higher-risk individuals from Intervention A (it these events were not “averted” so that they did not appear in the postexposure risk period) would add onto the excess postexposure risk for Intervention A as a result of confounding. Thus, if the postexposure risk was interpreted as approximating confounding, confounding would be overestimated. (By “overestimated”, we mean attrition bias would cause the postexposure risk estimate to diverge more from the null, either in a positive or negative direction, than it would if based just on confounding. This just represents the other possible scenario from the situation when attrition bias and confounding affect the main analysis effect estimate in the same direction. In this circumstance when attrition and confounding are biasing the main analysis effect estimate in the same direction, as discussed in the manuscript, the postexposure risk would underestimate confounding, or, in more extreme cases, even reverse the direction of postexposure risks from the direction of confounding).

Another potential disadvantage of the “all discontinuations” postexposure risk is that, for at least some individuals, it gives maximum amount of time for postexposure intervention-related effects to develop. That is, postexposure intervention-related effects would be of greatest concern typically among individuals discontinuing or being discontinued from the intervention particularly late in the study period, although there are certainly examples where brief interventions can have long-lasting postexposure intervention-related effects (e.g., vaccines). The impact of postexposure intervention-related effects can be either in the same direction or the opposite direction of the actual intervention effect. Since postexposure intervention-related effects can occur in either direction, these effects can also (along with attrition bias effects) affect the postexposure risks either in the same or the reciprocal direction as confounding.

Stated in more detail, postexposure intervention-related effects can be of two types. For convenience, we term one type ”persistent” effects (effects that are in the same direction as the intervention effect during receipt of the intervention) and the other type “discontinuation” effects (effects that generally would be expected to be in the opposite direction of the effect observed during receipt of the intervention, such as from a period of physiological “rebound”).

Also, while it is clear that shortening exposure time generally would be expected to reduce attrition bias, in some cases this shortening may not tangibly impact postexposure intervention-related effects. Two examples spring to mind: 1) a medication that starts to be effective very quickly and which has durable effects (e.g., a vaccine) would be expected to have a rapid onset of persistent effects. Thus, shortening exposure time would not necessarily substantially lessen this effect, until perhaps some minimal exposure was reached for which persistent effects were not as firmly established. (In the case of a vaccine, this threshold might never be crossed and a relatively constant postexposure effect might be observed after only a single exposure); 2) a medication that fairly rapidly conditions the body so that discontinuation effects are observed if the intervention is discontinued even fairly soon after it is initiated.

In some cases, prior knowledge or reasonable suppositions can be made about postexposure effects. (For example, for some medications whose active presence is important for its effects, such as some antibiotics, it may be reasonable to assume that persistent effects are not more potent than the actual exposure itself. That is, in some cases persistent effects might only occur at equal or attenuated effect sizes to those observed during exposure, and perhaps for no longer in duration than a period after exposure that equals the duration of the actual exposure itself.) However, rigorously determining if postexposure intervention-related effects are present or are even to be expected can be very challenging without sizable prior knowledge about the intervention. Thus, the optimal strategy for addressing the possibility of postexposure effects may be to establish conditions (such as brief exposure, ultra-brief exposure, and assigned-but-not-exposed conditions) where the chance of such effects is minimized. (See Supplement 21 for additional discussion). Finally, a briefer exposure time would also be expected to minimize time for effects related to the depletion-of-susceptible-individuals effects^22,23^ (see Supplement 4) to develop.

This supplement helps reinforce why we go to considerable lengths in our manuscript to always refer to nonexposure risk metrics as being intended to “approximate,” rather than “estimate,” confounding (see Supplement 1). In our view, even though both terms convey some degree of uncertainty, “approximate” implies less precision than “estimate.”

**Supplement 4.** **Possible influence of depletion-of-susceptible-individuals effects on nonexposure risk metrics**

Depletion-of-susceptible-individuals effects^22,23^ are a phenomenon in which risks are transiently lowered postexposure because members of a particularly high-risk pool have been “depleted” by having the occurrence of the outcome advanced in time with their initial exposure to an intervention. (This discussion assumes that the intervention increases the risk of the outcome, on average). However, for most studies the presence or magnitude of this phenomenon relative to attrition bias or postexposure intervention-related effects is uncertain, and likely would vary by outcome.

To elaborate: Depletion-of-susceptible-individuals effects are a possible bias affecting longitudinal studies. The bias results when the early impact of an intervention or exposure on a sample of individuals results in lower rates being observed associated with that intervention or exposure later in time if the most susceptible individuals of that sample tend to experience the effects sooner after initiation of the intervention or exposure than less susceptible individuals. (Put another way, if some individuals are near the threshold of developing a condition, and a sufficiently potent “insult” in the form of an intervention or exposure occurs that happens to push them over the threshold for experiencing an event at an accelerated rate, this can result in the relative absence of people close to the threshold for the event remaining in the sample for a period of time. This would result in a time period in which the risks of the event associated with the intervention are lower than previously, and quite possibly lower than the usual event rate associated with the intervention). Usually, this bias is thought to affect the remaining sample of individuals who are being exposed to the intervention or exposure. (For instance, over time a higher rate of events would be observed early during follow-up, followed by a relatively depressed rate of events subsequently due to depletion-of-susceptible-individuals effects, followed then by a more consistent intermediate rate subsequently). However, there is no reason why the temporarily-depleted rate of subsequent events expected to result from this bias could also not manifest in individuals who were initially exposed to the intervention or agent but have now entered a nonexposure period, especially early during that nonexposure period.

In the context of using nonexposure risk metrics that examine risks during nonexposure that occur after the initiation of the intervention (i.e., postexposure risks) in an effort to approximate confounding, depletion-of-susceptible-individuals effects represent another phenomenon separate from confounding that could cause postexposure risks to differ between two or more intervention or exposure arms. This difference can occur in different ways, for instance, one of the interventions has an effect (assumed here, for the purpose of illustration, to be an effect which increases the event rate) that specifically leads to the depletion-of-susceptible-individuals phenomenon (i.e., initially, there is a period with an higher-than-average event rate associated with the intervention, followed by period exhibiting a drop in the rate observed for that intervention arm to a lower-than-average event rate, which is then followed by a third period featuring a more consistent average event rate after those first two periods). For the purposes of this illustration, let us assume that the other intervention does not have an effect. In this second arm, there would therefore be no depletion-of-susceptible-individuals and thus no decrease in event rate for this arm later during follow-up. Comparing the postexposure risk between these two groups would be expected to produce a divergence from the null with lower postexposure risks associated with the intervention that had a genuine effect (i.e., that increased event counts while the intervention is in effect) if the first period of elevated rates was included in the exposure period, and the second period of decreased rates was included in the postexposure risk. (To be clear, there would not be a third period, because exposure would not be continuing, but it is plausible that the depletion-of-susceptible-individuals effects could reduce the risk seen in the first intervention arm to below the baseline risk for those individuals for a period, depressing the overall rate of events observed in the postexposure risk period below that that would be observed for the second intervention arm.)

Another scenario that could produce depletion-of-susceptible-individuals effects is simply if the two interventions had differed in the potency with which they induced increased or decreased event rates. In this case, the differential effects of the two interventions could result in a greater depletion-of-susceptible-individuals effects in one intervention arm relative to the other, and thus postexposure risks that, when compared between the two arms, diverged from the null.

Of note, compared to the other nonconfounding influences on postexposure risk, there is a pattern that would generally be expected for depletion-of-susceptible-individuals effects: the phenomenon would be expected to be observed in the intervention group associated with the higher rate of events during the exposure period, leading to lower rates of events observed, at least transiently, during the postexposure period. To put this in the context of our proposed briefly-exposed postexposure (bePE) and ultra-briefly-exposed postexposure (ubePE) risk metrics, in general, a depletion-of-susceptible-individuals effect would not be expected to occur in one intervention arm relative to the other without the presence of a genuine elevated risk in that intervention arm (relative to the other) occurring during the brief or ultra-brief exposure period.

Of course, the challenge is knowing if such a genuine elevated risk is occurring. There are circumstances in which an apparent elevated risk during exposure may occur but could be attributable to confounding, random error, or other biases rather than a genuine treatment effect. Similarly, instances can be foreseen in which no elevated risk is observed, but such an effect from the intervention is actually present but simply concealed by confounding, random error, or other biases. One reassuring consideration, however, is that in general depletion-of-susceptible-individuals effects may be relatively modest. This is especially true if the entire postexposure period being assessed is relatively long (and contains a number of outcome events) relative to the generally more transient period that would be expected to be affected by depletion-of-susceptible individual effects. Nonetheless, this topic could probably benefit from further research and thus represents another area for investigation.

Nevertheless, it should be noted that at least the potential exists for a more predictable direction of a depletion-of-susceptible-individuals effect than other potential nonconfounding influences on the postexposure risk. Specifically, there would be uncertainties about the direction of effects that might occur in relation to attrition bias (since it may be unclear which group may be experience attrition specifically due to factors related to the outcome, and in which direction this attrition may occur), postexposure intervention-related effects (since it may be unclear which group may have stronger postexposure intervention-related effects, and in which direction these effects occur), and regression to the mean (since it may be unclear which group is experiencing greater initial selection based on particular measurements or other factors). In contrast, depletion of susceptible individuals effects, to the extent that they occur, generally would be expected to be observed in the intervention arm showing increased risk during exposure, especially if that increased risk was greater than what was observed at baseline.

Also, of note, one approach to minimizing depletion-of-susceptible-individuals effects is the same approach used to potentially minimize attrition bias or postexposure intervention-related effects: minimize exposure time, thus minimizing the time for all of these effects to occur. In addition, typically, minimizing exposure time would lengthen follow-up time so that the portion of follow-up time affected by the depletion-of susceptible-individuals-effects would constitute less of the total follow-up time being evaluated. This minimization of exposure time is what both the briefly-exposed postexposure (bePE) risk metric and, more extremely, the ultra-briefly-exposed postexposure (ubePE) risk metric attempt to do. Minimizing the risk of this phenomenon is also important since two or more nonconfounding influences on the postexposure risk can occur. In such a circumstance, it is entirely possible no easily recognized pattern of risk occurs, even if depletion-of-susceptible-individuals effects are occurring.

There is one possible exception to this expectation that minimizing exposure time would minimize time for depletion-of-susceptible-individuals effects to occur. If, for instance, the brief exposure time was still sufficient for depletion-of-susceptible-individuals effects to arise (e.g., the intervention very rapidly depletes the susceptible individuals by producing effects in those individuals very shortly after the intervention starts), then in this instance it is possible that depletion-of-susceptible-individuals effects might be more noticeable in the briefly-exposed postexposure risk than the “all discontinuations” postexposure risk. Put another way, it is not an absolute rule that the briefly-exposed postexposure (bePE) risk metric would be expected to show less depletion-of-susceptible-individuals effects than the “all discontinuations” postexposure (adPE) risk metric. In some cases, the timing of the depletion and the timing of the start of the briefly-exposed postexposure risk might be fortuitously closely aligned in a manner so that the briefly-exposed postexposure (bePE) risk metric might be more sensitive to these effects than the “all discontinuations” postexposure (adPE) risk metric. This is another area that might warrant research in the future.

To reiterate, in general the most optimal approach for using postexposure risks to approximate confounding would appear to be to minimize exposure time as much as feasible in order to limit the amount of time for attrition bias, postexposure intervention effects, *and* depletion-of-susceptible-individuals effects to occur. For cohorts of the size we are studying, the briefly-exposed exposure period appeared to be the briefest exposure for us to feasibly use, considering likely random error. However, in much larger cohorts or in the future, the ultra-briefly-exposed (ubePE) postexposure risk metric or the assigned-but-not-exposed (abNE) risk metric may become feasible. If concerns about representativeness and random error are not too pronounced, these nonexposure risk metrics potentially may be even more desirable in some cases than the briefly-exposed postexposure (bePE) risk metric.

**Supplement 5. Incomplete representation of follow-up time**

All of our proposed nonexposure risk metrics except the assigned-but-not-exposed (abNE) risk metric do not approximate risks over the entirety of the follow-up time. Rather, there are days at the beginning of the follow-up period under study for the main analysis that are not reflected well in the postexposure risk metrics because these are the days for which exposure is allowed. Furthermore, the baseline preexposure (basePreE) risk metric does not evaluate the follow-up period under study for the main analysis at all. Estimates of postexposure risk, for instance, very early in the follow-up period will not be informed by as many individuals as will the estimates of postexposure risk later during follow-up. However, the significance of this “information deficit” is uncertain and likely varies for each analysis. For instance, the time period for which this “information deficit” occurs for the briefly-exposed postexposure (bePE) and ultra-briefly-exposed postexposure (ubePE) risk metrics is brief or very brief. Thus, this deficit likely will not have substantial impact unless there is a substantial difference in risk between the intervention arms for the specific time period falling within this “information deficit.”

To elaborate: Due to how it is defined, the briefly-exposed postexposure (bePE) risk metric would be expected to, in general, quantitatively approximate confounding less precisely over the follow-up time that consists of the days of exposure to the intervention that is allowed for the individuals being included in the metric (as well as any allowed “grace period”). For our definition of the briefly-exposed postexposure (bePE) risk metric used in the manuscript, for instance, the metric can fail to include, for particular individuals, any of the days for which they are exposed (i.e., from 1 – 30 days), plus a grace period totaling up to 17 days (for a total unrepresented period for some individuals of as much as 47 days). There will be some representation of the postexposure risk over days earlier than 47 days that will be provided, for example, by individuals who only had a single-day exposure, or 7-day or 14-day exposures to the intervention. These individuals, however, constitute only a small subset of the briefly-exposed sample examined in the manuscript. Put another way, for the briefly-exposed postexposure (bePE) risk metric analyses shown in our manuscript, it is reasonable to assume that the metric does not characterize the risks observed over the first 47 days of follow-up as well as it does for later times in the follow-up period.

The ultra-briefly-exposed postexposure (ubePE) risk metric would be expected to have considerably fewer concerns related to incomplete representation of follow-up time than the briefly-exposed postexposure (bePE) risk metric, since the allowed exposure (and corresponding grace period) is expected to be so much shorter.

In theory, the assigned-but-not-exposed (abNE) risk metric would be assumed not to have any concerns about incomplete representation of follow-up time, given that risk can be followed from the intervention assignment date (or, in other manifestations, the intended or presumed intended initiation date). In practice, however, the distribution of nonexposed days for the assigned-but-not-exposed (abNE) risk metric could differ from the distribution of days evaluated in the main analysis. Thus, there still could be some “relative incompleteness” in its representation of follow-up time.

Of note, the greatest concerns about representation of follow-up time could be expected to exist for the baseline preexposure (basePreE) risk metric. In a technical sense, the baseline preexposure (basePreE) risk metric includes none of the follow-up time being examined to assess the intervention in the main analysis. Thus, strictly speaking, this metric could be viewed as “representing” none of the follow-up time examined by the main analysis. However, for this metric the risks observed prior to exposure, but close to the time of initiation, are assumed to act as a surrogate for the confounding that is influencing risks throughout the intervention exposure. As discussed elsewhere, this assumption is likely not to be strictly accurate for highly time-varying confounding around the time of intervention initiation. However, this disadvantage may be partly counterbalanced by the baseline preexposure (basePreE) risk metric’s ability to sometimes approximate confounding for all or most of the cohort being studied. Perhaps a more important advantage of the baseline preexposure (basePreE) risk metric is the fact that this metric, being a preexposure risk metric, would not be expected to be affected by many of the potential influences on postexposure risk that can affect the postexposure risk metrics. (See Supplement 21 for more discussion).

The “all discontinuations” postexposure (adPE) risk metrics, in general, would be thought to have more concerns about incomplete representation of follow-up time than the briefly-exposed postexposure (bePE) risk metric or the ultra-briefly-exposed postexposure (ubePE) risk metric. However, the issue is nuanced. To consider this issue more fully, it is worth noting that the “all discontinuations” postexposure (adPE), briefly-exposed postexposure (bePE), and ultra-briefly-exposed postexposure (ubePE) risk metrics all contain some of the same individuals. So, the issue does not relate to the ultra-briefly-exposed postexposure (ubePE) or briefly-exposed postexposure (bePE) metrics containing individuals who start the postexposure period sooner, but rather that *on average* the postexposure period for the ultra-briefly-exposed postexposure (ubePE) and briefly-exposed postexposure (bePE) metrics start sooner. The representations that those postexposure metrics provide of the days early during the time period being evaluated by the main analysis would not be expected to be as “washed out” in the metric as for the “all discontinuations” postexposure (adPE) risk metric. This relative lack of representation of earlier follow-up in the adPE risk metric arises because individuals are included in the metric that start the postexposure period later (sometimes much later) in the follow-up period used for the main analysis.

To be specific, the “all discontinuations” postexposure (adPE) and briefly-exposed postexposure (bePE) risk metrics also contain additional individuals relative to the ultra-briefly-exposed postexposure (ubePE) risk metric. For example, if there is an individual exposed to the intervention for only a single day included within a study that is following individuals for up to 365 days, that individual’s nonexposure time will be included in all three metrics. However, the more that additional individuals are included in the metric whose postexposure periods started later in follow-up, the less the risk estimate provided by the postexposure metric would be expected to represent confounding well early in the time period under study, if time-varying confounding was present. (As indicated in Supplement 13, there may be ways to conveniently summarize how well the nonexposure risk metric is capturing risks close to intervention initiation, either by reporting the average start time for the nonexposure period among individuals included in the metric, or perhaps the 20^th^ or 25^th^ percentile value. There is likely no perfect approach to this concern, however, other than recognizing that, in general, the briefer the period of exposure allows a greater period of time over which confounding can be approximated by a postexposure risk metric.)

It is also worth noting that this potential issue becomes a concern only to the degree with which the relative risks between the two intervention arms that would be observed early in the follow-up period diverge from those seen later in the nonexposure period. In some cases, it may be a reasonable pragmatic assumption to assume that these risks do not differ, or do not differ by a large amount. However, because confounding is quite often time varying, and sometimes highly time varying, it is reasonable to expect that, in a number of circumstances, relative differences in risk between the two intervention arms arising from confounding may be greater earlier in the follow-up period than later.

We now discuss this issue of representation of follow-up time specifically in reference to the analyses we provide in the manuscript. Supplementary Table 1A shows the average starting date of the postexposure period for both the briefly-exposed postexposure (bePE) risk metric and the “all discontinuations” postexposure (adPE) risk metric (for our all-cause mortality and adverse event analyses). Supplementary Table 1A indicates that, on average, the briefly-exposed postexposure (bePE) risk metric’s postexposure period starts much earlier (day 46 after the intervention start date) than the “all discontinuations” postexposure (adPE) risk metric’s postexposure period (day 100 after the intervention start date). In general, it would be expected that in instances in which time-varying confounding is present, this earlier average start time could be a significant advantage. In circumstances in which confounding is greater early in the follow-up time period, this early portion of the follow-up period would be more likely to be affected by these larger amounts of confounding, making the briefly-exposed postexposure (bePE) metric’s typically increased ability to sensitively reflect these early differences in confounding between the intervention arms particularly important.

In addition, the average days of follow-up for each postexposure metric is provided. Of note, the average duration of the briefly-exposed postexposure (bePE) risk metric’s follow-up period is from 123 – 126% as long as the average duration of the “all discontinuations” postexposure (adPE) risk metric. This indicates that, on average, at least in these analyses, the briefly-exposed postexposure (bePE) risk metric represents risks during nonexposure that are observed over a larger percentage of the entire follow-up period under study. We expect this to be true in most applications of the briefly-exposed postexposure risk (bePE) risk metric, by virtue of its generally earlier start, on average, than the “all discontinuations” postexposure (adPE) risk metric.

However, it is also possible for a postexposure risk metric to represent *too much* follow-up time. It is disadvantageous if the nonexposure metric reflects considerable follow-up time that occurs after the follow-up time which is typically contained in the main analysis. This additional follow-up time would be expected to “wash out” or “dilute” the risk signal from the follow-up time that the metric has in common with the follow-up time examined in the main analysis.

This concern may be especially prominent for current recipient analyses in many medication studies. Current recipient analyses would often to be expected to end, on average, well before the maximum duration of the period under study, as individuals discontinue or change their initial intervention. As a result, especially in current recipient studies with low subsequent resumption or switching rates, postexposure risk metrics may be prone to overrepresenting follow-up time. This topic of how to best address imprecise representation of main analysis follow-up time related to the average termination date of the nonexposure risk metric is one of many aspects of nonexposure risk metrics that could warrant further study. For instance, it may be useful to investigate whether it would be worthwhile to arbitrarily censor individuals in the postexposure period earlier than the end of the study period to bring the average stop date of nonexposure risk follow-up closer to that of current recipients. Alternatively, one could investigate the strategy of choosing the largest sample of briefly-exposed individuals that provides a distribution of follow-up termination dates that more closely approximates that of the current recipient analysis.

| **Supplementary Table 1A:** **Information about the Follow-up Time Captured by the**  **briefly-exposed postexposure (bePE) and “all discontinuations” postexposure (adPE) risk metrics**  (365-day high-dimensional propensity score (hdPS)-matched Primary Analyses) | | | | | | | | |
| --- | --- | --- | --- | --- | --- | --- | --- | --- |
| **Outcome** | **bePE Risk Metric**  **Mean Start of Follow-up Time**  **(Days after Intervention Start)** | **Approximate**  **adPE Risk Metric**  **Mean Start of Follow-up Time**  **(Days after Intervention start)** | | **bePE Mean Follow-up (Days)** | | **Approximate**  **adPE Mean Follow-up (Days)** | | **Percentage of adPE Follow-up Time represented by**  **bePE**  **Follow-up Time** |
| All-Cause Mortality | 46 | 100 | | 203 | | 161 | | 126% |
| ≥ 15% Weight Gain | 46 | 99 | | 198 | | 160 | | 124% |
| Hypertension | 45 | 98 | | 196 | | 159 | | 123% |
| Hyperlipidemia | 45 | 95 | | 195 | | 158 | | 123% |
| **Supplementary Table 1B:** **Termination of Follow-up Time for the Main Analysis, briefly-exposed postexposure (bePE) risk metric and “all discontinuations” postexposure (adPE) risk metric**  (365-day high-dimensional propensity score (hdPS)-matched Primary Analyses) | | | | | | | | |
| **Outcome** | **Main Analysis** | | | | **Nonexposure (Postexposure) Risk Metric** | | | |
|  | **Average end to Main Analysis**  **Follow-up**  **(Current Recipient Analysis)**  **(Mean Days from Intervention Start)** | | **Average end to Main Analysis**  **Follow-up**  **(acITT^1^ Analysis)**  **(Mean Days from Intervention Start)** | | **Average end to bePE Risk Metric**  **Follow-up**  **(Mean Days from Intervention Start)** | | **Average end to adPE Risk Metric**  **Follow-up**  **(Mean Days from Intervention Start)** | |
| All-Cause Mortality | NA | | 271 | | 241 | | 253 | |
| ≥ 15% Weight Gain | 141 | | NA | | 236 | | 247 | |
| Hypertension | 135 | | NA | | 233 | | 243 | |
| Hyperlipidemia | 130 | | NA | | 231 | | 238 | |
| ^1^ “acITT analysis” = “additionally-censored intent-to-treat analysis”, our primary analysis of all-cause mortality (also called the “modified intent-to-treat analysis” in the manuscript). | | | | | | | | |

Supplementary Table 1B illustrates these points about the average termination of the nonexposure risk metric. Both the briefly-exposed postexposure (bePE) and “all discontinuations” postexposure (adPE) risk metrics overrepresent follow-up time for those analyses evaluated using current recipient analyses (≥ 15% Weight Gain, Hypertension, Hyperlipidemia). However, the briefly-exposed postexposure (bePE) risk metric overrepresents the duration of the follow-up period to a lesser extent than the “all discontinuations” postexposure (adPE) risk. (The briefly-exposed postexposure [bePE] risk metric ends, on average, on days 231-236 of the follow-up period, whereas the “all discontinuations” postexposure (adPE) risk metric does not end, on average, until day 238-247 of the follow-up period.) Although this difference is relatively modest, this does suggest that the briefly-exposed postexposure (bePE) risk metric’s postexposure follow-up time is more closely approximating the period that current recipients are exposed to the intervention than the “all discontinuations” postexposure (adPE) risk metric.

It might be tempting, from a superficial examination of this data as presented in Supplementary Table 1B, to conclude that the briefly-exposed postexposure (bePE) risk metric will perform better as an approximation of confounding than the “all discontinuations” postexposure (adPE) risk metric for current recipient analysis, while the “all discontinuations” postexposure (adPE) risk metric will perform better for the modified intent-to-treat analyses. This impression could arise from a comparison of how closely the briefly-exposed postexposure and “all discontinuations” postexposure follow-up times given in Supplementary Table 1B align with the follow-up time of the main analysis. However, considering both the representation of follow-up time and other relevant factors, we think this viewpoint that the “all discontinuations” postexposure (adPE) risk metric may be a preferred choice for modified intent-to-treat analyses would be almost certainly erroneous. Careful examination of Supplementary Table 1B suggests that the briefly-exposed postexposure (bePE) risk metric should be preferred for our mortality analysis (which was a modified intent-to-treat analysis), even based simply on considerations of follow-up time representation. While the “all discontinuations” risk metric terminates, on average, later than the briefly-exposed postexposure (bePE) risk metric, the “all discontinuations” postexposure (adPE) risk metric does not provide better “coverage” of the overall main analysis evaluation period than the briefly-exposed postexposure (bePE) risk metric. It must be remembered that Supplementary Table 1A indicates that the briefly-exposed postexposure (bePE) risk metric starts, on average, considerably sooner (and thus captures more days of early follow-up and, importantly, more days of total follow-up). Supplementary Table 1A actually makes this point explicitly, indicating that the briefly-exposed postexposure (bePE) risk metric represents approximately 25% more follow-up time than the “all discontinuations” postexposure (adPE) risk metric. So, we believe that the briefly-exposed postexposure (bePE) risk metric is the preferred nonexposure risk metric for all the outcomes examined in the manuscript.

In addition, consideration of other factors also suggest that the briefly-exposed postexposure (bePE) risk metric would be a better choice than the “all discontinuations” postexposure (adPE) risk metric. Most importantly, as outlined in the manuscript, the “all discontinuations” postexposure (adPE) risk metric would be expected to be more affected by nonconfounding influences on the postexposure risk than the briefly-exposed postexposure (bePE) risk metric.

One final point: although, for simplicity in the manuscript we referred to “incomplete representation of follow-up” time, in this Supplement we can see that that this term is somewhat inadequate. We chose “incomplete representation of follow-up time” for the manuscript for two reasons. First, we suspect that poor representation of early follow-up time is a particularly important facet of the general issue involving the divergence of the follow-up time evaluated in the main analysis and the follow-up time evaluated, on average, in the nonexposure risk metrics. Second, the term “incomplete representation” is very descriptive of the core issue – that there are parts of the follow-up period that the nonexposure metric does not represent well. Thus, we used this term in the main manuscript. (This is similar to how we emphasized “persistence effects” in the manuscript when we gave a specific example of postexposure intervention-related effects, rather than focus on the more complete concept that postexposure intervention-related effects can include either persistence effects or discontinuation risks). In this Supplement we have seen that it is also possible for the nonexposure risk metric to unfortunately achieve overrepresentation of the follow-up period (i.e., inclusion of days of follow-up beyond those days that are, on average, evaluated for the main analysis endpoint). For this reason, a more precise term would be “misrepresentation of the follow-up period” or, perhaps better, “imprecise representation of the follow-up period.” We favor the latter term because “misrepresentation” can sometimes convey a sense of a value concocted to falsely present information about that topic, perhaps even for nefarious purposes. In contrast, even in the typical case when the nonexposure metric does not completely reflect the main analysis follow-up time, there is typically overlap in the days evaluated by the main analysis and the nonexposure risk metric (the exception being the baseline preexposure [basePreE] risk metric). For this reason, we favor the term “imprecise” or “imperfect” representation of follow-up time. (We will use the term “imprecise representation of follow-up time,” for example, in Supplements 21 and 22).

**Supplement 6. Further thoughts about potential influences on nonexposure risk metrics, especially regression to the mean**

Regression to the mean can result in changes in event counts or outcome values occurring during exposure (or presumably postexposure), typically when the characteristic making up the outcome (e.g., blood pressure, depression severity) is being measured prior to allocation to the intervention or exposure *and* decisions about who receives the intervention or exposure are affected by values of that measure. How often this phenomenon occurs for other types of outcomes, such as the new onset of complexly-defined conditions, is uncertain.

To elaborate: We will use this section to discuss a couple nuances concerning regression to the mean to augment the basic summary of the issue presented above. The first nuance we will discuss is that, as far as we can determine, minimizing exposure time (an important approach for attempting to minimize the risk of attrition bias, postexposure intervention-related effects, and depletion-of-susceptible-individuals effects) would not be expected to address concerns about regression to the mean. Regression to the mean arises from a selection process (deliberate or otherwise) occurring before or at the point of initiation of the intervention or exposure. This selection process is based on variation (often with a component of random variation) in a measure or patient characteristic. As such, this selection process has already occurred prior to the start of the postexposure period. Therefore, we would expect regression to the mean to be relatively unaffected by differences in exposure time. The second nuanced point is that we would expect regression to the mean to potentially affect all of our proposed nonexposure risk metrics (for those outcomes susceptible to regression to the mean). We believe this to be the case because regression to the mean arises from a phenomenon that impacts both the baseline measures relating to an outcome and the measures obtained after the intervention has been initiated (either during active exposure or during a period of nonexposure). Thus, the baseline preexposure (basePreE) risk metric would be expected to be affected, as well as the three proposed metrics examining nonexposure risk after the intervention starts (i.e., the assigned-but-not-exposed [abNE] risk metric, the ultra-briefly-exposed postexposure [ubePE] risk metric, and the briefly-exposed [bePE] postexposure risk metric).

It should be clarified that it is our understanding that typically for regression to the mean to occur, selection to receive the intervention needs to be based on some measurable quantity thought to be predictive of outcome or, perhaps, non-completely measurable impressions of how pronouncedly an individual needs or is appropriate for the intervention. Thus, not all outcomes would be expected to be sensitive to regression to the mean.

We should also clarify that we are discussing regression to the mean in the context of a study involving two or more intervention arms. So, what we are really concerned with is *differences in the amount of regression to the mean* occurring in the different intervention arms. Furthermore, the problems posed by regression to the mean can be a particular concern in at least two circumstances: 1) when some type of pre-post comparison is performed (e.g., change in blood pressure, if measures after the intervention starts are compared with measures taken at baseline), and 2) in a design when only values observed after the intervention starts are compared between two or more intervention arms *if* baseline characteristics predictive of the outcome are analytically brought into closer balance between individuals, as usually happened in matched and stratified analyses. In this circumstance, if, for example, a particular value was elevated above the mean at time of medication initiation in one Intervention arm (“Intervention A”), then bringing this elevated value into balance compared to values that are obtained for the other intervention arm (“Intervention B”), which is not under the same selection, can help establish conditions where the values after initiation observed in the Intervention A arm relative to the Intervention B arm can be expected to be lower, based on the values in the Intervention A arm “regressing” to the mean value.

To give a concrete example, consider a propensity score matched study examining the blood pressure safety of two antidepressant medications during usual care. If one medication (Antidepressant A) is already suspected or known to increase blood pressure, then there may be a tendency for clinicians not to start the medication for individuals with higher blood pressures at baseline. This same selective pressure may not exist for Antidepressant B. Thus, regardless of whether baseline blood pressure values are brought into close balance (e.g., through propensity score matching), then conditions may exist in which individuals on average in the Antidepressant A arm of the study exhibit lower than their mean blood pressure at baseline. (This lower-than-average blood pressure would be due to the normal variability in blood pressure combined with a systematic selection force to start Antidepressant A only in people with low-to-moderate blood pressure). If this selective force resulted in some individuals being selected for Antidepressant A due to variability in blood pressure leading to them having a lower-than-typical blood pressure at baseline, then these individuals will, on average, exhibit an apparent “increase” in blood pressure in their next blood pressure reading or readings. This occurs because, on average, the individuals’ typical blood pressure is higher than what they exhibited at baseline when they were selected to start Intervention A, thus the blood pressure readings are expected to “regress to the mean.” (In this case “regressing” to the mean actually would be observed as an increase in blood pressure). In this manner, Antidepressant A may appear to have a greater effect on blood pressure during exposure than the medication actually causes, due to this phenomenon of “regression to the mean.”

The above discussion about regression to the mean in general could also apply, however, to the values that are used to determine event/outcomes rates in the nonexposure risk metrics as well. It is interesting to note, however, that for a given analysis susceptible to regression to the mean, typically when regression to the mean is present, the baseline preexposure (basePreE) risk metric might be expected to deviate in the *opposite* direction from the null than the direction of deviation expected from regression to the mean for the other nonexposure metrics. In circumstances in which regression to the mean may be particularly prominent, it might be possible to exploit this difference by examining, for example, both the baseline preexposure (basePreE) risk metric and the briefly-exposed postexposure (bePE) risk metric to potentially detect instances when regression to the mean is substantially impacting the nonexposure risk metrics. This is another potential topic for future research.

The complexities around regression to the mean suggest that one avenue of additional research for nonexposure risk metrics is to evaluate quality assessments that help detect if regression to the mean might be occurring. Furthermore, we expect that future research would be valuable that examined how commonly, how extensively, and under what conditions regression to the mean may affect our proposed nonexposure risk metrics and how to best detect these effects.

**Supplement 7.** **The assigned-but-not-exposed risk metric and ultra-briefly-exposed postexposure risk metric are likely to have particularly pronounced random error**

In most instances, the assigned-but-not-exposed (abNE) risk metric, and to a lesser extent the ultra-briefly-exposed (ubePE) risk metric, would be expected to be particularly susceptible to random error due to small sample sizes. In specific, the assigned-but-not-exposed (abNE) risk metric and the ultra-briefly-exposed postexposure (ubePE) risk metric would be expected to typically be much more susceptible to random error than the briefly-exposed postexposure (bePE) risk metric and especially the baseline preexposure (basePreE) risk metric.

To elaborate: The statement in the manuscript that some nonexposure risk metrics may only “become practical as databases become larger in the future” is expected to be most applicable to the assigned-but-not-exposed (abNE) risk metric, which is expected to typically have the smallest sample size of any of the proposed metrics. Perhaps the simplest approach for using this metric in treatment studies would be to examine individuals who were assigned to receive the treatment but never initiated the treatment. For nonrandomized studies of medications, for example, this might consist of individuals who were dispensed a medication but never actually received it (e.g., they did not present to a pharmacy to pick up the medication). The number of people meeting this criterion in current structured databases is likely to be exceedingly small in most cases but would be also expected to increase in number as databases expand with time to include ever larger numbers of individuals.

Until then, another approach worth considering outside of using structured data to identify assigned-but-not-exposed individuals would be to augment the cohort of individuals identified with structured data with individuals who received the medication, but chart review or natural language processing of their record indicated that they did not actually use the medication. For instance, they may have told their provider subsequently that despite picking up the medication, they never actually took any doses. This expanded search strategy to identify members of the assigned-but-not-exposed sample might add a considerable number of individuals to the sample.

However, under such an augmented approach of identifying assigned-but-not-exposed individuals, it would seem that follow-up time for those individuals who received the intervention but did not use it should only start with the date at which the individual informed others they had not used the treatment. ^24,25^ Otherwise, in our judgment, the potential for immortal person-time bias would be created. Nevertheless, even though follow-up time would need to be started later for these individuals, it would still be follow-up time from individuals who have not been exposed at all to the intervention, meaning concerns about attrition bias, postexposure intervention-related effects, and depletion-of-susceptible-individuals effects would potentially be completely avoided.

The ultra-ultra-briefly-exposed postexposure (ubePE) risk metric is also expected to have pronounced susceptibility to random error in circumstances when this subsample is particularly small. Unfortunately, unlike the assigned-but-not-exposed (abNE) risk metric, since some exposure has occurred, nonconfounding influences of attrition bias, postexposure intervention-related effects, and depletion-of susceptible individual effects can potentially affect the ultra-briefly-exposed postexposure (ubePE) risk metric. However, in many circumstances, some or all of these effects could be expected to be quite minimal or even negligible.

**Supplement 8. Methods used in the real-world data demonstrations of the briefly-exposed postexposure (bePE) risk metric**

This Supplement provides details of the methods employed for the analyses presented in the manuscript.

Derivation of Sample

All individuals receiving care from the Veterans Health Administration (VA) nationwide who were recipients of VA-prescribed lithium or valproate from October 1, 1998 – July 31, 2020 (and who have data recorded in the VA Corporate Data Warehouse) were identified. Follow-up time continued until December 31, 2020. Both medications are mood stabilizers used for treating bipolar disorder, with generally similar monitoring requirements (done through periodically obtaining blood samples to quantify serum levels of the medications). Thus, these medications likely have similar frequency of follow-up visits and clinical reassessment.

The study excluded individuals who were not “first new users” of the medications in the Veteran Affairs (VA) health system as well as those individuals for which there was clear evidence suggesting prior use of the study medications outside the VA system. We use the term “first new users” to clarify that we are not simply defining “new users,” but also not allowing patients to become re-eligible for admission into our study cohort if they do not fully qualify for admission at the time of their first use of either study medication. At the time of their first recorded VA use of either lithium or valproate, individuals either qualified for our cohort or did not, and if they did not, they were not able to enter the cohort later. We view this approach as maximizing the advantages of a “new user” design^26^ by helping to exclude, to the greatest extent practical, individuals who had prior experience with either medication that might influence the selection of what medication they subsequently received. (Any prior use could plausibly increase the likelihood of a re-trial on that medication if the medication was found to be it acceptable or effective. Alternatively, prior use could increase the likelihood of a trial on the alternative medication if the first of the study medications tried was found to be ineffective or associated with undesirable adverse effects). For this reason as well, as described later in this Supplement, individuals’ follow-up time was no longer evaluated and included in the analysis after the point in which they resumed the study medication they had been receiving after the medication had been stopped, or at any point once they switched study medications. Continuing to evaluate follow-up time after these prescription events would in our view, constitute a “second trial” of the study medications and allowing this follow-up time to contribute to the analysis could bias the analysis (through knowledge of efficacy and acceptability of the medication just recently used).

To put it in simpler terms, recipients of the medications were only eligible to enter an intervention arm at the time of their first VA prescription of either medication. Individuals were excluded, however, if they had had prior prescriptions of either medication in their linked Medicare/Medicaid data (for those patients which used Medicare or Medicaid). The term “first new user” is used to clarify that if individual’s first use of either of the study medications, lithium or valproate, did not meet criteria to allow their inclusions [e.g., they did not have a long enough prior history of VA service use at this point] they were not allowed to qualify at a later point even if they had not used either medication for several years. This is a particularly stringent “new user” definition, but one which we believe maximizes the advantages of a new user design, since it minimizes or eliminates *any* prior use of the studied medications.

The study also excluded individuals who previously received a serum blood level lab test for either lithium or valproate as recorded in their VA data. (Having received such a blood test in the past would suggest the possibility that they had been receiving lithium or valproate, even in the absence of a prescription for one of these medications being recorded in the VA or Medicare/Medicaid systems.) Individuals receiving lithium or valproate for investigational purposes were excluded due to uncertainty concerning whether (in some study designs) they might have received a placebo instead of active medication. Our eligibility criteria also excluded individuals initiating both medications on the same day, although no patients actually met this criterion.

To help increase the potential to detect likely prior use of either of these two medications (and especially to help detect ongoing use of either medication that could incorrectly appear to be “new use” at the point an individual transferred their prescriptions to the VA), individuals were required to have used the VA pharmacy to obtain mental health medications at least twice in the two years prior to starting lithium or valproate. To be specific, prior use of the VA pharmacy to obtain mental health medications was required to have occurred once in the year prior to new start (specifically, in days 31-365 prior to new start) and at least once in the preceding year (days 366-730). These criteria established, at a minimum, some pattern of use of the VA pharmacy for mental health purposes for at least the previous 366 days, and in most cases for a longer period, prior to starting either lithium or valproate.

Once potential “first new users” in the VA of lithium and valproate were identified, then diagnostic inclusion criteria were applied. Specifically, cohort members were required to have diagnoses codes recorded in data from their electronic VA medical record for a mood disorder (major depression or depression unspecified, bipolar disorder or other cyclic mood disorders, or schizoaffective disorder) that had been entered into their record on day 0 (i.e., their new start date) or in the prior 28 days. For the purposes of this study, the following codes were used to identify mood disorder diagnoses:

ICD-9 Codes: 295.7, 296.0, 296.00 – 296.06, 296.1, 296.10-296.16, 296.2, 296.20-296.23, 296.25, 296.26; 296.3, 296.30-296.33, 296.35, 296.36, 296.4, 296.40-296.46, 296.50-296.56, 296.6, 296.60-296.66, 296.7, 296.8, 296.80-296.82,296.89, 296.9, 296.90, 296.99, 300.4, 301.13, 311.

ICD-10 Codes: F25.x, F30.1, F30.10-F30.13, F30.2, F30.3, F30.4, F30.8, F30.9, F31.0, F31.10-F31.13, F31.2, F31.30-F31.32, F31.4, F31.5, F31.60 – F31.64, F31.7, F31.70-F31.78, F31.81, F31.89, F31.9, F31.90, F32.0, F32.1, F32.2, F32.4, F32.5, F32.8, F32.89, F32.9, F33.x, F33.0, F33.1, F33.2, F33.4, F33.40, F33.41, F33.42, F33.8, F33.9, F34.x, F34.0, F34.1, F34.8, F34.89, F34.9, F39.

The above codes include diagnostic codes for schizoaffective disorder, in addition to bipolar disorders. Both lithium and valproate are acceptable treatments for at least some forms of schizoaffective disorder. We decided to include these diagnostic codes because not only are lithium and valproate acceptable treatments, but they might be particularly indicated (along with other mood stabilizing medications) for the treatment of “schizoaffective disorder, bipolar type,” one of the two recognized types of schizoaffective disorder (along with schizoaffective disorder, depressed type). Unfortunately, the International Classification of Diseases, 9^th^ Revision (ICD-9), the coding schema used during the enrollment of the majority of our cohort has only a single code for schizoaffective disorder. ICD-9 thus does not discriminate between schizoaffective disorder, bipolar type and schizoaffective disorder, depressed type. Therefore, we chose to view the prescription of lithium or valproate to an individual with the code 295.7 for schizoaffective disorder as suggestive evidence that the individual either has a diagnosis of schizoaffective disorder, bipolar type, or is thought to possibly have the condition to a degree to warrant a trial on lithium or valproate. International Classification of Diseases, 10^th^ Revision (ICD-10) codes have more information, but for consistency we imposed the same approach using these codes, allowing individuals with any type of current schizoaffective diagnosis into the cohort who were also starting lithium or valproate.

In addition to including individuals with schizoaffective disorder in our basic inclusion criteria, we did not exclude individuals receiving a diagnosis of another primary psychotic disorder (Schizophrenia, Psychosis NOS, Brief Psychotic Illness) in the past 28 days, if these individuals also received one of our inclusion criteria diagnoses during that period.

To limit the possibility that individuals were receiving the lithium or valproate for another mental health condition besides a mood disorder, but who happened to have a mood disorder diagnosis as well, we excluded individuals with the following additional VA diagnostic codes indicating past presence at any time of certain other persistent mental disorders (ICD-9 Codes: 293.x, 294.8, 294.9x; ICD-10 Codes: F01.x-F07.x) and ICD-10 codes for Developmental Disabilities, Pervasive Developmental Disorders, or Autism (ICD-10: F70.x, F71.x, F88.x, F84.9, F84.8, F84.0).

We applied additional exclusions based on VA diagnostic, healthcare utilization codes, or laboratory findings demonstrating that the patients had evidence of suffering from a variety of medical conditions or states that might strongly influence prescribing of one or the other treatment, especially in a manner that might introduce confounding. Specifically, since valproate can be used to prevent seizures in addition to treating mood or psychiatric symptoms, individuals were excluded if they had a lifetime history of an active seizure disorder (ICD-9 Code: 354.x; ICD-10 Code: G40.x), or had evidence of conditions thought to increase the likelihood that any newly-started valproate might be being used to treat seizures or as seizure prophylaxis (Skull Fracture or Subdural or Epidural Hematoma [ICD-9 Codes: 432.1, 800.x-804.x, 852.41, 852.21, 852.22, 852.42, 852.43, 907.0; ICD-10 Codes: S06.2, S06.5, I62.00, S06.4], Primary or Metastatic Brain Cancer [ICD-9 Codes: 191.x, 198.3, 198.4, 225.0, 239.6; ICD-10 Codes: C71.x, D33.0, D33.1, D33.2, D33.9, D49.6, and C79.3, C79.31, C79.32], Stroke/Cerebrovascular Accident [ICD-9 Codes: V12.54, 431.x; ICD-10 Codes: Z86.73, I61.9], Hemangioma [ICD-9 Codes: 228.02; ICD-10 Code: D18.02] and Dementia [ICD-9 Codes: 290.x, 290.0, 290.1, 290.2, 291.1, 292.82, 294.1x, 294.2, 331.0, 331.1, 331.19, 331.82, 333.4; ICD-10 Codes: G30.x, F01.x, F02.x, F03.x, F10.97, F13.27, F13.97, F18.17, F18.27, F10.27, F18.97, F19.17, F19.27, F19.97, G10, G31.1, G31.83, S09.8XXA]). Individuals with Traumatic Brain Injury in the last 730 days were also excluded (ICD-9 Codes: 854.01-854.04, 854.06, 907.0; ICD-10 Codes: S06.2, or S06.3, or clinic “stop codes” that represent clinics specialized in traumatic brain injury care [VA primary or secondary clinic stop codes of 197 or 198]).

We also excluded people who were acutely hospitalized in the VA in a non-mental health setting at the time that lithium or valproate was initiated, to decrease the likelihood that valproate recipients in particular might have been receiving the medication for seizures or seizure prophylaxis or for agitation. Similarly, Veterans who were housed in a setting (Nursing Homes or Hospice) that increased the likelihood of these conditions or dementia were also excluded through VA primary and secondary clinic stop codes (VA clinic stop codes 351, 651, 652, 656).

In addition, because some literature exists supporting the use of lithium and anticonvulsants for treatment of cluster headaches and migraine headaches, respectively we excluded anyone with an ICD-9 diagnosis of a cluster or migraine headache in the past 30 days (ICD-9 Codes: 346.x, 339.0, 339.2, 339.4, 339.8) or, for ICD-10 diagnoses, a diagnosis of any headache in the past 30 days (G44.0, G44.2, G44.4, G44.8), due to uncertainties about whether these specific headache conditions would always be reliably coded.

Since lithium and valproate can influence renal or liver function, respectively, and this tendency could plausibly affect prescribing decisions and mortality, we also excluded all lithium and valproate recipients with a baseline estimated Glomerular Filtration Rate (eGFR) < 60 or alanine aminotransferase (ALT) ≥ 120 at their most recent measurement, if present, in the 2 years prior to initial use. In addition, we excluded individuals with the following diagnoses or other indicators of renal impairment:

Presence of diagnoses of Chronic Kidney Disease (CKD) Stage 3, 4, 5, unspecified, or End Stage Renal Disease (ESRD) (ICD-9 Codes: 580.8, 580.81, 580.89, 580.9, 583.81, 583.89, 585.3, 585.4, 585.5, 585.6, 585.9, 639.3, 995.9x (except not 995.93), O38.x; ICD-10 Codes: N18.3, N18.4, N18.5, N18.6).

In addition, we excluded patients with severe Hypertension-related CKD (ICD-9 Codes: 403.01, 403.11, 403.91; ICD-10 Code: I12.0), and other diagnostic codes (from the Elixhauser Renal Failure-Severe Comorbidity Category, which includes the following additional codes ICD-9 Codes: V56.1, V56.2, V56.31, V45.12, V42.0, V45.11; ICD-10 Codes: Z49.01, Z49.02, Z49.31, Z49.32, Z91.15, Z99.2, Z94.0, I13.2, I131.1, I12.0).

Also excluded were patients with nephrotic levels of proteinuria (Urinary Protein ≥3500 or Protein Nonquantitative Dipstick = 5 or Urinary Albumin ≥ 1968 or Urinary Albumin/Creatinine Ratio$\geq$ 1968) or estimated Glomerular Filtration Rate (eGFR) of < 60 (based on the 2021 CKD-EPI equation) at their most recent measurement (within the last two years) prior to medication initiation. In addition, individuals with any diagnosis of Acute Kidney Injury Acute Kidney Injury (ICD-9 Codes: 584.x, ICD-10 Code N17.x, N19.x) in the last 180 days were excluded.

Finally, individuals were excluded if they had diagnoses of Severe Liver Failure (ICD-9 Codes: 456.0x-456.2x, 456.8, 572.2x-572.8x, ICD-10 Codes: B19.0, B19.11, B19.21, I85.00, I85.01, I85.10, I85.11, I86.4, K70.40, K70.41, K72.10, K72.11, K72.90, K72.91, K76.5, K76.6, K76.7, K91.82), or Hepatic Encephalopathy (ICD-9 Code: 572.2, ICD-10 Code: K72.90).

After these inclusion and exclusion criteria were applied, our final cohort included a fairly similar proportion individuals who were lithium first new users in the VA compared to all individuals receiving lithium prescriptions from the VA (21,979 / 159,021, or 13.8%) and individuals who were valproate first new users in the VA compared to all individuals receiving any valproate prescriptions from the VA (63,625 / 540,040, or 11.8%).

It should be noted that this same cohort was used as the initial basis for the Weight Gain, Hypertension, and Hyperlipidemia analyses. Additional individuals were excluded from the Hypertension and Hyperlipidemia analysis samples if they had prior diagnoses of these conditions in the past 2 years, and the exact sample size varied based on the trimming and matching of the sample that occurred during the high-dimensional propensity score (hdPS) matching procedures.

Of note, however, all of the other steps of the Weight Gain, Hypertension, and Hyperlipidemia analysis were performed separately for each outcome. This included the process of selecting covariates for inclusion into the propensity score (i.e., variables were included that had an age- and sex-adjusted hazard ratio of > 1.2 or < 1/1.2 for the specific outcome being studied^27^), the derivation of separate propensity scores for each outcome, and separate propensity score matching in addition to separate survival analyses.

Exposure to the Medications of Interest

Exposure was defined as receiving one or more prescriptions of lithium or valproate. Exposure was judged as starting on the release date of the first prescription. Exposure to lithium or valproate was judged to end after the supply of medication as defined by days of prescribed treatment (also termed the “days’ supply”) of the last prescription to be initiated within the days’ supply plus, if needed, any additional days that made up the “allowable gap” period (also termed the “grace period”^28,29^). This “allowable gap”/”grace period” was instituted to take into account the possibility that the exposure may not have started precisely on the release date and/or may have extended longer than the days’ supply because of missed doses or deliberate intermittent adherence. If no prescription started after the first prescription within the days’ supply and allowable gap period, exposure was judged to have ended after the end of the allowable gap for the first prescription. If new prescriptions did occur (as judged by the release date of the prescription) within the days’ supply or allowable gap of the first prescription or subsequent prescriptions, exposure was judged to have ended after the end of the allowable gap of the latest prescription that met these criteria. We also accounted for overlap in supply resulting from new prescriptions being released while supply from the previous prescription still would be expected to still exist (if the medication was used as prescribed). This additional supply was added onto the days supplied by the subsequent prescription (since presumably the supply from the subsequent prescription would not be started to be used until the supply from the earlier prescription was used up completely), and this excess could be carried forward.

Our “allowable gap” / “grace period” definition includes both fixed and variable components to account for system factors that could influence when a patient receives the medication, as well as a variable component intended to reflect patient behaviors that may affect when they needed a refill of medication. Specifically, for the first prescription, to account for the fact that adherence may be lower when a patient is first starting a prescription due to ambivalence about taking the medication, the variable component was set at 40% of the first prescriptions’ “days’ supply” (allowing for 60% adherence during the prescription), and the fixed component was set at 5 days, to reflect possible reluctance to start the medication initially, refill it once it had been expended, or a lag in receiving the medication if it was mailed to the patient rather than picked up at a VA pharmacy. For all subsequent prescriptions, the variable portion of the “allowable gap” after the days’ supply of the medication was restricted to 30% of the prescription’s days’ supply (thus requiring 70% adherence to the medication, a standard of 10% lower than the standard frequently applied in randomized trials of use of 80% or more of prescribed doses. This lower threshold was chosen for this real-world study of everyday use to account for the lower adherence expected to be observed in the less tightly monitored circumstances that exist outside of a randomized trial). For all prescriptions after the first prescription, the fixed gap was restricted to a 3-day gap to allow for possible mailing of the prescription. No “allowable gap”/”grace period” definition is perfect: for example, not all prescriptions are mailed, and for those that are, anecdotally sometimes patients will report a gap of up to 7 days in receiving the prescription.

Medicare/Medicaid data was not used for determining the initial prescription of lithium or valproate, since we did not expect to have the detailed VA covariate information at that date that would be needed to construct a propensity score for that individual. Instead, any Medicare/Medicaid prescriptions that preceded the first VA prescription excluded the individual from the cohort, as mentioned earlier. However, once an individual was identified as being a “first new user” within the VA (and if they qualified to be in our cohort based on our inclusion/exclusion criteria), any Medicare/Medicaid prescription data for the individual was used in conjunction with their VA prescription data to determine the length of the current recipient period. That is, the individual was judged to still be a current recipient of lithium or valproate even if the days’ supply of their VA medication, plus the grace period, was exceeded, as long as a Medicare/Medicaid prescription occurred before the end of the grace period.

Outcome Definitions

All-Cause Mortality: Individuals suffering all-cause mortality were identified from the VA/DoD mortality registry, a comprehensive source of National Death Index information concerning death and cause of death. This source provides information about an individual’s decedent status regardless of whether the individual’s death was noted in their VA health record, even including the deaths of individuals who were no longer receiving VA care.

Hypertension: Individuals suffering new onset Hypertension (as defined by not having a prior VA [or VA CMS] diagnosis in the past 2 years) were identified with ICD-9 codes (401.x or 437.2) or ICD-10 codes (H35.031-.033, H35.039, I10, I11.0, I11.2, I11.9, I12.0, I12.9, I13.10, I13.2, I15.0-15.2, I15.8-15.9, I16.0, I16.1, I16.9, I67.4, I131.1, O10.111-O10.113, O10.119, O10.12, O10.13, O10.02, O10.03, O10.211-O10.213, O10.219, O10.311-O10.313, O10.319, O10.32, O10.33, O10.411-O10.413, O10.419, O10.42, O10.43, O10.911-O10.913, O10.919, O10.92, O10.93, O11.1, O11.1-O11.5, O11.9, O16.1-O16.5, O16.9) codes.

Hyperlipidemia: Individuals suffering new onset hyperlipidemia (as defined by not having a prior VA diagnosis in the past 2 years) were defined by ICD-9 codes (272.0-272.4) or ICD-10 codes (E78.0, E78.01, E78.1-E78.4, E78.41, E78.49, E78.5).

Weight Gain: Weight gain was defined as ≥ 15% weight gain from their most recent baseline weight in the last 2 years for those individuals who had baseline weight recorded in the last two years in data from their VA medical record.

Propensity Score Methods

Propensity scores were derived based largely on the approach of Patrick and coauthors.^27^ From a set of approximately 6200 covariates built to represent indicating and comorbid mental health, substance use, and nonmental health diagnoses and diagnostic categories (e.g., Charlson and Elixhauser Comorbidity categories), general outpatient and inpatient mental health and nonmental health utilization, total number of current and recent mental health and nonmental health prescriptions, total number of specific mental health, substance use, and nonmental health outpatient visit types, mental health, substance use, and nonmental health hospitalizations, a composite indicator of tobacco smoking status, suicidal ideation and suicidal behavior diagnostic codes and VA “Health Factor” information, and nonmental health diagnostic testing, including imaging, PFTs, EKGs, and blood laboratory visits,^30^ as well as laboratory values for 22 different blood tests and vital signs (systolic blood pressure, diastolic blood pressure, pulse, and weight).

For most baseline covariates that were considered for inclusion into the propensity score, missing values were incorporated as zeroes, since the available data did not provide evidence that that condition was present. This procedure was used for indicators of individual medications and classes of medications, med classes and diagnoses, medical procedures, outpatient visits and inpatient stays, clinic stops, bed sections, and data from clinical assessments. For example, suicidal behavior was separated into various time-based windows (according to the number of days prior to new start) and was valued as 1 if a given diagnosis or indicator of suicidal behavior was present, and a 0 if it was not. Laboratory and vital sign data (blood pressure, weight, etc.) and age were coded in deciles with an additional category for missing data.

Covariates were selected to be included in the propensity score model if they met two criteria: 1) the covariate had a prevalence of ≥100 individuals out of the combined sample^31^ of lithium and valproate new users (85,604 individuals), and 2) the covariate showed a 20% or greater association with the outcome, as defined by exhibiting a univariate hazard ratio of > 1.2 or < 1/1.2 in a Cox regression model that included terms for age in years, age^2^, and sex.^27^ The hazard ratio for the outcome was determined among current recipients from the larger of the two intervention group (the valproate group) over the first 365 days of follow-up. Variable selection was done in this way so as to eliminate or minimize the potential to select variables without a direct effect on the outcome but with an association with the outcome that was due to their association with initiating or adhering to either medication, if that medication had an actual influence on the outcome risk.

Some variables related to age, gender, race and ethnicity at new start were included in all propensity score models regardless of their univariate association with outcome, along with variables reflecting Elixhauser (per the Agency for Healthcare Quality and Research software) or Charlson comorbidity categories, the overall weighted and unweighted Elixhauser Comorbidity score, and estimated GFR (eGFR) at baseline.

Once covariates were identified, a propensity score was generated using logistic regression. This step also resulted in some variables with very low counts being excluded, sometimes because their inclusion would be redundant given other variables that were included in the model.

The all-cause mortality model contained 2,274 variables and had a c statistic of 0.733. The Weight Gain, Hypertension, and Hyperlipidemia models contained 1,956, 1,224, and 1,037 variables, respectively and had c statistics of 0.721, 0.696, and 0.695, respectively.

Individuals with a propensity score (PS) falling outside the “Common Support Area” (i.e., the area in which the propensity scores for individuals initiating lithium or valproate overlapped) were excluded. From within the retained individuals, greedy nearest-neighbor 1:1 matching of lithium to valproate recipients was conducted, with the order of matching of lithium recipients being determined randomly using the SAS PS MATCH program from SAS GRID 8.3 (Cary, North Carolina). All pairs that were able to be matched within calipers of two-tenths of the propensity score logit (i.e., 0.2 * PS logit) were included.

For the all-cause mortality analysis, out of 21,979 individuals initiating lithium, three lithium recipients were excluded for being outside the propensity score Common Support Area, and out of 63625 valproate recipients, 130 valproate recipients were excluded for being outside of the Common Support Area. The 1:1 propensity score matching resulted in 20,139 lithium and 20,139 valproate recipients being in the final matched cohort, a 91.6% match of our final first new user cohort of 21,979 individuals initiating lithium.

Similarly, the same procedures (including trimming to the common support area and 1:1 greedy next neighbor matching using calipers of 0.2*the PS logit) were done for the weight gain, hypertension, and Hyperlipidemia analyses. The starting samples that existed prior to matching were smaller for each of these outcomes than for the all-cause mortality analysis because individuals were excluded from the initial sample if they had prior occurrence of the outcome within 2 years of new start (since the intention was to evaluate the association of lithium and valproate with the new onset of particular medically-significant adverse events) or, for the weight gain outcome, if they lacked baseline data. These restrictions reduced the initial new user cohort meeting all eligibility criteria to 21,096 lithium recipients for the Weight Gain outcome, 13,132 lithium recipients for the Hypertension outcome, and 11,198 lithium recipients for the Hyperlipidemia outcome. After trimming to the common support area and propensity score matching, the cohorts for these outcomes were further reduced to 19,480, 12,383, and 10,568 individuals per treatment arm, respectively, which represented 92.3, 94.3, and 94.4 percent matches of lithium recipients when compared to the first new user cohort meeting all eligibility criteria (including no prior occurrence of the condition).

Most covariates in the propensity scores had rather small initial standardized differences prior to matching, and these differences became considerably smaller after propensity score matching. For example, for the mortality analysis the average standardized difference between the lithium and valproate treatment groups across all included covariates prior to matching was 2.05%. After matching, the average standardized difference narrowed further to 0.44% (approximately a 4.5-fold reduction in average standardized difference). An even greater change after matching occurred among covariates with larger standardized differences . Prior to propensity score matching, the largest standardized difference in the mortality analysis was 25.2% while after propensity score-matching, the largest standardized difference was 1.80% (a 14-fold reduction). For the three adverse event outcomes, the maximum standardized difference for any covariate included in the model after propensity score matching was 1.99% (Weight Gain), 2.50% (Hypertension), and 2.57% (Hyperlipidemia).

Definition of Current Recipient and various forms of Intent-To-Treat Follow-up Time Measures

For our modified intent-to-treat analyses (termed here as an “additionally-censored” intent-to-treat analyses), follow-up time was censored upon the standard criteria of the death of the individual (which also counted as the outcome for the all-cause mortality analysis), or if the end of the study period of interest was reached (e.g., 365, 730, or 1,825 days), or if the end date for the study was reached (December 31, 2020). In addition, follow-up time for all participants were censored upon switching to the other study medication (e.g., valproate if they had been started on lithium, and vice versa), with the date of censoring being determined by the release date of the other study medication, or upon resumption of their original medication if they had previously exceeded the allowable period (i.e., upon the release date of another prescription for the original study medication if the release date occurred after the allowable gap period had been exceeded). Censoring upon resumption was done, even though the study medication was the same, because this follow-up time was viewed as no longer representing a “new user” trial of that study medication. Such patients had already received the medication earlier, and had met our definition for stopping the medication, thus ending that earlier, “first new user” trial of the medication.

If these were the only censoring conditions, these analyses, as described in the manuscript were termed “modified” intent-to-treat analyses. (In this supplement, we will also use the more descriptive term “additionally-censored” intent-to-treat analyses for the same analyses, as described below and in Supplement 9). These analyses were identified as a form of “intent-to-treat” analyses since patients were followed regardless of whether or not they were still receiving the study intervention, a major principle of intent-to-treat analyses. However, we use the qualifying term “additionally-censored” because individuals were not allowed to continue to be in the cohort if they switched to the other study medication at any time or resumed their initial study medication after the grace period had been exceeded. Therefore, not all individuals were followed until they either experienced an event or the end of the study period occurred (unlike a traditional intent-to-treat analyses).

Thus, our additionally-censored intent-to-treat analyses do not perfectly match what is typically implemented in randomized trials. (Randomized trials, it should be noted, are often of much shorter duration and typically feature considerably less frequent discontinuation of the intervention than nonrandomized cohort studies.) However, additionally-censored intent-to-treat time does capture completely and exclusively the total of the two other types of follow-up time that were of interest in our study: current recipient and former recipient follow-up times (defined below). (That is, these two subsets of follow-up time do sum exactly to equal the additionally-censored intent-to-treat analysis follow-up time.)

The adverse event analyses (i.e., the analyses of Hypertension, Hyperlipidemia, and ≥ 15% Weight Gain) were completed using current recipient analyses instead of the modified (i.e., additionally-censored) intent-to-treat analyses used for the all-cause mortality analysis. This was done because traditionally current recipient analyses (i.e., analyses of individuals still actively exposed to the intervention or within the allowable “gap” or “grace period”) are usually of primary interest when studying adverse effects from medications. The reason why current recipient analyses are traditionally used for adverse event analyses is because there are concerns that including nonexposed time could reduce any effect and create a misleading impression of a lack of an adverse effect (i.e., an overestimation of an intervention’s safety). Current recipient analyses added one additional censoring criteria to those used for the additionally-censored intent-to-treat analyses: follow-up time was censored once the individual’s initial treatment course was completed. The initial treatment course was judged as completed at the point in which the individual had exceeded all the days’ supply the individual had been prescribed plus the “allowable gap” or “grace period”, as described above in the “Exposure to Medications of Interest” section.

Practically speaking, this meant that no current recipients were censored for resuming the original study medication, because during current recipient time the original treatment course was not yet considered to be completed. Completion of the original treatment course is a precondition for an individual to be judged as having later resumed the original study medication. Put another way, during current recipient follow-up time, any seeming gaps in active treatment fell within the allowable grace period and thus considered to be part of the ongoing current recipient exposure period.

Thus, both the additionally-censored intent-to-treat analyses and the current recipient analyses had five censoring conditions, however, the current recipient analysis lacked a censoring condition that the additionally-censored intent-to-treat analysis had (resumption of the original study medication) and possessed an additional censoring criteria (the end date of the initial treatment course) which the additionally-censored intent-to-treat analysis lacked.

“Time to event” for the adverse event analyses was determined based on the first occurrence during follow-up of the diagnosis that specified the outcome (or, for the weight gain analysis, the first weight measure that was ≥ 15% of the baseline weight).

Definition of the briefly-exposed Postexposure (bePE) Follow-Up Time Measure

To quantify the briefly-exposed postexposure risk, we quantified postexposure time by starting follow-up following the end of the “briefly-exposed current recipient” follow-up time. “Briefly-exposed current recipient” (beCR) follow-up time included the days’ supply the individual received initially (up to a maximum of 30 days’ supply) plus the allowable grace period from the single prescription they received before the allowable grace period was exceeded.

For this study, our definition of who was “briefly exposed” restricted the cohort to only those individuals receiving only the briefest number of prescriptions of lithium or valproate possible (a single inpatient or outpatient prescription), and that prescription had to be for a duration (i.e., days’ supply) of 30 days or less. (This restriction in the duration of the prescription really only impacted outpatient prescriptions, since all inpatient prescriptions were considered a new prescription every single day. Thus, for individuals starting treatment as inpatients, only a single day’s exposure was allowed.) Individuals receiving multiple outpatient prescriptions were excluded from the “briefly exposed” cohort, even if those multiple prescriptions only totaled to 30 days or less of follow-up time.

That is, all briefly-exposed individuals had received, as their entire initial treatment course, at most only a single inpatient or outpatient prescription for either lithium or valproate of 30 days or less. Briefly-exposed postexposure follow-up time then started once the allowable gap period associated with the duration of their initial brief treatment course was exceeded. Briefly-exposed postexposure follow-up time was then continued to the point when any of the censoring criteria for the “additionally-censored” intent-to-treat analyses were met (resumption, switching, death, end of follow-up period, or the end of study data).

Statistical Analyses

Relative risk ratios were used to assess briefly-exposed postexposure (bePE) risks associated with different cohort derivation steps. (NOTE: in the future, using rate ratios for this purpose could be considered and would cause the Study Flowchart briefly-exposed postexposure (bePE) risk to more closely match the unmatched survival analyses findings.) Cox regression was used to determine hazard ratios for individuals throughout follow-up (additionally- censored intent-to-treat analyses) and for those individuals contributing to the briefly-exposed postexposure (bePE) risk metric. In the latter case, as discussed above, follow-up time began at the end of current recipient time, and this start of follow-up could be 47 days after the start of follow-up for the entire cohort (for those patients receiving a single prescription of 30 days duration, plus the 17 day allowed grace period) and continued to the end of the study period, as long as the individual did not resume their medication, switch to the other medication, or reach the end of follow-up.

All statistical analyses were performed using SAS GRID 8.3 (Cary, North Carolina). Calculation of relative Risk Ratios for Table 2 and Quality Assessment values for Table 5 were done using Microsoft Excel (Microsoft Excel for Microsoft 365 MSO 2506 Build 16.0.18925.20216 32-bit).

Survival plots (Kaplan-Meier curves) were inspected visually to assess for whether the proportional hazards assumption was met for the “additionally-censored” intent-to-treat and current recipient analyses. In addition to our primary propensity score matching analyses, “unmatched” analyses were conducted for each time period evaluating the hazard ratio observed in the study period for all cohort members meeting our inclusion/exclusion criteria. These analyses quantified risks observed in this larger unmatched cohort, prior to application of the propensity score (i.e., prior to both trimming to the Common Support Area and conducting propensity score matching).

**Supplement 9.** **Rationale for the use of “modified” or “ additionally-censored” intent-to-treat analysis censoring conditions**

We added censoring conditions to our intent-to-treat analyses, ending follow-up time when individuals resuming the study intervention or switched to the other study medication. These additional censoring conditions created “modified” (or, alternatively, “additionally-censored”) intent-to-treat analyses. This was done because switching or resuming study medication was judged to end the “first new user” medication trial of the study medication.

To elaborate: As mentioned above, we employed a modified intent-to-treat criteria in our study and referred to it as such in our manuscript. However, since the term “modified intent-to-treat” is typically used to denote a sample in which not all individuals who are assigned an intervention are included in the analysis (e.g., in many randomized trials), we use the term “additionally-censored intent-to-treat” analysis in these Supplements instead. Our rationale for these additionally-censored intent-to-treat (acITT) analysis criteria is related to the practice of adopting “new user” designs.^26^ that has been used in many nonrandomized studies of medications for approximately two decades. “New user” designs are intended to avoid biases of providers or patients deliberately choosing to initiate (or deliberately choosing to avoid initiating) a medication previously trialed by the patient. Typically, individuals are included in the study cohort only if there is evidence that they have not used the medication previously, which, for practical purposes, is usually defined as not having a record of receiving the medication within some “clean period” of months or years prior the start date for the intervention. This approach addresses the concern that, if knowledge exists from a previous trial that the medication either provides benefit to the patient or causes no or only limited adverse effects, the providers or patients will be more willing to resume that medication, creating an overly favorable impression of the medications’ efficacy or lack of side effects. Alternatively, providers or patients could also choose not to re-initiate a medication or preferentially switch to the alternative medication if the original medication that was previously started did not have benefit or was not well tolerated, also creating the same type of bias. We enhance this eligibility criteria beyond the criteria sometimes implemented by only allowing individuals to enter the cohort once, with their first use of the medications under study within our database. If they do not meet criteria for a “clean period” at that point, then those individuals are not included in the study cohort (i.e., they cannot enter the cohort later, at a point when they might fulfill “clean period” criteria, and individuals cannot provide data from more than one course of treatment to the study). Because of this additional restriction beyond how a “new user” definition is sometimes implemented, we term this approach a “first new user” approach.

Along these same lines, we view the switching to the other study medication, along with any resumption of the original study medication after the allowable gap period has been exceeded, to constitute a “second medication trial.” As such, we would expect this second medication trial to be subject to the biases discussed above. To avoid allowing this bias to enter our analysis, we simply censor follow-up time at the point of re-initiating the first medication or switching to an alternative medication. For this reason, we term this modification to the approach that is sometimes implemented for intent-to-treat analyses (which sometimes only censors individuals if they reach the end of the study period or experience an outcome event), as an “additionally-censored” intent-to-treat analysis.

**Supplement 10. Alternative definitions of nonexposure periods can be considered that vary based on the stringency with which follow-up time on other interventions are excluded from the metric**

In this initial demonstration of these methods, we adopted perhaps the simplest possible definition of nonexposure: the period of time for which individuals are receiving none of the interventions or exposures being assessed. More stringent nonexposure definitions can be considered that exclude follow-up time during which other interventions that also affect outcome risk are received. These more stringent nonexposure definitions probably have some advantages, as long as adequate power to limit random error still exists.

To elaborate: For the briefly-exposed postexposure [bePE] risk metric used in the manuscript, follow-up time was censored if the end of the follow-up period was reached, or if the original medication was resumed or the alternative medication under study was initiated (i.e., if valproate was started when lithium was originally initiated, or vice versa). The result of these additional censoring conditions is that the nonexposure period as thus defined is free of known exposure to either of the medications under study.

However, it is easy to think of additional restrictions that could be included in the definition of the nonexposure period. These additional restrictions would generally be expected to make the briefly-exposed postexposure (bePE) risk metric an even better approximation of confounding (as long as the increasing potential for random error did not overly affect the estimate). At least four types of increased stringency of the nonexposure period definition can be easily envisioned. We discuss them here in reference to the briefly-exposed postexposure (bePE) risk metric using hypothetical examples based on our weight gain analysis.

For instance, a more stringent (i.e., more restrictive) definition of the postexposure period could include censoring upon the initiation of some other particular medications besides those under study that are thought to influence the risk of the outcome. For our analysis of weight gain, for instance, it would seem likely that initiation of other mental health medications might occur with some frequency for at least some individuals during the briefly-exposed postexposure follow-up period. Some mental health medications are known to have weight-altering potential. Thus, a logical additional censoring condition that could be considered would be to also censor postexposure risk follow-up time for the weight gain analysis upon initiation of any other mental health medication with weight-altering potential.

An even stricter definition of the briefly-exposed postexposure period would include censoring individuals initiating *any* medication with known weight-altering potential during the follow-up period (whether used for mental health or other types of treatment).

A yet stricter definition of the briefly-exposed postexposure period could include censoring individuals upon initiation of any new *treatment* with weight-altering potential (i.e., a medication or a non-medication intervention). Such a non-medication intervention might include a weight management program or an exercise program that would be expected to have weight-altering potential.

A yet still stricter definition could include censoring the follow-up time upon initiation of *any* new medication if one is particularly concerned that prior knowledge of which medications can cause alterations in weight is not sufficiently robust.

Finally, the most stringent or strict nonexposure risk period definition that we can envision would be to censor individuals initiating any new interventions, including any medications or non-medication interventions, with the rationale that some interventions might have downstream effects on weight (e.g., a non-weight focused psychotherapy increasing an individual’s motivation to meet their goals, which might include weight loss or gain).

Obviously, there are tradeoffs between a) the convenience of implementation of the metric, and of particular importance, b) considerations about statistical power and c) the potential for imbalanced frequency of censoring to occur. For instance, in general the more censoring conditions that exist, the greater likelihood that less statistical power for the metric will be present. In addition, if one intervention under study is viewed as even slightly more of an “end stage” or “last resort” intervention, then this circumstance may affect the likelihood that individuals will initiate other treatments after discontinuing their original medication under study. This could lead to some biases being introduced by censoring. All of these tradeoffs around the censoring considerations for the briefly-exposed postexposure (bePE) risk metric could warrant consideration when deciding the stringency with which the nonexposure period is defined. It is likely that decisions about briefly-exposed postexposure (bePE) risk metric stringency will vary on a case-by-case basis. That said, we would expect future validation studies to show that, in general, as long as statistical power is sufficient, stricter nonexposure period definitions are more likely, on average, to result in briefly-exposed postexposure (bePE) risk estimates that even more closely approximate actual confounding than the simpler approach we took in this initial manuscript. This expectation may not be universally true, however, and this question of how to best define nonexposure periods warrants further research.

**Supplement 11. Clarification of the purpose of using a nonexposure risk metric during cohort derivation**

As an important clarification, one should not necessarily expect every step of a cohort derivation to lead to less confounded effect estimates (as approximated by a nonexposure risk metric). Some cohort derivation steps are traditionally performed to refine the cohort based on other considerations than simply addressing confounding. Nevertheless, observing that a cohort derivation step appears to increase, rather than decrease, confounding, may prompt further useful consideration of whether that step in the cohort derivation is truly needed or desirable.

To elaborate: We wish to note that the purpose of using a nonexposure risk metric to inform cohort derivation is not to attempt to be able to observe the nonexposure risk metric progressively better approximate the null (i.e., progressively converge on a null value) at every step of the process. Rather, it is simply a tool to help one to assess whether the cohort derivation step is functioning as you expected, and, if not, to help inform whether that deviation from expectations at that point should be a substantial concern. Some steps, for instance, are important standard practice to better define the individuals for whom the study’s effect estimates will apply, especially if that definition can help prevent biases, such as biases from prior knowledge. For instance, the well-established cohort derivation step of removing individuals who are not “new users”^26^ of an intervention is intended to prevent biases from prior experience with the interventions from influencing individual-level intervention choice and thus effect estimates.

In our view, the actual degree to which the cohort derivation steps create two study cohorts with nonexposure risk metrics that come close to approximating the null is of greatest interest *for the final step in the derivation.* For the other steps, if nonexposure risk metrics such as the briefly-exposed postexposure (bePE) risk metric suggests that a particular step appears to not potentially reduce confounding, then these metrics can simply serve to “flag” that step for potential further examination. Specifically, those steps that do not produce more comparable briefly-exposed postexposure (bePE) risks between the intervention arms may benefit from reexamination about whether they are really needed. However, those steps that do not appear to clearly reduce confounding should not be excluded from a cohort derivation process if there are other valid reasons to include them.

For instance, the study cohort derivation we presented in the manuscript actually represents the “collapsing” of information obtained from performing the cohort derivation as a larger series of steps. We did this to simplify our presentation for readers so that the main point that the briefly-exposed postexposure (bePE) risk metric can assist in tracking changes in potential confounding was not obscured. Not every one of the detailed steps led to a briefly-exposed postexposure (bePE) risk closer to the null. For instance, in one of these more detailed steps we excluded individuals with serum lab levels for lithium or valproate that preceded their VA lithium or valproate use. This exclusion was based on the assumption that, in general, individuals would not be receiving those labs in the absence of a VA prescription for these medications unless they were already receiving lithium or valproate from a non-VA source. For whatever reason, excluding first VA users of the study medication who had experienced prior serum lab testing for lithium or valproate from our study cohort actually increased the divergence of the briefly-exposed postexposure (bePE) risk from the null in the remaining cohort very slightly compared to the previous step. (That is, the briefly-exposed postexposure (bePE) relative risk actually decreased slightly at that step, creating a ratio further from the null.) However, we certainly wanted to avoid including people in our cohort who could have been past users of lithium or valproate, and so we felt that this serum lab exclusion was well-justified for reasons separate from confounding. It is unclear why this increased divergence from the null for the briefly-exposed postexposure (bePE) risk metric is occurring at this step, or whether these increased divergences from the null have any significant importance.

If this divergence further from the null had occurred in the final step of the cohort derivation, then it might have warranted more detailed consideration. This consideration would center on whether the rationale for that step was sufficiently strong to retain it in the cohort derivation procedure despite the possibility it might actually lead to an increase in confounding between the two intervention arms (although other explanations, such as random error, are also possible). Further research will be needed to determine if the briefly-exposed postexposure (bePE) risk can be used to make judgments about confounding based on such potential fine distinctions (such as the minor changes in the briefly-exposed postexposure (bePE) risk metric that might accompany the final step in a cohort derivation process).

In summary, we wish to make the point that currently, in our judgment, it is not necessary for study cohort derivation to proceed in a “lock step” fashion whereby each step is accompanied by progressively diminishing briefly-exposed postexposure (bePE) risk differences between the intervention arms. However, there may be value to reconsidering certain contemplated restrictions when they are expected to reduce confounding but results from nonexposure risk metrics such as the briefly-exposed postexposure (bePE) risk metric actually suggest that this confounding reduction may not be occurring.

Incidentally, we also think it is important to note that seemingly there is no reason why this basic approach to cohort derivation could not be employed *using some other approaches to approximating confounding* as well, such as negative control outcomes. We hypothesize that, in theory, there is no reason why a genuine “negative control outcome” (i.e., one that is genuinely completely unaffected by the interventions/exposures under study) should not show the same basic pattern during the cohort derivation steps that we have shown for briefly-exposed postexposure (bePE) risk metric. If the briefly-exposed postexposure (bePE) risk is indeed approximating the total confounding (i.e., both measured and unmeasured confounding) that still exists at that stage of cohort derivation, it would seem like other valid indicators of confounding would exhibit the same basic patterns of changing risk. That is, we might expect for the study described in the manuscript that a genuine negative control outcome would show a similar pattern of reduction in the differences in risk observed between the intervention arms for the negative control outcome as the cohort is being defined. In specific, we would expect a sizable difference in risk for a valid negative control outcome to be associated with the two intervention arms at the relatively unselected samples of individuals typically occurring early in the cohort derivation process. We also might expect that the differing risk between intervention arms for a valid negative control outcome would progressively diminish in later stages of the cohort derivation process. We are not sure whether negative control outcomes (or other approaches to approximating confounding) have been used in this fashion to help guide cohort derivation. If not, we suggest that the topic of using other confounding indicators such as negative control outcomes to inform cohort derivation in a manner similar to what we have described in the manuscript for the briefly-exposed postexposure (bePE) risk metric be pursued in future research. This discussion is speculative, of course, and the subject area of using tools to estimate confounding to help guide cohort derivation (or, at a minimum, track the potential changes in confounding that result from the process), whether the briefly-exposed postexposure (bePE) risk metric or other approaches, could certainly benefit from further investigation.

**Supplement 12. Further consideration of why using the briefly-exposed postexposure (bePE) risk metrics to aid cohort derivation is not expected to allow additional biases to affect the main analyses**

As discussed extensively in the manuscript and these Supplements, we expect that the briefly-exposed postexposure (bePE) risk metric will typically serve as an approximation of confounding rather than a completely accurate estimate of confounding. Beyond that potential for at least some degree of imprecision, however, we don’t expect using the briefly-exposed postexposure (bePE) metric as an aid during cohort derivation will allow overt biases or prior beliefs on the part of the investigator to affect cohort derivation. The expectation when using the briefly-exposed postexposure (bePE) risk metric is, if the metric does effectively approximate confounding, that nonexposure risks that more closely approximate the null will represent less confounded analyses. Importantly, this expectation for the briefly-exposed postexposure (bePE) risk metric exists regardless of any expectations about the presence or magnitude of any intervention effect during active intervention exposure. Put another way, information about active intervention effects is not examined in this briefly-exposed postexposure (bePE) risk metric-informed cohort derivation process. Thus, information about active intervention effects generally does not inform or bias the cohort derivation process.

To elaborate: When the main analyses investigated in a study are current recipient analyses, then the briefly-exposed postexposure (bePE) risk metric does not use any information that is included in the main effect estimate. Therefore, in our view, using the metric to help guide the cohort derivation process should not bias the analysis in any way outside of potential imprecision in the metric itself as an approximation of confounding. This imprecision can be the result of either random error or other nonconfounding influences. To be more specific, use of the briefly-exposed postexposure (bePE) risk metric does not allow investigator’s expectations of what might be the “proper,” “likely,” or “expected” effect of the intervention to affect the analysis of interest. This is because the briefly-exposed postexposure risk period being examined is entirely external to the current recipient exposure period. There is no need to consider current recipient risks when deriving the cohort, or, indeed, any reason to, since the briefly-exposed postexposure (bePE) risk metric does not rely on such information.

The situation is slightly more nuanced when considering analyses in which the main analysis is evaluated using some form of an intent-to-treat approach. In our view, the use of the briefly-exposed postexposure (bePE) metric also does not risk biasing intent-to-treat main analyses, but for slightly less clearcut reasons. For various forms of intent-to-treat analyses, the briefly-exposed postexposure (bePE) metric *does* in fact include a fraction of the nonexposed follow-up time that is included in the main analysis results (i.e., the intent-to-treat effect estimate for the main cohort). So, for intent-to-treat analyses, as opposed to current recipient analyses, the briefly-exposed postexposure (bePE) risk metric does not quantify risks that are completely separate from what is being evaluated in the main effect estimate. However, in the ideal case in which the briefly-exposed postexposure (bePE) risk metric is only reflecting confounding and not the other nonconfounding influences which can affect nonexposure risks, then using the briefly-exposed postexposure (bePE) risk metric during cohort derivation would simply be expected to help lead to less confounded risks being observed during nonexposure time (and hopefully, through assisting in cohort derivation, during exposure time as well). This lack of confounding in the nonexposure period that is included in the intent-to-treat analysis would only improve the analysis (again, in the ideal case in which the briefly-exposed postexposure (bePE) risk is solely approximating confounding). A cleaner, more valid “signal” of the active treatment effects would be expected to emerge from an unconfounded or close-to-unconfounded intent-to-treat analysis. That is, to the extent that investigators’ cohort derivation efforts would be minimizing the risk difference between intervention arms for some portion of follow-up, it would be minimizing the risk difference occurring during a time period in which is not the primary focus of the analysis, and this minimization would allow the active intervention effects that are the focus to be observed more clearly.

This discussion does reinforce the importance, when using the briefly-exposed postexposure (bePE) risk metric, of assessing the metric quantitatively or qualitatively using “quality assessments.” These quality assessments can provide at least some sense of how likely the risk metric is to be approximating confounding relatively free from other influences.

**Supplement 13. Additional quality assessments and other measures that could be considered to assess nonexposure risk metrics**

A considerable number of additional quality assessments can be readily envisioned, which is important because the stand-alone value of any specific single quality assessment metric is uncertain. In addition, consideration should be given to the use of composite measures as well as a possible role for machine learning or other automated approaches in efforts to judge nonexposure risk metric quality.

To elaborate: A variety of additional quality assessments can be readily envisioned, ranging from progressive refinements of already proposed quality assessments to entirely new assessments measuring different aspects of the performance of the briefly-exposed postexposure (bePE) risk metric. In this supplement we touch upon at least four types of additional refinements that could be considered.

First, there are entries in Table 5B of the manuscript which feature within-intervention-arm-quality assessments, such as the beCR/fs1stRx Rate Ratio per Intervention Arm and the beCR/bePE Rate per Intervention Arm quality assessments. For such assessments, our presentation only shows the results for the particular intervention arm that shows the greatest divergence from 1.0. We did this to streamline our presentation to the reader. But more composite or automated quality assessment procedures could consider more quality assessments than could easily be assimilated by a human reader. In this context, findings from both intervention arms for these within-intervention-arm assessments may have some value. For example, it may be the case that there may be at least some value in considering the divergence from 1.0 observed separately in both intervention arms (i.e., even the arm with the lesser degree of difference).

Second, other refinements to existing quality assessments could be more wide-ranging. These could include providing both quality assessments that are less precisely or stringently defined along with quality assessments that are more precisely and stringently defined. This suggestion may seem counterintuitive, with the more rigorous, precise assessments to be universally favored (and perhaps further research will show this to be the case). However, often there is a trade-off in analyses between the rigor of an analysis and its susceptibility to random error. For example, one alternative assessment (noted in Table 4) is a version of the beCR active intervention/Full Sample 1^st^ Rx active intervention (beCR/fs1stRx) Quality Assessment that would alter the comparison group in this Quality Assessment. Instead of using a comparison group that includes the period of active intervention for all individuals receiving their first prescription (the “full sample”), this alternative Quality Assessment would use a comparison group consisting of only the individuals who receive an initial prescription of ≤ 30 days in duration. In addition, a quality assessment could be defined in which this subsample would be carefully selected, if possible, so as to have the same proportion of individuals receiving single day inpatient prescriptions, 7-day outpatient prescriptions, 14-day outpatient prescriptions, 30-day outpatient prescriptions, and other durations of prescriptions that are ≤ 30 days in duration as the briefly-exposed current recipient sample. Put another way, the subsample would feature a subset of individuals receiving their first prescription so that this comparison subsample would more closely have the same or a similar mean number of follow-up days of active intervention per individual as the briefly-exposed postexposure sample. These changes, however, would be expected to result in a quality assessment that has more random error than the quality assessment based on the full sample.

Another change that could be considered and that would add stringency to the Quality Assessments would be to exclude the individuals in the briefly-exposed postexposure (bePE) postexposure subsample from the denominator in any assessments when we are comparing this subsample to either the whole cohort or the set of briefly-exposed postexposure (bePE)-eligible individuals. For simplicity, we did not implement this exclusion in the relevant Quality Assessments presented in the manuscript. However, our inclusion of briefly-exposed postexposure (bePE) individuals into both the briefly-exposed postexposure (bePE) subsample and the overall comparison group would be expected to attenuate differences between these comparison groups and the briefly-exposed postexposure (bePE) sample. It may be found to be more useful to exclude the briefly-exposed postexposure (bePE) individuals from the quality assessment indicator denominator prior to comparing the risks.

Third, there are potentially new quality assessments that may be devised beyond those presented in the manuscript’s Table 5B. As just one example, rather than just examine mean propensity score differences between intervention arms in the briefly-exposed postexposure (bePE) sample versus the full cohort, it might be even more helpful to examine the mean standardized difference between propensity score covariates between the intervention arms (and then between the briefly-exposed postexposure sample and the full cohort). As discussed in Supplement 14, there may not be a direct relationship between the differences in the propensity score difference between intervention arms seen between the briefly-exposed postexposure (bePE) sample and the full cohort and the differences in covariate balance seen in these two samples. Using a “difference in mean standardized difference (in propensity score covariates)” quality assessment might provide information of even more direct value than the difference in mean propensity score difference.

Another additional quality assessment that could be considered, albeit more cautiously, is similar in some ways to the “Directional Consistency” quality assessment. Another form of consistency between the changes in the unmatched to matched main analysis effect estimates and the associated briefly-exposed postexposure (bePE) risk metric would be a “Size Consistency” assessment. This would examine whether the briefly-exposed postexposure (bePE) risk appears to changes to a similar extent that the main effect estimate changes. Care may need to be taken during any attempts to assess “size consistency” to ensure that the samples being compared are highly similar or identical, and possibly that the effect estimate measure is one that is “exchangeable.” Indeed, it is unclear at this time the degree to which, even if random error is minimal, strict size consistency between main analysis changes and briefly-exposed postexposure (bePE) risk changes are to be expected. For instance, authors investigating negative controls have emphasized that “the magnitude of bias due to uncontrolled confounding cannot generally be inferred from the magnitude of a detected… non-null association without extra assumptions based on firm scientific understanding.”^1^ However, at a minimum, it may be significant to note if large differences exist between the changes that are observed in the main analysis effect estimate (e.g., when going from unmatched to matched analyses) and the changes observed in the associated briefly-exposed postexposure risk estimates. In this case, at a minimum there may be at least some value in examining some of the quality assessments we present in our manuscript, or additional quality assessments for the briefly-exposed postexposure (bePE) risk metric yet to be developed, to assess if some obvious nonconfounding influence on the briefly-exposed postexposure (bePE) risk metric seems plausible. In the future, however, research may help determine whether there are circumstances in which at least inexact conclusions can be drawn about whether confounding appears to account for all or most of a main analysis effect estimate, based on the relative size of the main analysis and briefly-exposed postexposure (bePE) risk metric’s divergences from the null.

Finally, there are alternate types of quality assessments, such as those that give simple descriptive information, rather than those actively assessing whether conditions are present that could suggest that there are other influences besides confounding that are affecting a given nonexposure risk metric. For instance, as we will describe further below, various measures of the “extensiveness” of the nonexposure risk metric could be considered for those metrics which assess postexposure risks. For example:

A) Earliest Nonexposure: the absolute earliest point at which the nonexposed risk metric starts for the individual with the longest nonexposed risk period in the cohort. Before this point in time, a postexposure risk metric cannot estimate anything quantitatively or semi-quantitatively about confounding during that particular time period from intervention initiation. However, if this period is brief, reasonable assumptions may be able to be made about how closely the confounding that is not measured may approximate the confounding that is captured in the nonexposure risk metric.

B) Median Nonexposure Initiation: the median starting point that the nonexposed risk metric starts in the briefly-exposed postexposure (bePE) cohort (in terms of days past the intervention start date).

C) Given Percentile of Nonexposure Initiation: the point at which, for example, at least 20% of the individuals included nonexposed risk metric have begun follow-up time. This is obviously a slightly arbitrary benchmark, but it or something similar to it (e.g., 10%, 25%) may serve as a convenient and useful assessment of the period over which the assessment is likely to begin to more effectively approximate confounding.

The above follow-up time benchmarks are likely to be most useful for the briefly-exposed postexposure (bePE) risk metric, since by definition the ultra-briefly-exposed postexposure (ubePE) risk metric typically begins within a few days of initiation, and these considerations do not apply to the baseline preexposure (basePreE) risk metric, which uses a measure that does not quantify risks observed in the postexposure period at all.

Similarly, and especially for the briefly-exposed postexposure (bePE) risk metric, it may be helpful to summarize the mean amount of time individuals are potentially exposed to the intervention before the postexposure period starts. In our study, for instance, patients were allowed to receive outpatient prescriptions for up to 30 days of duration. Determining the exact mean duration of treatment may be valuable (as we do in Supplement 5), with briefer periods of treatment being desirable.

Although not easily measurable, it is worth reflecting that an uncertain fraction of individuals in the postexposure risk metrics subsamples is likely completely unexposed. In a medication study, this unexposed fraction would include those individuals that picked up their prescription but later indicated that they never actually started taking the medication. Indeed, this fraction of individuals would also include individuals who did not actually start taking the medication whether or not they ever disclosed this information. This is in mild contrast to those individuals assessed for the assigned-but-not-exposed (abNE) risk metric. Those assigned-but-not-exposed individuals would be those for whom some objective evidence suggests that they never started the intervention (at least from the VA system). One scenario that we can conceive fulfilling these criteria is when individuals are issued a prescription but there is an actual record that they never actually received the prescription, such as a record indicating that they never picked the prescription up at the pharmacy.

The “unrecognized” unexposed individuals who are not known to be unexposed until their later self-report, or who are never known to be unexposed (but are in fact unexposed) would not be affected by several nonconfounding influences that can interfere with the use of postexposure risks to approximate confounding. Specifically, they would not be affected by postexposure intervention-related effects or depletion-of-susceptible-individuals effects. In addition, their attrition out of the exposed sample would not be due to actual effects of the intervention (although it still could be affected by the perceived likelihood of actual effects). As such, although difficult to identify specifically, this is a valuable subsample to have constituting some of the postexposure risk subsample, since these individuals are less susceptible to these nonconfounding influences. However, it may be impossible to ever know how large a proportion of the postexposure sample is composed of these unexposed individuals.

It is also worth reflecting that to the extent this unexposed fraction is included in the subsample of individuals making up the ultra-briefly-exposed postexposure, briefly-exposed postexposure, and “all discontinuations” postexposure (adPE) risk metrics, in almost all circumstances these individuals would be expected to make up a larger fraction of the total individuals meeting the criteria for the briefly-exposed postexposure (bePE) and ultra-briefly-exposed postexposure (ubePE) metrics than for the “all discontinuations” postexposure (adPE) metric. The “all discontinuations” postexposure (adPE) risk metric would include these genuinely nonexposed individuals, but since it also includes individuals that stop receiving the intervention later during follow-up, these genuinely nonexposed individuals will constitute less of the overall metric’s risk estimate than for the briefly-exposed postexposure (bePE) and ultra-briefly-exposed postexposure (ubePE) risk metrics. This is another advantage of these risk metrics relative to the “all discontinuations” postexposure (adPE) risk metric.

The fact that additional quality assessments can easily be devised is important in the context that no individual quality assessment appears to be definitive. As indicated in the manuscript, an evolution of quality assessments towards an index or indices that combines multiple quality assessments may be advisable; such an index conceivably could include a large number of assessments, especially if some are weighted more than others. The manuscript seeks to establish the value and feasibility of quality assessments, rather than provide any sort of definitive list of what quality assessments should be used. Finally, there may be a role for machine learning or other approaches to rapidly devise what weighting of quality assessments appears the most informative drawing from information across a number of studies. Whether these approaches would allow for a tailored weighting of quality assessments within a particular study at hand is less clear. Conceivably, such specific weighting might be possible if some of the outcomes being assessed within the study already had well-established intervention effects from which to judge if confounding was present and how well the briefly-exposed postexposure risk revealed that confounding. But this is speculation and a potential topic for future research.

Regarding the value of any single quality assessment, unless the quality assessment value observed is particularly extreme, it is difficult to know how conclusively the quality assessment will suggest whether or not the briefly-exposed postexposure (bePE) risk metric is affected by other nonconfounding influences. This is one reason why the development of additional quality assessments is desirable. In the ideal circumstance, multiple quality assessments would exist for each potential nonconfounding influence on nonexposure risk. In this way, consistency between the different quality assessments evaluating the plausibility of potential nonconfounding influence could be assessed. Ultimately, as mentioned, there may be a role for machine learning or other automated approaches that could integrate findings from across multiple quality assessments. A particular ideal circumstance would be if the state of the science around quality assessments evolved to the point that, through repeated validation studies, a range of values from a bank of quality assessments could be used to predict that (not considering random error) a certain set of values would suggest that the briefly-exposed postexposure (bePE) risk metric would be with a range of X to Y in terms of its accuracy approximating confounding. Such an extreme degree of value for nonexposure risk metric quality assessments may be an unattainable goal, and, at a minimum, may not occur for a considerable time in the future. Nevertheless, it may be a worthwhile goal to pursue.

Clearly, work on quality assessments for nonexposure risk metrics is at an extremely early stage. We have presented and discussed them to the extent we have in the manuscript and supplements primarily to simply call attention to the important opportunity that they represent. Specifically, the opportunity that these quality assessments present is that an investigator or reader does not have to simply passively accept information from nonexposure risk metrics. Nor do investigators or readers need to simply dismiss input from the metric due to concerns that other nonconfounding influences could be affecting the metric and interfering with its ability to validly approximate confounding. Rather, quality assessments for nonexposure risk metrics do exist that can be helpful, and further assessments can be developed that we expect to help improve the inferences made using nonexposure risk metrics. We are also exploring the possibility of using other approaches beyond those described here to help assess nonexposure risk metrics.

**Supplement 14. Further discussion of the Hyperlipidemia analysis briefly-exposed postexposure (bePE) risk metric quality assessment findings**

In our adverse event analyses, it is difficult to know whether any of the potential phenomena assessed by the quality assessments are substantially responsible for our Hyperlipidemia analysis briefly-exposed postexposure (bePE) risk metric findings. Specifically, we observed that the value obtained for the briefly-exposed postexposure (bePE) risk metric for the high-dimensional propensity score (hdPS)-matched Hyperlipidemia analysis was not as close to the null as the value obtained for the briefly-exposed postexposure (bePE) risk metric for the unmatched Hyperlipidemia analysis. This finding may suggest nonconfounding influences are affecting the Hyperlipidemia briefly-exposed postexposure (bePE) risk metric(s) However, the possibility also cannot be excluded that, despite our analytic efforts, the Hyperlipidemia hdPS-matched estimate is simply more confounded than the unmatched estimate.

To elaborate: The four quality assessments for which the Hyperlipidemia analysis shows the most extreme values (relative to the analyses of Hypertension and Weight Gain) are:

1) The difference in mean propensity score difference between the full cohort and the briefly-exposed postexposure (bePE) risk metric subsample (measured as an absolute difference);
2) The difference in mean propensity score difference between the full cohort and the briefly-exposed postexposure (bePE) risk metric subsample measured as a ratio of differences (a quality assessment that is closely related to the assessment listed above);
3) The maximum ratio observed from within either of the intervention arms of the briefly-exposed current recipient event rate versus the full sample cohort event rate during the first prescription current recipient period; and
4) The ratio of the ratios observed within each intervention arm of the briefly-exposed postexposure (bePE) sample size versus the number of individuals eligible to become briefly exposed (e.g., the number of individuals receiving a ≤ 30-day initial outpatient prescription or a single inpatient prescription).

The first two of these quality assessments attempt to assess whether differences in the control of confounding from measured factors between the two groups could be influencing the briefly-exposed postexposure (bePE) risk metric ability to approximate confounding. Simply put, the optimal circumstance is for the briefly-exposed postexposure (bePE) risk metric subsample to have the identical difference in propensity score as the whole cohort, since the briefly-exposed postexposure (bePE) risk metric is intended to “stand in” as a surrogate for the whole cohort in terms of approximating confounding. For this quality assessment, the difference in mean propensity score observed between the intervention arms compared to and the difference that was observed for the whole cohort is more than twice as large for the Hyperlipidemia analysis as was observed for either of the other two outcomes. (In fact, the difference was > 3X as much as that observed for the ≥ 15% Weight Gain outcome.) So, other considerations being equal, this quality assessment indicator suggests that less weight perhaps should be put on the briefly-exposed postexposure (bePE) risk metric results derived from the Hyperlipidemia hdPS-matched analysis, since the quality assessment suggests that it is plausible that the metric may not be approximating measured confounding (and possibly unmeasured confounding as well) as well as the briefly-exposed postexposure (bePE) risk metric for the hdPS analyses of the other two adverse event outcomes. But, as pointed out in the manuscript, it is hard to know what level of difference should raise particular concern.

One way to potential gain insight into whether any observed degree of increase in mean propensity score differences in the briefly-exposed postexposure (bePE) sample are of concern, for instance, suggests itself: examining individual covariate-level changes in risk factor prevalence, either for the whole set of propensity score covariates or for some suspected potential major confounders. Without doing such a procedure it would be difficult to judge how much of a change in risk factor prevalence will accompany a given change in propensity score difference between intervention arms. (We have not thought about this question thoroughly, but we surmise it likely relates, in part, to the amount of confounding existing between the two intervention groups before propensity score matching is applied, and to how extreme the differences are in who gets assigned what intervention. To state this another way, it may relate in part to how overlapping the propensity score distributions are between the two intervention arms. This is a question that might have already been more definitively studies, or that might merit more investigation). After looking at the changes in prevalence in individual covariates between the intervention arms, one could consider using the Bross equation or other approaches to provide at least a rough estimate of how much change in confounding might accompany these changes in measured risk factor prevalence. We have not tried this approach, but it might help make inform judgments about the likely impact of a less than 0.005 units of propensity score difference (observed for our Hyperlipidemia analysis) between the briefly-exposed postexposure sample and the full, main analysis cohort. This in turn would help inform judgments about how much weight to give this quality assessment finding when making judgments about how well or how poorly the briefly-exposed postexposure (bePE) risk metric might be representing confounding in the full, main analysis cohort.

Of course, if one were to do this more detailed investigation, another approach might be simply to compare the covariate balance between the full unmatched cohort and the full hdPS-matched cohort instead and possibly use the Bross equation or other approaches for this comparison to calculate whether the hdPS-matching seems to be reducing measured confounding. As mentioned in Supplement 13, this mean standardized difference comparison could be another quality assessment.

Either of these approaches would lead unmeasured confounding unaddressed, but an investigator could also consider inspecting their data for some of the conditions which can make a given amount of unmeasured confounding more problematic. For instance, an investigator could review the propensity score covariates in search of plausible instrumental variables or semi-instrumental variables that may increase bias in the presence of unmeasured covariates,^32^ or examine the c statistic for their propensity scores to determine if certain propensity scores (such as those propensity scores for which the briefly-exposed postexposure risk metric suggest that matching using those propensity scores does not appear to reduce confounding) have notably larger c statistics (which can be problematic).^33^

There may also be simple expectations based on prior knowledge that analyses of certain outcomes are more likely to contain unmeasured confounding (e.g., analyses of some mental health outcomes, or outcomes for which motivation can play a major role) than analyses of other outcomes (e.g., cardiovascular outcomes, if data about the most important well-established risk factors are present in one’s data).

In this manner, as discussed in the manuscript, the briefly-exposed postexposure metric (and possibly some of the quality assessments) might play a valuable role, if resources are limited, in helping to identify which analyses would benefit from more in-depth investigation. This investigation could involve, among other things, delving more into the functioning of the propensity score methods that were applied.

For now, the point remains that among our adverse event analyses, the Hyperlipidemia briefly-exposed postexposure (bePE) risk metric mean propensity score difference (between intervention arms) does not show as close agreement with the mean propensity score difference for the full matched cohort as is observed for the other outcomes (Hypertension and ≥ 15% Weight Gain).

Perhaps unsurprisingly, the Hyperlipidemia analysis also had the most extreme value for the 2^nd^ quality assessment relating to mean propensity score changes observed between the briefly-exposed postexposure (bePE) risk metric and the full cohort, but this time expressed as a ratio of the mean propensity score differences. For this assessment, the changes observed between the Hyperlipidemia analysis and at least the Weight Gain analysis were even more dramatic. For the difference in mean propensity score differences quality assessment, approximately a 3.5-fold difference was observed between the Hyperlipidemia and Weight Gain analyses. When these differences were expressed as a ratio, the ratio of the briefly-exposed postexposure (bePE) risk metric mean propensity score difference between arms was 13.13-fold larger than the difference in the main analysis for the Hyperlipidemia analysis, while for the Weight Gain outcome it was only 1.6-fold larger (an 8.2-fold difference in these ratios). Unfortunately it is not clear currently which version of the mean propensity score quality assessment is more valuable (and they might both have value). Future research might investigate which assessment is more consistently predictive of the amount of standardized difference change in covariate balance resulting from the change in mean propensity score difference. As pointed out for the first assessment, one logical next step to further pursue findings of interest from either of these quality assessments would be to inspect individual covariate changes. As mentioned above, such an inspection could either look for the size of prevalence changes in important covariates or for all of the propensity score covariates (potentially to facilitate an estimate of possible measured confounding changes).

The third quality assessment for which the Hyperlipidemia analysis showed the most extreme values is intended to partially assess whether or not the briefly-exposed postexposure (bePE) risk sample is likely to be particularly nonrepresentative of the rest of the cohort. This metric measures how different are the effects that are observed associated with active treatment in the briefly-exposed postexposure (bePE) cohort compared to the effects observed associated in the full cohort during active treatment during that same period of time. Many potential reasons why differences in these active effects might be observed can be foreseen, ranging from non-specific patient characteristics (impulsivity, past history, etc.) to factors related specifically to the intervention that these individuals are receiving. For instance, individuals with particularly brief exposure to an intervention can often be expected to experience, on average, less efficacy from the intervention than the full cohort (hence a particular willingness to discontinue the intervention, especially if any side effects are occurring).

Presumably, the greater the difference between the outcome rate observed in the briefly-exposed postexposure (bePE) sample during their active intervention period and the rate observed for the overall sample during that same time period, the greater the basis to have concern that the briefly-exposed postexposure (bePE) group might be enriched for individuals who, on average, are particularly likely to have a different inherent event rate than the full sample. As pointed out in the manuscript, such a difference in inherent event rate need not invalidate the briefly-exposed postexposure (bePE) risk metric, if it is equivalently present in both intervention arms. Indeed, this is information conveyed by another quality assessment, the beCR/fs1stRx Ratio of Rate Ratios during active intervention quality assessment. This quality assessment indicates that the rate ratios in this measure observed for the two intervention arms are rather similar. (The ratio of the ratios for this quality assessment for the Hyperlipidemia analysis is only 1.05, implying a fairly close similarity in this assessment as measured in each of the intervention arms. In fact, this ratio is considerably less pronounced than observed for the Hypertension analysis, for which this ratio is 1.31. Thus, it seems less likely that the small differences seen in the event rate between the briefly-exposed sample and the full cohort during the very early time period when the briefly-exposed individuals are receiving the active intervention [and the corresponding time period in the full sample] are substantially responsible for the observation that the hdPS-matched Hyperlipidemia briefly-exposed postexposure (bePE) risk metric is further from the null than the unmatched Hyperlipidemia analysis. We make this judgment because a considerably larger ratio of ratios is observed for the Hypertension analysis, an analysis for which the hdPS-matched briefly-exposed postexposure (bePE) risk metric does in fact exhibit a value that is closer to the null than the briefly-exposed postexposure (bePE) risk metric for the unmatched Hypertension analysis.

However, even if there is not a substantial difference in the briefly-exposed current recipient event rate and that of the full cohort over the corresponding time period between treatment arms, the existence of some degree of discrepancy between these rates within one or both of the treatment arms should probably produce at least some mild concerns that the briefly-exposed postexposure might be not fully representative of the full cohort. However, as we have mentioned in the manuscript and will mention below, currently work on these quality assessments is at such an early stage that it is not possible to know whether or not the ratio of 0.75 observed for one of the treatment arms for this quality assessment should be a basis of particular concern.

The fourth quality assessment mentioned reflects one of the most basic potential indicators of nonrepresentativeness, by assessing whether or not a different percentage of individuals from the full cohort become only briefly exposed. The desired finding is that equal percentages across intervention arms of those individuals who are eligible to become briefly exposed in fact become briefly exposed. In a sense, this quality assessment is similar to tracking attrition in randomized trials, with the assumption being that the more differential the attrition rate that is observed between treatment arms, the greater chance that bias may result. In the case of this quality assessment, the bias would result from the different briefly-exposed postexposure (bePE) samples for the different intervention arms being potentially more and less representative, respectively, of the whole treatment arm that they are representing (by virtue of one intervention arm having a greater proportion of the sample becoming just briefly exposed).

It is also worth stating what might be obvious: no critical mass of research exists to judge when the size of effect observed for these quality assessments is sufficient to raise particular concerns about a finding. (In the strictest sense, one could say the same thing about measures of attrition from a randomized trial.) One of the major reasons for presenting these quality indicators so early in their development is to simply underscore the point that such quantitative or semi-quantitative assessments are possible to construct, along with presumably other yet-to-be-developed assessments. The benefits of these assessments are that the results from a nonexposed risk metric do not have to be taken simply at face value.

Further complicating the process of quality assessment is the fact that two or more processes or phenomena related to the quality of a metric can be occurring at the same time. The ability to assess the quality of nonexposure risk metrics is likely to always be imperfect, but it is also not nonexistent, and we expect that monitoring quality assessment indicators will be a worthwhile pursuit when employing nonexposure risk metrics.

If the Hyperlipidemia hdPS-matched analysis is in fact less confounded than the unmatched analysis, then we suspect that it is most likely that simple random error is the largest contributor to the fact that the central estimate of the briefly-exposed postexposure (bePE) risk is further from the null than that associated with the unmatched analysis. Another possibility would be that the briefly-exposed postexposure (bePE) risk metric subsample is less representative of the full cohort, and potentially overrepresents confounding in the main analysis, as suggested by the greater divergence in the mean propensity scores. However, we have no way to confirm or refute these suppositions, and it is certainly possible that the other nonconfounding influences suggested by one of the other quality assessments could have played a small or large role in the observed briefly-exposed postexposure risk.

If, for some reason, the hdPS-matched analysis is more confounded than the unmatched analysis, we suspect that one potential reason may relate to the fact that we did not individually force different variables for each outcome into the propensity score model (or minimize the number of forced variables altogether). Rather, we had a set of variables that were plausibly the most important for all-cause mortality that were simply also included in the propensity score models for other outcomes. Also, as pointed out in the manuscript, our high-dimensional propensity score models were particularly rich in covariates (including from 1037 variables to 2274 variables). Thus, despite our best efforts, some problematic variables (semi-instrumental variables or collider variables) could have been included in our propensity scores, which are thought to have the potential to increase confounding if unmeasured confounding is present. These possibilities are supposition, however, and there is some evidence that suggesting that the inclusion of problematic variables is not likely to be a major factor in introducing confounding into our models. Specifically, that evidence is that:1) our c statistics were modest (See Supplement 8), 2) all variables included in our propensity scores were required to show at least a +/-20% univariate association with the outcome,^27^ and 3) the briefly-exposed postexposure risk metrics for the unmatched analyses, if they are in fact closely approximating confounding, simply do not suggest much unmeasured or residual confounding is present in our samples (at least considering the central estimates of the briefly-exposed postexposure (bePE) risk metrics). This is important because it would imply that not much unmeasured confounding exists for the problematic variables to increase or “amplify.”

These considerations may suggest that renewed attention should be given to random error as one plausible explanation for the greater divergence from the null of the briefly-exposed postexposure (bePE) risk metric associated with the hdPS-matched analysis than the unmatched analysis. Again, however, we wish to point out that it is the existence of a metric like briefly-exposed postexposure (bePE) risk metric that allows such issues to be highlighted for further attention or investigation.

Obviously, the approach to assessing the quality/accuracy of the briefly-exposed postexposure (bePE) risk metric is still in its infancy. This is clearly a topic for researchers to investigate going forward. This is especially true since no particular single quality assessment (or possibly even a combination of assessments) is likely to be completely definitive. Nevertheless, even though the quality assessments we have presented and discussed in the manuscript and in this Supplement and Supplement 13 are not definitive (and it is possible, given the complexity of phenomena that can affect many of these quality assessments, definitive quality assessments will never be identified), in our view they are valuable. They are expected to help inform investigators about the plausibility of nonconfounding influences affecting the bePE risk metric. In epidemiology, there is a long tradition of relying on informed judgment of investigators is one of multiple tools used to help interpret study findings. Furthermore, the importance of identifying approaches to approximating total confounding (e.g., from measured and unmeasured factors) is so great that approaches can have value even if they are imprecise or sometimes are not valid indicators of confounding. Quality assessments can help inform the judgment of how much weight to ascribe to a briefly-exposed postexposure (bePE) risk metric finding, which in turn can inform the judgment of how much weight to put on a particular main analysis.

**Supplement 15.** **A potential sensitivity analysis to assess for possible attribution bias, postexposure intervention-related effects, and/or depletion-of-susceptible-individuals effects**

If there is sufficient sample and follow-up time in the subsample of individuals informing the briefly-exposed postexposure (bePE) risk metric, then there may be value as a sensitivity analysis in exploring whether the postexposure risk is different for individuals discontinuing the intervention with the briefest exposures compared to those with longer exposures (all falling within the permissible brief exposure allowed). Of course, random error often may be a particular challenge for this sort of comparison, given that a sample that already is smaller than the full study cohort is now being further subdivided.

To elaborate: At least one sensitivity analysis can be envisioned to imprecisely assess whether nonconfounding influences on briefly-exposed postexposure risk may be present. This sensitivity analysis would compare the nonexposure risk between intervention arms observed for the briefly-exposed individuals with the shortest exposure period to nonexposure risk between intervention arms observed for the briefly-exposed individuals with the longest exposure. (All of the exposures would need to fall within the briefly-exposed limit, however.) If there is a substantial change in the postexposure risk observed between the intervention arms when comparing the even more briefly-exposed individuals with those briefly-exposed individuals with longer exposures, this sensitivity analysis might suggest that some type(s) of nonconfounding influence exists. The premise would be that with the briefer exposure, less time would exist for these nonconfounding influences to arise, or at least to have the same impact as would occur with a longer exposure. However, if confounding is extremely time varying, this could also potentially produce a substantial change in postexposure risk. (Nevertheless, in our view, this would have to be a somewhat extreme form of time-varying confounding. For example, confounding would have to be sufficiently more extreme in the earlier days of follow-up that are included in the metric for the individuals with a shorter duration of brief exposure to substantially change the postexposure risk between treatment compared to the individuals with longer durations of brief exposure. These earliest days of follow-up may constitute only ≤ 10% of the total days of postexposure follow-up evaluated in the metric for a given individual. Thus, the change in confounding may need to be substantial to appreciably affect the postexposure risk estimate).

In addition, there potentially could be complex interactions between multiple nonconfounding influences on postexposure risk as the exposure time is varied. This sensitivity analysis, therefore, would not be definitive. However, future research could attempt to assess whether an absence of change in the postexposure risk estimate when shorter versus longer durations of brief exposure are compared generally increases confidence that there may be an absence of substantial contributions to the postexposure risk from nonconfounding influences.

Random error would also be a sizable concern. For instance, in a medication study, if 30-day prescriptions were the limit for “briefly-exposed” individuals, the proportion of individuals receiving prescriptions of considerably less duration than 30 days (e.g., prescriptions of 15 days or less) may be much smaller than the proportion of individuals receiving prescriptions of > 15 days in duration.

To reiterate, in general, the most optimal approach for using postexposure risks to approximate confounding would appear to be to minimize exposure time as much as feasible so as to limit the amount of time for attrition bias, postexposure intervention-related effects, and depletion-of-susceptible-individuals effects to occur. However, once a briefly-exposed sample is chosen (perhaps on the basis that it is the sample possessing the briefest exposure time that might still provide reasonable power, as long as it is felt to be brief enough to sufficiently reduce or minimize these nonconfounding influences), further subdividing the sample as a sensitivity analysis might be helpful. This subdivision might provide at least suggestive evidence to help inform judgments of whether some of these nonconfounding influences are likely to be having a substantial influence on the briefly-exposed postexposure (bePE) risk metric.

**Supplement 16. Considerations that are relevant to concerns that the baseline preexposure measure may not precisely capture confounding at the point of intervention initiation**

Past studies have shown that confounding around the time of intervention for a treatment can be highly time-varying.^9^ This may pose challenges for the baseline preexposure (basePreE) risk metric if sample sizes are not large enough to allow the preexposure period to be very brief. Even if the preexposure period can be defined to be very brief, there are also concerns that this nonexposure metric may not capture the level of time-varying confounding occurring later in the main analysis follow-up period, which would be expected to be often different from that experienced early in the follow-up period. Counterbalancing these concerns at least slightly is the advantage that the baseline preexposure (basePreE) risk metric can often include risks from a large fraction of the full study cohort (and sometimes even the entire study cohort).

To elaborate: As also discussed in Supplement 21, the baseline preexposure (basePreE) risk metric has some distinct advantages and disadvantages relative to other nonexposure risk metrics. For instance, a primary advantage is that it can potentially include all individuals who are in the cohort under study. A disadvantage is that it cannot be used for new onset or terminal conditions, since such conditions will not allow for prior events to occur in the baseline preexposure period to be quantified. Another disadvantage of the baseline preexposure (basePreE) risk metric, discussed in this Supplement, is that risks prior to initiation can be highly time-varying. As a result, there are concerns that the baseline preexposure (basePreE) risk metric may not capture the confounding occurring just at the point of intervention initiation as accurately as desired. In addition, since it does not track confounding over time during follow-up, it may not track confounding occurring much later in the follow-up period as accurately as desired, either.

To put it another way, from a practical perspective the baseline preexposure (basePreE) risk metric needs to include enough events to adequately limit random error. However, in some cases potentially the only way to accomplish this objective is to adopt a preexposure “window” that is so long that the confounding occurring very close to the time of intervention initiation is “washed out” by less dramatic differences in risk between the intervention arms occurring more remotely to the point of intervention initiation. Of course, the briefly-exposed postexposure (bePE) risk metric contains some uncertainties as well, such as the fact it may not capture time-varying confounding occurring extremely early in the follow-up period. Nevertheless, in general we expect that the briefly-exposed postexposure (bePE) risk metric will be less affected by the tendency that confounding can vary to a pronounced degree around the point of medication initiation than the baseline preexposure (basePreE) risk metric. This is speculation, however, and the briefly-exposed postexposure (bePE) risk metric may be more affected by certain nonconfounding influences than the baseline preexposure (basePreE) risk metric. This is also speculation, however, and this subject area could be a topic for further research.

The fact that both the baseline preexposure (basePreE) risk metric and the briefly-exposed postexposure (bePE) risk metric have anticipatable drawbacks in some circumstances is worth noting when using these metrics. In addition, this is a reason to consider using multiple nonexposure risk metrics when feasible (see Supplement 21).

**Supplement 17. The optimal form for the briefly-exposed postexposure (bePE) risk metric effect measure (and other nonexposure risk metrics effect measures) may need exploration**

Understandably, longitudinal studies have not traditionally focused as much on follow-up time that starts at a variable point after exposure initiation. As a result, it is not completely clear what the best effect estimate measure will be to use for various nonexposure metrics.

To elaborate: For convenience, we expressed the briefly-exposed postexposure (bePE) risk metric in our analyses as Hazard Ratios, the same form of effect measure that we adopted for our main analyses. This may not be the best way to express briefly-exposed postexposure (bePE) risk metric, due to the assumption of proportional hazards underlying hazard ratios.

In addition, more nuanced concerns have been raised that measures based on rates, in the strictest sense, can be “noncausal.”^34-36^ Some of these concerns relate to phenomena we already anticipate and discuss in the manuscript, and design our metrics to minimize (e.g., depletion-of-susceptible-individuals effects).

In addition, to choose to avoid using a rate measure for the metric would seemingly ignore the potential impact of differences in follow-up time between the intervention arms on the nonexposure risk metric being examined. For example, in our cohort derivation example, using a relative risk measure in manuscript Table 2 (i.e., a measure that does not take into account differences in follow-up time) suggested that the briefly-exposed postexposure (bePE) risk between the two intervention arms was numerically closer to the null than the rate-based (i.e., hazard ratio-based) measure presented in manuscript Table 3. (We observed a Relative Risk central estimate of 0.99 for the unmatched lithium versus valproate cohorts briefly-exposed postexposure (bePE) risk metric over 365 days in manuscript Table 2. However, in manuscript Table 3 we observed a Hazard Ratio central estimate of 0.91 for the exact same cohorts’ briefly-exposed postexposure (bePE) risk metric over the same time period of 365 days. The difference in value relates to differences in follow-up time between the two intervention arms that exist in the briefly-exposed postexposure sample that is not considered in the Relative Risk measure but is considered in the Hazard Ratio measure).

Further research may be needed to determine the optimal effect measure to use when using the briefly-exposed postexposure (bePE) risk metric, and other nonexposure metrics, to compare risks between two intervention arms.

**Supplement 18.** **Suggested approaches to using the briefly-exposed postexposure (bePE) risk metric in analyses including regression covariates or weighting approaches**

The briefly-exposed postexposure (bePE) risk metric generally examines risk only in a subsample of individuals experiencing the intervention (i.e., only those individuals who are briefly exposed). Nevertheless, when applying the briefly-exposed postexposure (bePE) risk metric to analyses involving weighting or involving regression coefficients, we expect that primarily it will be important to apply the weights or regression coefficients that were used in the main analysis (that is, the analysis based on the final full study cohort) to the briefly-exposed postexposure sample, rather than weights or coefficients derived from the briefly-exposed postexposure sample itself. Although it may seem counterintuitive not to use weights or coefficients derived for the briefly-exposed subsample itself, it must be remembered that the briefly-exposed postexposure (bePE) risk metric is attempting to approximate how well confounding has been resolved in the main analysis. As a result, in our view, in circumstances in which the main analysis involves regression coefficients or weights, we expect it to be more valid to apply the same weights and/or regression coefficient values that are used in the main analysis (based on the final full cohort) in the procedure to determine the briefly-exposed postexposure (bePE) risk metric than to respecify those weights or coefficients based on the briefly-exposed postexposure sample itself.

To elaborate: One unfortunate, but hopefully temporary, aspect of nonexposure risk metrics is that it is not yet clear how well they will work to approximate confounding in analyses that include individual covariates in a regression model or that use weighting (e.g., inverse-probability-of-treatment weighting). In our manuscript, we focused on the use of the briefly-exposed postexposure (bePE) risk metric in analyses using high dimensional propensity score matching. The same basic approach used for matching (that is, using the observed briefly-exposed postexposure risks directly, without modification, as an approximation of confounding) should also be applicable to analyses using stratification. However, some degree of modification to this approach will almost certainly be needed when using the briefly-exposed postexposure (bePE) risk metric to inform analyses using regression covariates or weighting. It is our expectation that the best way to proceed is to apply the weights or coefficients derived from the entire sample for the main analysis to the briefly-exposed postexposure (bePE) subsample as well. To allow the regression coefficients or weights to be respecified for the briefly-exposed sample would seem to potentially underestimate confounding. Put in other terms, to allow coefficients or weights to be re-optimized based just on the briefly-exposed subsample would run the risk of overestimating how well confounding has been resolved (i.e., underestimate confounding).

This approach of using full cohort-derived weights or covariate coefficients should be relatively simple to implement. In a weighted analysis, for instance, it should be a simple task for those individuals who make up the briefly-exposed postexposure (bePE) subsample to retain their weights from the main analysis. After all, all the individuals in the briefly-exposed subsample are also part of the main analysis as well and thus should have weights available from the main analysis. Similarly, although these do not involve values specified at the level of each individual examined in the study, the regression coefficients for each covariate from the main analysis simply could be applied to those same covariates when quantifying the briefly-exposed postexposure (bePE) risk metric estimate. When using this approach, however, concerns about how representative the briefly-exposed postexposure sample is of the full cohort would seem to take on possibly even greater importance.

It is also possible that allowing weights or coefficients to be respecified for the briefly-exposed postexposure (bePE) risk metric and assessing how much they have changed between the full sample and briefly-exposed postexposure (bePE) sample could potentially be another type of “quality assessment” used to evaluate how well the briefly-exposed postexposure (bePE) sample appears to represent the full sample as a whole. Minimal change in the weights or regression coefficients would suggest that the briefly-exposed postexposure (bePE) risk metric is representing the full cohort well. And finally, as pointed out in Supplement 13, there may also be at least some value in deriving the briefly-exposed postexposure (bePE) risk metric both ways, as part of a strategy for gaining maximal information that can help inform judgments of the quality of the briefly-exposed postexposure (bePE) risk metric.

We have not yet tested either of these approaches. (Specifically, we have not tested either the approach of using full cohort-derived weights or regression coefficients or of allowing these weights or coefficients to be rederived from just the briefly-exposed postexposure subsample as a means to assess the representativeness of the sample). However, we present our thinking here about how to use the briefly-exposed postexposure (bePE) risk metric when using these popular analytic strategies to hopefully benefit and accelerate research on this question.

**Supplement 19. Use of nonexposure metrics to validate instrumental variables**

It is widely appreciated that it is very difficult or impossible to confirm whether the assumptions necessary for valid instrument variables (IVs) are actually fulfilled for the analysis being conducted. However, if the briefly-exposed postexposure (bePE) risk metric and other nonexposure risk metrics do serve as useful approximations of confounding, then these may be useful as a possible important “check” of IVs.

To elaborate: Traditionally one approach to evaluating the question of whether IVs have addressed confounding, at least for dichotomous IVs, is to examine the balance seen within a table of baseline characteristics when the sample is stratified based on the presence or absence of the IV. However, an important limitation to this approach is that such a table only allows for assessment in the balance of measured covariates. Yet, the particular strength of the IV approach is its potential to control unmeasured confounding. Use of the briefly-exposed postexposure (bePE) metric or other nonexposure metrics, when valid, would in theory allow for the assessment of how well the IV has resolved total confounding, that is, confounding from both measured and unmeasured factors. The idea would be that if the briefly-exposed postexposure (bePE) risk metric for the IV analysis approximates the null, and there is reason to suspect that nonconfounding influences are not substantially affecting the metric, then this finding could represent important supportive or suggestive evidence that the IV analysis being conducted has in fact effectively reduced or eliminated confounding.

We have not yet tested this approach, but discuss it here given its possible value for potentially strengthening and spreading the use of IVs. It should be noted that this approach seems to highly similar to the rationale and approach used in peer-reviewed methods of instrumental variable validation involving negative control outcomes (for example, the osteoporosis study of Liu and colleagues^37^). We certainly believe that using nonexposure risk metrics in a similar manner to validate IV analysis, as described in this Supplement, is an approach that warrants further investigation.

**Supplement 20.** **The baseline preexposure risk metric may be able to be leveraged to rapidly derive “surveillance-quality” effect estimates from large, diverse samples**

The baseline preexposure (basePreE) risk metric is the only nonexposure risk metric expected to be present for most or all cohort members for many outcomes or studies. As such, in theory it should provide a tool to use for rapidly helping identify candidate interventions for further assessments. In specific, we propose that investigators use optimization methods to iteratively derive a sample to be studied that is the largest subsample available from the overall sample that provides a baseline preexposure (basePreE) risk metric approximating the null. In this manner, both rapid screenings of candidate interventions may be possible, and it may be possible to derive relatively unconfounded effect estimates from a larger, more generalizable sample than otherwise could be conveniently evaluated.

To elaborate: We hypothesize that it should be possible to employ an iterative optimization program using numerical estimation methods to maximize a value measuring, for example, some form of the product of sample size and the inverse of the divergence of the baseline preexposure (basePreE) risk metric from 1.0. (We envision a convergence function that maximizes a value derived from an equation taking some version of the following form: (Number of individuals [in the subsample]/(Absolute value of [1.0 - the baseline preexposure risk metric]). This equation would reach larger and larger values the closer the baseline preexposure (basePreE) risk metric’s value is to 1.0 (i.e., as the denominator got smaller and smaller). In this circumstance, as the denominator got smaller, the value for the overall fraction would get larger, resulting in a larger value for the convergence function. Similarly, the function would have a larger value as the number of individuals included in the subsample became larger. In this manner, to the extent that the baseline preexposure (basePreE) risk metric is an effective surrogate for confounding occurring during follow-up in the main analysis, it might be possible for this approach to identify the largest study cohort from within the overall sample that would be expected to provide a relatively unconfounded intervention effect estimate.

Of note, it is quite possible that multiple different weightings of the numerator or denominator (e.g., multiplying one or the other term by a constant) may need to be explored to provide a convergence function that best obtains a sample that approaches the desired size and minimization of confounding (as approximated by the baseline preexposure (basePreE) risk metric). That is, constant terms may need to be included in the convergence function to increase or decrease the numerator values relative to the denominator values in order to balance an investigator’s priorities to obtain both a sufficiently large subsample and a subsample that minimizes approximated confounding to a degree that the investigator finds optimal (based on what can be achieved in that cohort).

We mention the baseline preexposure (basePreE) risk metric specifically for this function because this nonexposure metric is the only one of our proposed nonexposure metrics that potentially can estimate confounding for the entire cohort being analyzed, or a large fraction of the cohort. If some members of the study cohort lack baseline outcome measures, as pointed out in Table 1 of the manuscript, it may be worth considering performing a subanalysis on just those individuals with such measures, especially if they constitute a majority or sizable majority of the entire cohort. If this same optimization approach was attempted using the briefly-exposed postexposure (bePE) risk metric, we expect that what would result is simply the identification of a particular briefly-exposed subsample that came closest to maximizing the convergence functions value that would not affect the rest of the individuals who were included in the full cohort. Thus, the desire to identify a large sample within the full cohort that can provide relatively unconfounded effect estimates would not be attained.

Also, as pointed out in the manuscript, there are only a limited number of types of outcomes that can be evaluated using the baseline preexposure (basePreE) risk metric. In addition, there are concerns that the metric may not work well when confounding is highly time-varying. Nevertheless, for the outcomes for which the baseline preexposure (basePreE) risk metric is appropriate, this approach may be a very useful tool. This approach may allow the rapid estimation of relatively unconfounded effect estimates across wide, and thus presumably more generalizable, populations. As a result, this approach might also be useful for anticipating the results that might be achieved by a “large simple” or “pragmatic” trial, which deliberately examines an intervention’s effectiveness in broad, heterogeneous populations.

We have not tested this approach, but we hope that we and other researchers can investigate this approach in the future.

**Supplement 21. A rationale to examine multiple nonexposure risk metrics when feasible**

Each nonexposure risk metric has some distinct advantages and disadvantages compared to the others. Thus, when feasible, there may be advantages to assessing whether the findings from multiple nonexposure risk metrics appear to give similar approximations of confounding.

To elaborate: Each nonexposure risk metric has its own distinct advantages and disadvantages. Supplementary Table 2 presents qualitative estimates of various nonexposure risk metrics’ sensitivity to different influences besides confounding. This comparison is based on our current theoretical expectations about how different nonexposure risk metrics might perform, since we have only tested the briefly-exposed postexposure (bePE) risk metric to date. Practically speaking, for many studies current limitations of sample size may dictate that the most feasible nonexposure risk metrics to compare would be the baseline preexposure (basePreE) risk metric and the briefly-exposed postexposure (bePE) risk metric. However, as pointed out in the manuscript, the baseline preexposure (basePreE) risk metric cannot be used for a number of types of outcomes. When the baseline preexposure (basePreE) risk metric cannot

| **Supplementary Table 2. Expected Sensitivity to Different Validity Threats for Nonexposure Risk Metrics During Typical Use** | | | | | |
| --- | --- | --- | --- | --- | --- |
| Potential Validity Threat  (Alternative Influence on the Nonexposure Risk other than Confounding) | Nonexposure Risk Metric | | | | |
|  | Postexposure Nonexposure  Risk Metrics | | | Preexposure  Nonexposure  Risk Metric | Never Exposed  Nonexposure  Risk Metric |
|  | adPE | bePE | ubePE | BasePreE | abNE |
| Unrepresentativeness of Cohort | 0 to +++ | +++ | ++++ | 0 to +++ | +++++ |
| Imprecise Representation of Follow-up Time | +++ | ++ | + | +++++ | 0 |
| Attrition Bias | +++ | + | +/0 | 0 | 0 |
| Postexposure Effects (if present) | +++ | + | +/0 | 0 | 0 |
| Depletion-of- Susceptible-Individuals Effects | +++ | ++ | + | 0 | 0 |
| Regression to the Mean | ? | ? | ? | ? | ? |
| Random Error (compared to Main Analysis) | 0 to +++ | +++ | ++++ | 0 to +++ | +++++ |
| NOTE: The expected strength of each nonconfounding influence is ranked on a scale from 0 (no expected effect) to +++++ (maximum expected effect). It should be stressed that these strength ratings are estimates, and they attempt to estimate “typical” applications of the metric, not all possible manifestations of them. Just as an example, the adPE risk metric (“all discontinuations” postexposure (adPE) risk metric) could be completely nonrepresentative of the full cohort if no individuals discontinue the intervention in the follow-up period (in which case, however, the metric would be valueless and completely uninformative). Or this metric could be completely representative of the individuals in the full cohort if all cohort members discontinue the intervention within the follow-up period under study. (However, in this case while all individuals would be represented in the adPE risk metric, the impact of confounding that they experience over follow-up would still not be completely represented, due to the fact each individual’s postexposure time would begin only after their exposure to the intervention had ended). We expect that both of these extreme scenarios (no representation and complete representation of the cohort being studied in the main analysis), however, are likely to be extremely infrequent. More typically, the adPE risk metric is likely to be rather representative of the study cohort (and more representative than the bePE, ubePE, or abNE risk metrics). A question mark (?) indicates that the phenomenon may or may not be present based on the outcome examined, and that the degree to which it will be present is difficult to predict. | | | | | |

be used, if it is desired to have an additional nonexposure risk metrics to which to compare the briefly-exposed postexposure (bePE) risk metric, it is conceivable that it may even be worthwhile to examine whether the “all discontinuations” postexposure (adPE) risk metric can serve this function. We state this idea with reservations, since in many cases there may be concern that the “all discontinuations” postexposure (adPE) risk metric may be too affected by influences such as attrition bias, postexposure effects, or depletion-of-susceptible-individuals effects to serve this function. However, in those cases in which there is reason to believe that the “all discontinuations” postexposure (adPE) risk metric might be sufficiently free of from nonconfounding influences, then a comparison to the briefly-exposed postexposure (bePE) risk metric might help increase or decrease confidence in the briefly-exposed postexposure (bePE) risk metric findings.

Comparing approximations over two or more nonexposure risk metrics is one approach to deriving a “doubly robust” assessment of confounding for a given analysis. To be specific, the confidence placed in an approximation of confounding would likely increase if a similar approximation of confounding is provided by different methods with different underlying analytic characteristics. Studies employing negative control outcomes, for instance, often examine multiple outcomes, and there have even been efforts to use “double-negative controls” that more formally combine information from negative control exposures and negative control outcomes.^38^ However, future research is needed to determine how often such a combined nonexposure risk metric approach is feasible and how likely it is to improve, rather than potentially confuse, the assessment of confounding for that given analysis. The potential for confusion arises from the fact that one approach may have more limitations than another, and the approaches may also vary in their sensitivity to random error. Thus, divergent approximations of confounding are certainly possible even when all the nonexposure metrics being used are generally free from nonconfounding influences.

Finally, an expanded version of what could be termed “doubly robust” confounding evaluation could be performed by combining information from nonexposure risk metrics with that from negative control outcomes, negative control exposures, self-controlled case series, or other approaches. In fact, further research is needed to develop an appreciation of the role that the briefly-exposed postexposure (bePE) risk metric should play, relative to these other approaches, in approximating confounding. The briefly-exposed postexposure (bePE) risk metric is of interest in part because it seeks to estimate confounding using a subsample of the same individuals being evaluated in the study, for the same outcome that is being evaluated, *and* over some or much of the same time period that the full cohort is actually being evaluated. This congruence with the study population, study outcome, and the follow-up time being studied in the main analysis may be important. However, there are a substantial number of conditions which need to be fulfilled for the briefly-exposed postexposure (bePE) risk metric to yield a close approximation of confounding. We expect that even approximations of confounding that contain some imprecision will be highly useful for researchers.

Nevertheless, other approaches to evaluating or addressing confounding, such as negative control outcomes, negative control exposures, or self-controlled case series have advantages as well. These approaches may be less specific to the population, outcome, and/or time period examined, at least in some cases, but may require fewer conditions to be met for their approximations of confounding to be highly useful. As indicated, combinations of approaches may be particularly useful, as would research investigating the circumstances for which some approaches are more useful than others.

**Supplement 22. Additional thoughts about the value of our proposed nonexposure metrics**

Reaching definitive conclusions about the value of nonexposure risk metrics will likely involve the work of many investigators. Our manuscript proposes four additional nonexposure risk metrics as improvements to the “all discontinuations” postexposure (adPE) metric. We expect that these metrics will be important new approaches for assessing confounding for researchers to investigate further and to employ. Furthermore, our manuscript provides a much more thorough discussion than previous work, to our knowledge, concerning the important considerations that must be considered when applying and interpreting nonexposure risk metrics. In an effort to aid other researchers, this Supplement consolidates and summarizes the key findings from our manuscript supporting the value of the briefly-exposed postexposure (bePE) risk metric and the practical considerations that also suggest these metrics will be valuable. This supplement also summarizes a number of considerations to keep in mind when investigating and employing these nonexposure risk metrics.

To elaborate: We hope that the work described in our manuscript and in these Supplements will be highly valuable to other researchers. In particular, we hope that our proposed nonexposure metrics will provide another valuable means of conducting a type of “negative control” assessment,^1,39^ similar to the accepted procedures for negative control outcomes^1,40^ and negative control exposures.^1,41^

Furthermore, the manuscript does demonstrate the feasibility of an approach that has the potential to quantify both measured and unmeasured confounding *within the sample and time period being evaluated* for the main analysis. In our view, the briefly-exposed postexposure (bePE) risk metric (and the ultra-briefly-exposed postexposure [ubePE] and baseline preexposure [basePreE] risk metric) could be viewed as what can be termed an “internal validation”^21^ or “internal calibration” approach, in contrast to some other “negative control” approaches. Given this ability to quantitatively approximate confounding within a subsample of the primary sample being evaluated, we hope that the briefly-exposed postexposure (bePE) risk metric will constitute an important advance for many intervention and some exposure studies.

Given the potential importance of the briefly-exposed postexposure (bePE) risk metric, it is worth reiterating that the briefly-exposed postexposure (bePE) risk metric is intended to accurately approximate confounding only under certain specific conditions. Those conditions include, in the strictest sense, the absence of 1) attrition bias, 2) postexposure intervention-related effects, 3) any depletion-of-susceptible-individuals effects, 4) regression to the mean, and 5) any differences in time-varying confounding in the brief period not well-measured by the metric. The briefly-exposed subsample of individuals also needs to be adequately representative of the full cohort so that it can provide an accurate approximation of the confounding affecting the full cohort. In addition, strictly speaking, an absence of random error is needed to absolutely ensure the briefly-exposed postexposure (bePE) risk estimate is accurately reflecting confounding. However, some chance for random error is likely to be always present. Fortunately, the likelihood of random error can be estimated and incorporated into assessments using well-established methods.

In a real-world setting, the complete absence of at least some of these influences may be impossible or near-impossible to achieve. Minimization of some or all these influences may be the best that can be typically achieved. Minimization of these influences is precisely what our proposed nonexposure metrics seek to accomplish.

Further complicating the investigation of confounding is the fact that some of these influences, when present, in theory could be “canceled out” by other influences, leading to a briefly-exposed postexposure (bePE) risk estimate close to the null but that is in fact affected by multiple nonconfounding factors. It is not completely clear whether this sometimes could be an advantageous circumstance. Conceivably in some instances this circumstance might allow an accurate approximation of confounding to emerge despite these other influences. Alternatively, in other instances this might be a disadvantageous circumstance. For instance, the presence of multiple nonconfounding influences might make it harder to detect the presence of those nonconfounding influences. And this might be a particular concern at times when attrition bias is one of these nonconfounding influences. Attrition bias would be expected to produce effects on the postexposure risk metrics that are the opposite of its expected effects on the main analysis effect estimate. Thus, conceivably, sometimes “cancelling out” of the effects of nonconfounding influences (especially when one of those influences is attrition bias) might conceal influences on the main analysis, confounding or otherwise, that would be important to recognize. In our view, these uncertainties are a topic worthy of further research.

To aid other researchers who might consider conducting such research, Supplementary Table 3 gives a listing of all the factors that we currently foresee as potentially interfering with the ability of the briefly-exposed postexposure (bePE) risk metric and other postexposure risk metrics to straightforwardly approximate confounding. There certainly may be some additional influences that we have not yet considered.

The reason why we did not present this entire set of potential influences in the main manuscript is because it is not clear how common some of these influences are, and we felt it might overcomplicate many readers’ attempts to understand what was being proposed. We also felt it would be too challenging to present all these potential considerations in a concise manner that would address all the questions that would be posed in some reader’s minds. Finally, we felt like any confusion that might be produced could interfere with some readers’ ability to gain a “big picture” view of some of the most important aspects of nonexposure risk metrics.

**Supplementary Table 3. Processes and Phenomena that can influence Postexposure Risk Metrics^1^**

**such as the briefly-exposed postexposure (bePE) risk metric**

Confounding

Attrition Bias*

Postexposure intervention-related effects*

Depletion-of-susceptible-individuals effects*

Random error

Regression to the mean

Nonrepresentativeness of sample experiencing nonexposure compared to the full sample^2^

Imprecise representation of follow-up time^2,3,^*

Differences in the average starting point or finishing point for the nonexposure period between intervention arms in the presence of time-varying confounding after intervention initiation^3,4,^*

Changes in risk factor prevalence after intervention initiation between intervention arms during either exposure or nonexposure that affects the risk observed in nonexposed individuals^5^

Measurable differences in starting or stopping other interventions between the intervention arms that affect the risks observed for individuals during nonexposure (see Supplement 10)^5^

Unmeasured/unrecognized use of interventions during postexposure time^5,6^

*The asterisk notes those processes or phenomena would be generally expected to be less influential on the postexposure risk if the period of time for intervention exposure was restricted to a briefly-exposed or ultra-briefly-exposed duration, although not necessarily in every case (e.g. some instances of postexposure intervention-related effects, such as vaccines).

^1^ In general, all these processes and phenomenon (other than Confounding) would be expected to cause the postexposure risk metric to approximate more than just the confounding experienced by the study cohort over the follow-up time. Thus, on average, these processes and phenomena would be expected to make the metric a less precise approximation of confounding. (Exceptions are possible, such as a circumstance when the presence of attrition bias raises risks in the postexposure risk metric for Intervention A relative to Intervention B while the presence of postexposure intervention-related effects lowers risks in the postexposure risk metric for individuals discontinuing Intervention A relative to individuals discontinuing Intervention B. In this case, a postexposure risk metric that still closely approximates confounding can sometimes still result, although in addition to any confounding, other biases on the main analysis effect estimate such as attrition bias would still be present).

^2^ Unrepresentativeness or imprecise representation of follow-up time do not directly contribute to postexposure risk, but these phenomena do increase the chance that the measured postexposure risk metric may more imprecisely approximate the confounding observed for the whole cohort over the entire period of follow-up time.

^3^ To clarify, “imprecise representation of follow-up time” refers to imprecision in how well the average duration and timing of the briefly-exposed sample’s postexposure period matches the duration and timing of follow-up evaluated during the main analysis. “Differences in the average starting or finishing point for the nonexposure period between intervention arms” relates to differences in how the nonexposure risk period is represented between the intervention arms.

^4^ In fact, differences in starting or finishing times would produce inaccuracy in the bePE risk metric’s approximation of confounding not from a “nonconfounding” influence technically, but rather from, in cases of time-varying confounding, potentially by representing confounding over slightly different time periods or potentially representing confounding more fully for one intervention arm than the other. However, if a bePE or ubePE risk metric are used, in almost all cases these differences in the starting point of the nonexposure period are expected to be very minor due to the tight constraints on how long the exposure period can last, and thus when the nonexposure period is allowed to start.

^5^ These two phenomena can be viewed as variants of the same phenomenon. That is, one could lump these categories into a broader one involving the change in risk factors for the outcome that occur after initiation of the treatment. Interventions that affect the outcome can be seen as simply one of those risk factors. We have broken these categories out separately in this Table simply because interventions often can have a dramatic effect on the risk of outcome and may warrant particular attention on their own.

^6^ This category simply notes that unrecorded, unrecognized use of the study interventions and/or other interventions during the postexposure period can always occur as well, and this uncertainty is not addressable in the same way that measured changes in other interventions or measured resumption or switching of the study interventions.

In our view, these particularly important aspects of nonexposure risk metrics include: 1) if other influences on the nonexposure risk do *not* exist, the nonexposure risk (within the uncertainties produced by random error) would be expected to directly approximate confounding; 2) however, there are multiple other phenomena that can, in fact, affect nonexposure risk (particularly postexposure risk, which is the basis of metrics such as the briefly-exposed postexposure [bePE] risk metric); 3) as a result, multiple

conditions need to be met for nonexposure risk metrics to most strictly approximate confounding; 4) however, the proposed metrics have been designed to minimize these other influences on nonexposure risk, and also 5) quality assessments are available (and additional assessments can be designed) to help indicate when these other nonconfounding influences may be potentially affecting the nonexposure risk.

Given the array of considerations that might need to be kept in mind when using nonexposure risk metrics, one might ask: why bother with nonexposure metrics at all? As we see it and describe

below, there are several clear reasons for investigating and employing these metrics. These will be discussed specifically in reference to the briefly-exposed postexposure (bePE) risk metric, the nonexposure risk metric that receives the most detailed attention in our manuscript.

1) Real-World Findings Support the Value of the bePE Risk Metric, Especially the Close Correspondance Between Changes in the bePE Risk Metric Findings and the Presumed Changes in Confounding Occurring During Cohort Derivation

First, and of particular importance, our findings suggest that the briefly-exposed postexposure (bePE) metric works well as an approximation of confounding in the examples that we have evaluated and present in the manuscript. Given these encouraging findings, this manuscript probably should be seen as falling somewhere between a simple “demonstration of concept” and a rigorous “proof of concept” of the briefly-exposed postexposure (bePE) risk metric. We use these terms because, while we do have legitimate benchmarks upon which to assess the performance of the briefly-exposed postexposure (bePE) risk metric, these benchmarks are not as rigorous as would be provided by a simulation study or a real-world study with a more firm evidence base supporting the expected size of unconfounded intervention effects. Alternatively, a real-world study that uses other firmly-established “positive” or “negative” controls may also have value for evaluating the briefly-exposed postexposure (bePE) risk metric.

We do find consistency, however, over the multiple analyses that we present in our manuscript to provide an initial assessment of the value of the briefly-exposed postexposure (bePE) risk metric. These analyses include the briefly-exposed postexposure (bePE) risk metric estimates that accompany a) our Study Flowchart Cohort Derivation Diagram, b) our unmatched versus hdPS-matched results in general, and c) quite possibly, our specific analysis of all-cause mortality.

In our view, the strongest evidence provided in our manuscript that supports the use of the briefly-exposed postexposure (bePE) risk metric as a useful approximation of confounding is the observation that the briefly-exposed postexposure (bePE) risk metric appears to consistently track with expected decreases in confounding accompanying logical, well-established cohort derivation procedures (Manuscript Table 2). It should be noted that Manuscript Table 2 presents the greatest changes in the

briefly-exposed postexposure (bePE) risk metric values that are presented anywhere in the manuscript. A large change in confounding during our cohort derivation process is to be expected, given the circumstances (progressing from a completely unrestricted cohort that could plausibly be heavily confounded to a highly restricted study cohort that is likely to be much less confounded). In our view, it is quite noteworthy that this cohort derivation process that would be expected to produce a large change in confounding is accompanied by a large change in the briefly-exposed postexposure (bePE) risk metric. This simple observation deserves strong emphasis. Despite all the conceivable uncertainties that could affect the briefly-exposed postexposure (bePE) risk metric that are mentioned in the manuscript and in these Supplements, the observation remains that, in the setting of our cohort derivation process, the briefly-exposed postexposure (bePE) risk metric does appear to be working largely as desired. Specifically, in this important application of informing or actively aiding cohort derivation, the briefly- exposed postexposure (bePE) risk metric does appear likely to be providing some sort of approximate “window” into confounding.

Additional “real-world” evidence that supports the briefly-exposed postexposure (bePE) risk metric is the fact that in five of the six analyses we presented, the briefly-exposed postexposure (bePE) risk metric suggested that a well-established approach for addressing confounding, high-dimensional propensity score matching, resulted in less confounded findings. This consistency was observed despite the fact that for all the analyses, the unmatched briefly-exposed postexposure (bePE) risk metric findings were already relatively close to the null, increasing the “degree of difficulty” needed for the briefly-exposed postexposure (bePE) risk metric to suggest that benefits do accompany high-dimensional propensity score matching. Out of all the analyses presented (three analyses evaluating all-cause mortality over different time frames and three analyses evaluating 365-day risks for Weight Gain, Hypertension, and Hyperlipidemia outcomes), all of the hdPS-matched analyses except for the Hyperlipidemia analyses were associated with briefly-exposed postexposure (bePE) risks that were closer to the null than those observed for the unmatched analyses. Furthermore, the analysis of the Hyperlipidemia outcome was noteworthy in that it showed the most extreme values for four of our ten briefly-exposed postexposure (bePE) risk metric “quality assessments” (although two of these assessments were closely related). In addition, while the briefly-exposed postexposure (bePE) risk metric associated with the hdPS-matched Hyperlipidemia analysis did not exhibit the widest Confidence Interval Width in our analyses, the width of the Confidence Interval was still quite substantial. Thus, there are indications that perhaps we should view the Hyperlipidemia briefly-exposed postexposure (bePE) risk metric findings as having some important uncertainties.

It should not be overlooked that the briefly-exposed postexposure (bePE) risk metrics for the hdPS-matched all-cause mortality analysis showed findings that were closer to the null for all three time periods than the briefly-exposed postexposure (bePE) risk metrics associated with the unmatched all-cause mortality analysis. Furthermore, the hdPS-matching main analysis, which the briefly-exposed postexposure (bePE) risk metric findings suggested was less confounded than the unmatched main analysis results, showed findings that were closer to the null than the unmatched findings. The actual association of lithium on one- to five-year all-cause mortality, when compared with valproate, is unfortunately not known. However, we strongly suspect our hdPS-matched findings , which are supported by the briefly-exposed postexposure (bePE) risk metric, are closer to the genuine association than our unmatched findings. We also suspect our briefly-exposed postexposure (bePE) risk metric-supported hdPS-matched findings are closer to the genuine association than some prior studies suggesting large (perhaps implausibly large) mortality benefits for lithium.

An unintended disadvantage can sometimes occur from presenting a substantial list of potential caveats or outside factors to consider about an indicator or a model. Such lists (and ours in Supplementary Table 3 is no exception) typically do not present information about either the frequency with which that other factor occurs, nor the typical magnitude of its impact on the indicator or model being discussed. Thus, these potential nonconfounding influences on briefly-exposed postexposure (bePE) risk can seem to present a sobering number of challenges to the basic approach. In actuality, however, the approach may work well in many circumstances. We think the available evidence thus far, discussed in this Supplement and in our manuscript, provides an important indication suggesting that the briefly-exposed postexposure (bePE) risk metric can serve as a helpful approximation of confounding. But future research will be needed to confirm this impression and determine how often or not this is in fact the case.

Indeed, as mentioned before, simulation studies or larger real-world studies would likely provide an even stronger assessment of the value of the briefly-exposed postexposure (bePE) risk metric. We are disseminating this manuscript at this early point to inform other researchers about our work on nonexposure risk metrics. We expect that the study of nonexposure risk metrics will prove an important and valuable undertaking. We believe that this study of nonexposure risk metrics is likely to advance the most rapidly if a number of skilled researchers become interested in this approach and pursue its investigation.

2) The Proposed Nonexposure Risk Metrics Are Designed to Minimize the Influences that can Interfere with using Nonexposure Risks to Approximate Confounding

The second overall reason for using these metrics is that the four new nonexposure risk metrics that we have proposed are designed to, in different ways, minimize these influences that can affect nonexposure risks. Thus, while each of the processes or phenomena mentioned in Supplementary Table 3 can, in theory, influence one or more postexposure nonexposure risk metrics, and cause them to diverge from simply reflecting confounding, how much real-world impact these phenomena make is uncertain. Certainly, one cannot assume that any one of these potential influences will represent a substantial validity threat in every study. Rather, while this list attempts to be somewhat comprehensive, in actual practice some of these conditions may be a particular concern for a particular study, and some may not be a concern.

In particular, the design of the briefly-exposed postexposure (bePE) risk metric is intended to minimize or resolve, by keeping the exposure period very brief, the first three nonconfounding influences listed in Supplementary Table 3. To be specific, the briefly-exposed postexposure (bePE) risk metric is intended to minimize attrition bias, postexposure intervention-related effects, and depletion-of susceptible-individuals effects. However, the degree to which the brief exposure resolves these concerns will vary on a study-by-study or outcome-by-outcome basis. The fourth influence, random error, can be estimated through tools such as 95% confidence limits.

It is less clear to us currently what differences might exist between the various nonexposure risk metrics in their sensitivity to the 5^th^ nonconfounding influence listed, regression to the mean, or whether this is a similar concern for all the metrics. Nevertheless, the potential vulnerability of an analysis to regression to the mean can often be at least partially predicted from knowledge of the study design. (For instance, one might assess whether a reasonable chance exists that some predictor of the outcome will vary substantially and that values of this predictor will be used to help select who receives or does not receive the intervention).

The presence of any nonrepresentativeness of the briefly-exposed postexposure (bePE) sample relative to the full sample (the 6^th^ nonconfounding influence listed) cannot be completely measured (due to the unknown impact of unmeasured factors). Nevertheless, potential nonrepresentativeness of the briefly-exposed sample relative to the full cohort can be quantitatively assessed at least to a partial degree (that is, specifically in reference to measured factors).

Convenient comparisons such as our quality assessments examining the mean propensity score difference between the intervention arms in the briefly-exposed postexposure (bePE) sample compared to the full sample can also help inform judgments of representativeness. If a briefly-exposed postexposure (bePE) risk metric appears to be nonrepresentative of the full cohort, this raises concerns that its approximation of confounding may not entirely apply to the full cohort. On a more basic level, even inspecting what percent of the full sample is represented in the briefly-exposed postexposure (bePE) sample may give a very simple sense of potential representativeness. (For instance, in our analyses, the briefly-exposed postexposure (bePE) sample made up 26-27% of the overall sample, depending on the outcome.) At a minimum, potential nonrepresentativeness should definitely be suspected if the briefly-exposed postexposure (bePE) risk metric is not similar to the full cohort in terms of the frequency of measured covariates. Briefly-exposed risk metric findings and/or findings from the quality assessments examining mean propensity score difference could help highlight which analyses to prioritize for this more in-depth investigation, if resources are limited. Alternatively, one could consider automating the calculation of the change in covariate prevalence between intervention arms (in comparison to the difference observed in the full cohort) accompanying all briefly-exposed postexposure (bePE) risk metric analyses. This would become, in effect, another quality assessment. In addition, while typically the briefly-exposed postexposure (bePE) risk metric would be expected to be less representative of the full cohort than the “all discontinuations” postexposure (adPE) risk metric, in general the briefly-exposed postexposure (bePE) risk metric would be expected to be more representative of the full cohort than the ultra-briefly-exposed postexposure (ubePE) risk metric or the assigned-but-not-exposed (abNE) risk metric.

Regarding the 7th nonconfounding influence listed, imprecise representation of follow-up time, the briefly-exposed postexposure (bePE) risk metric would be expected in general to be less sensitive than the “all discontinuations” postexposure (adPE) risk metric to this influence. The briefly-exposed postexposure (bePE) risk metric would be expected to, on average, usually have an earlier start date for the nonexposure risk period among individuals making up the metric than the “all discontinuations” postexposure (adPE) risk metric (see Supplement 5). This is an important consideration when time-varying confounding is a concern, especially when such confounding is thought to be prominent close to the point of intervention initiation.

Nevertheless, since there is a period of exposure that is allowed for many or most of the individuals making up the briefly-exposed postexposure sample, in situations in which early time-varying confounding is prominent, it is possible such confounding will not be fully captured in the briefly-exposed postexposure (bePE) risk metric. Sometimes there may be concerns that the time-varying confounding between the intervention arms is more or less pronounced very early in the follow-up period under study than later in the follow-up period (when such confounding can be more easily represented by the briefly-exposed postexposure [bePE] risk metric). In this circumstance, it may be necessary to factor into analytic judgments the possibility the briefly-exposed postexposure (bePE) risk might represent a misestimation of confounding or, if possible, determine if other nonexposure risks can be feasibly applied.

The eighth nonconfounding influence listed, differences in the average starting or finishing time of the briefly-exposed postexposure (bePE) risk metric between the intervention arms, can also be specifically measured (see Supplement 5). Furthermore, quite possibly, differences in average starting time between intervention arms may be less pronounced for the briefly-exposed postexposure (bePE) or ultra-briefly-exposed postexposure (ubePE) risk metric than the “all discontinuations” postexposure (adPE) risk metric, given the tight limits of exposure that exist for these two nonexposure risk metrics. However, it should be noted that while the occurrence of differences in starting (or finishing) time between the intervention arms can be quantified, the ramifications of these differences cannot be fully quantified. (However, some degree of sensitivity analysis can be envisioned in which, if sample size allows, researchers deliberately manipulate the discrepancies in starting or finishing times between the intervention arms for nonexposure risk metrics. Subsamples of the individuals informing the nonexposure risk metric could be created to assess how much difference exists in the briefly-exposed postexposure (bePE) risk metric estimates associated with investigator-produced differences in follow-up time).

The final two or three nonconfounding influences listed in Supplementary Table 3 could be, if preferred, grouped into one large category: the possibility of “changes in risk factors post-initiation.” In this broad category, “changes in risk factors” would refer to a) the specific starting or stopping of other interventions affecting the outcomes post-initiation, or b) unmeasured or unrecognized resumption or switching of the study interventions during the postexposure period, or c) changes in other risk factors post-initiation that affect the outcome. We use the term “post-initiation” (of the intervention) rather than simply “postexposure” for considerations “a” and “c” because, in reality, even changes in these risk factors during the current recipient period as well as the postexposure period would influence the postexposure risk in a manner such that it would not simply be reflecting confounding from baseline factors.

Of note, these concerns can be at least partly addressed through choice of censoring conditions for the nonexposure period (as pointed out in Supplement 10). There may also be weighting approaches that can address these concerns, although this is speculation and something we have not yet explored.

As others have pointed out,^16^ unmeasured or unrecognized use of the study interventions could occur if the individual under study is using an intervention intermittently (e.g., receiving a prescription for a medication and using that medication intermittently), either appropriately (e.g. “as needed” use) or inappropriately (e.g., incomplete adherence to recommended treatment). Another possible cause of this influence, unfortunately, is if research is being conducted in a setting in which unrecorded use of the intervention can occur. One example would be a medication study that is conducted in a setting lacking uniformly accessible health care records, such as can occur in many locations lacking universal health care.

The list of processes or phenomena in Supplementary Table 3 may not be complete. Other additional influences may be identified later that also need to be minimized or completely resolved for briefly-exposed postexposure (bePE) risk to most accurately approximate confounding. One additional possibility that might produce nonconfounding influences on the briefly-exposed postexposure risk could relate to the fact that individuals need to “pass through” the briefly-exposed current recipient period without suffering an outcome or a mortality event to become “briefly exposed.” Thus, this intrinsic requirement to “survive through” the briefly-exposed current recipient period conceivably could lead to some degree of nonconfounding differences in the briefly-exposed postexposure (bePE) risk metric. However, how large this effect might be is unclear. (Furthermore, it is possible these effects are quite minor or negligible, unless outcome rates or mortality rates are substantially different in the briefly-exposed current recipient period between the intervention arms). This is speculation, however, and we have not investigated this question. This topic could warrant investigation in future research.

3) The bePE Risk Metric Examines a Subset of Risks Internal to the Sample and Time Period Being Analyzed (How Important an Advantage This Provides Is Uncertain, but It may be Notable and Worth Investigating)

The third overall reason for exploring and using the briefly-exposed postexposure (bePE) risk metric is that there may indeed be an advantage to examining confounding using individuals drawn from within the exact sample and time period, and for the exact outcome, that is being evaluated in the main analysis. How much of an advantage is actually provided by this characteristic of the briefly-exposed postexposure (bePE) risk metric is uncertain and needs to be investigated. Again, the value of the briefly-exposed postexposure (bePE) risk metric compared to other nonexposure risk metrics or other approaches to assessing confounding is likely to vary on a study-by-study, or even on an outcome-by-outcome, basis.

4) Comparing Findings Across Multiple Nonexposure Risk Metrics or Across Multiple Methods to Assess or Address Confounding May Help Minimize the Limitations in any One Method

Fourth, it is worth remembering that, as discussed in Supplement 21, it is likely that over time the most robust practice for assessing confounding will be to compare findings from two or more nonexposure metrics when possible, or from nonexposure metrics and other approaches for assessing or addressing confounding. Thus, no one approach for approximating confounding may need to be perfect, although it is obviously desirable for each approach that is used to be as accurate as possible. It is worth noting that even instrumental variables, one of the best-established methods for addressing confounding that includes unmeasured confounding, require a number of important assumptions that may be only partially testable. This state of affairs only reinforces the point made in Supplement 21 that a combination of approaches, when feasible, may be the best approach. We hope that investigators continue to make increasing use of the growing panel of methods for approximating confounding that includes both measured and unmeasured confounding.

As for the role of nonexposure risk metrics in this growing panel of methods, it may be determined that strengths of the briefly-exposed postexposure risk metric and some other nonexposure include 1) the capacity of these metrics to evaluate confounding with the same study population, study outcomes, and much or all of the same time period examined in the main analysis risk, and 2) the capacity to perform quality assessments to judge how well the nonexposure risk metric may be operating as an approximation of confounding. Although existing quality assessments are not definitive, merely having indicators that can assess some of the conditions which can impair nonexposure risk metric’s ability to accurately approximate confounding may be a relative strength of these methods compared to other methods. Future research will hopefully determine the degree to which this is the case.

Finally, and perhaps most importantly, it is worth considering a perspective that is not unique to the briefly-exposed postexposure (bePE) risk metric but actually applies to all efforts to address the impacts of confounding. This perspective, in our view, is simply that refinements to existing study methodology for assessing or addressing confounding that lead to even small- or moderately-sized incremental improvements may actually ultimately have large benefits for research overall. This is especially true if those advances consistently provide improvements in effect estimates and are consistently implemented. To use the world of finance as an example, small improvements in the valuation of assets, when consistently acted upon, can often yield large gains. Although a single perfect indicator of confounding would be the ideal objective, we expect the consistent application of nonexposure metrics and other approaches to assessing or addressing confounding will improve the overall quality of many intervention studies and some exposure studies. Put another way, we expect that the uncertainties surrounding how well a given nonexposure metric (or other approaches to assessing or addressing confounding) might approximate confounding may prove to be, on average, smaller than the uncertainties produced by conducting intervention studies without their guidance at all. In many circumstances, having an imperfect measure can be much better than having no measure at all. But this point of view needs to be supported or refuted by future research.

As a side note, we note that there may be value in considering the use of large-scale automated approaches or even machine learning to help optimize the use of nonexposure risk metrics. As mentioned in Supplement 13, automated methods and/or machine learning approaches could be used to optimize the development of “quality assessments” for the nonexposure risk metrics and to speed the evaluation of what collection or weighting of quality assessments would be the most helpful. Automated methods could also help speed the application of nonexposure risk metrics (or other approaches to addressing confounding) overall, which in turn could help speed the time it takes to produce a robust sense of the general value of these approaches. Finally, these approaches could help leverage the benefit provided by nonexposure risk metrics or other approaches to assessing or addressing confounding to inform study methodologies intended to reduce confounding. As just one example, compared to non-automated approaches, automated techniques could presumably cycle much more rapidly through variations of analytic methods (e.g., different selection criteria governing which covariates are included in a high-dimensional propensity score model) and assess which approaches should be preferred, using as a “yardstick” the impact of these changes on the chosen indicator or indicators of approximate confounding.

In closing, key topics concerning the briefly-exposed postexposure (bePE) risk metric specifically, and concerning the nonexposure risk metrics in general, are still largely unexplored. Three of these topics are 1) the impact that different nonconfounding influences can potentially have on nonexposure risk metrics, 2) how easily and rigorously the quality of nonexposure risk metrics as approximations of confounding can be assessed, and 3) the potential for the application of large-scale automated approaches or machine learning techniques to analyses featuring nonexposure risk metrics. In addition, we have highlighted a number of specific potential research questions about nonexposure risk metrics in these Supplements. Thus, it is clear that much additional research will be needed to allow these potentially highly valuable approximations of confounding to be used to their fullest advantage. We hope that our manuscript and these Supplements provide a “roadmap” that helps inform efforts to conduct this valuable needed research.
